# A Deterministic Approach to the Dynamics of Visceral Leishmaniasis and HIV Co-infection with Optimal Control

**DOI:** 10.64898/2026.02.24.26346958

**Authors:** S Nivetha, Sunil Maity, A. Karthik, Tanu Jain, Chhavi Pant Joshi, Mini Ghosh

## Abstract

Visceral leishmaniasis (VL) is considerably more severe among individuals infected with human immunodeficiency virus (HIV), leading to higher parasite loads, frequent relapse, and increased mortality. To examine the epidemiological interaction between the two diseases, we develop a comprehensive VL–HIV co-infection model that incorporates transmission pathways, treatment effects, and relapse dynamics. The model is parameterized using real-time data from Bihar, India, including monthly VL-only and VL–HIV co-infected cases and annual HIV prevalence data. Our analysis shows that HIV infection drives the resurgence and persistence of VL even in settings where VL alone would not sustain transmission, underscoring the amplifying effect of HIV-induced immunosuppression on VL dynamics. We further demonstrate that increasing HIV treatment coverage substantially reduces co-infection prevalence and lowers VL relapse rates. Numerical simulations and optimal control analysis highlight the effectiveness of integrated intervention strategies that combine awareness, treatment enhancement, and vector control. Overall, this study emphasizes the need for coordinated VL and HIV control programs and provides data-driven guidance for designing sustainable intervention strategies in endemic regions.

## Introduction

Visceral leishmaniasis (VL), also known as kala-azar, is a major neglected tropical disease (NTD) caused by Leishmania protozoa transmitted through the bite of infected female phlebotomine sandflies [1]. The disease predominantly affects the spleen, liver, and bone marrow, leading to persistent fever, anemia, weight loss, and, if left untreated, high fatality. A notable clinical consequence of VL is post–kala-azar dermal leishmaniasis (PKDL), a chronic dermal condition that can serve as an important reservoir for ongoing transmission in endemic regions [2]. Human immunodeficiency virus (HIV), the causative agent of acquired immunodeficiency syndrome (AIDS), is transmitted primarily through unprotected sexual contact, contaminated needles, and mother-to-child exposure [3]. VL–HIV co-infection is increasingly recognized as a serious public health challenge, as each pathogen accelerates the progression and severity of the other, resulting in higher parasite loads, frequent relapses, and increased mortality [4]. Once infected, individuals experience progressive immune suppression, which greatly increases vulnerability to opportunistic infections, including VL, thereby creating a biologically synergistic and clinically severe co-infection scenario in endemic regions.

Understanding the complex interaction between VL and HIV, and their combined impact on disease burden, has motivated the use of mathematical modeling to study transmission dynamics and assess control strategies. Mathematical modeling offers a powerful framework to quantify disease spread, explore co-infection pathways, and evaluate the effectiveness of targeted interventions. Previous models of VL [5, 6] have examined seasonal variation and transmission across diverse epidemiological settings, highlighting the importance of vector-targeted strategies. Compartmental approaches have further investigated human–animal–vector interactions [7], the contribution of post–kala-azar dermal leishmaniasis (PKDL) to sustained transmission [8, 9], and the critical role of early PKDL detection in preventing disease resurgence [10]. Additional studies emphasize cost-effective diagnostics [11], timely case detection [12], and vector control [13] as essential components of successful VL elimination programs. For HIV, mathematical models have analyzed transmission dynamics, therapeutic strategies, and the potential for long-term remission [14, 15, 16, 17]. These studies highlight the importance of accurate parameterization, preventive measures, and the optimization of antiretroviral therapy (ART).

VL–HIV co-infection was first reported in India in 1999, with most cases concentrated in the state of Bihar [18]. Although the overall prevalence remains relatively low (0.029–0.4%), co-infection markedly accelerates disease progression and leads to poorer treatment outcomes. Existing models of VL–HIV dynamics [19, 20, 21] emphasize the influence of transmission and recovery parameters, the contribution of PKDL to sustained transmission, and the disproportionately higher VL incidence among co-infected individuals. Clinical evidence further demonstrates severe cutaneous and mucocutaneous manifestations in HIV-positive patients with VL [22, 18].

Together, these findings indicate that HIV-induced immunosuppression exacerbates VL pathology, while VL infection accelerates HIV disease progression, creating a mutually reinforcing cycle. Despite extensive work on VL and HIV individually, relatively few studies have examined their synergistic interaction at the population level. To address this gap, we develop a compartmental VL–HIV co-infection model that explicitly incorporates HIV treatment, its immunosuppressive effects, and a suite of optimal control strategies designed to evaluate the impact of targeted interventions.

Co-infection of VL and HIV poses a serious public health challenge in regions where the two diseases overlap, particularly in Bihar, India, which continues to report a substantial VL burden despite prolonged elimination efforts [23]. In this study, we integrate real-time epidemiological data using monthly VL and VL–HIV case reports from Bihar, along with annual HIV prevalence data from the National AIDS Control Organization (NACO), to calibrate and validate our model. In addition to a treatment compartment for HIV, we introduce control variables that represent community awareness campaigns, increased adherence to antiretroviral therapy, expanded use of protective bed nets to reduce exposure to sandflies, and intensified vector control measures that increase sandfly mortality. These controls allow us to capture how behavior change, education, and vector-targeted strategies interact with clinical treatment to influence co-infection dynamics.

Through this integrated approach, we demonstrate that HIV substantially amplifies VL transmission dynamics, leading to a resurgence of VL cases among co-infected individuals. The immunosuppressive effect of HIV increases parasite persistence, elevates infectiousness, and contributes to repeated VL episodes, thereby undermining ongoing elimination efforts. However, our findings also show that strengthening HIV treatment particularly increasing ART uptake and adherence plays a pivotal role in reducing VL relapse rates and improving outcomes in co-infected populations. These results provide evidence based insights for policymakers on the effectiveness of coordinated strategies that combine education, treatment, and vector management. More broadly, this work contributes to the growing literature on neglected tropical diseases, illustrating how co-infection dynamics can complicate elimination programs and underscoring the need for data-driven, region-specific modeling. Ultimately, our model bridges theoretical epidemiology with practical public health application, supporting more sustainable and targeted control efforts in VL endemic settings.

Overall, this study provides a data-driven modeling framework that captures the bidirectional effects of VL–HIV co-infection and evaluates a comprehensive set of intervention strategies tailored to an endemic region. This paper is structured as follows: Section 1 details the model formulation; Sections 2 and 3 analyze the VL-only and HIV-only models; Section 4 examines the co-infection model; Section 5 introduces the optimal control framework; and Section 6 concludes with key insights and future research directions.

## 1 Mathematical Model Formulation

This section presents the mathematical formulation of the VL-HIV co-infection model. The human population is divided into 19 compartments to capture the transmission dynamics of both diseases, while three additional compartments represent VL transmission in sandflies. Following the VL transmission mechanism proposed in [24], we structured our model accordingly. The descriptions of the variables and parameters are provided in Tables 1, 2 and 3.

**Table 1.** Descriptions of variables for the co-infection model.

| Variable | Description |
| --- | --- |
| $N$ | Total human population |
| $S$ | Susceptible humans |
| $L_L$ | Latent VL-infected humans |
| $I_L$ | Actively VL-infected humans |
| $I_D$ | Dormant VL-infected humans |
| $I_P$ | PKDL-infected humans |
| $R_L$ | Recovered from VL |
| $I_H$ | HIV-infected humans |
| $I_A$ | AIDS-infected humans |
| $I_{HLL}$ | Latent VL-infected humans with HIV |
| $I_{ALL}$ | Latent VL-infected humans with AIDS |
| $I_{HL}$ | Co-infected with VL and HIV |
| $I_{AL}$ | Co-infected with VL and AIDS |
| $I_{HD}$ | Dormant VL-infected humans with HIV |
| $I_{AD}$ | Dormant VL-infected humans with AIDS |
| $I_{HP}$ | PKDL-infected humans with HIV |
| $I_{AP}$ | PKDL-infected humans with AIDS |
| $T$ | Individuals receiving HIV treatment |
| $N_S$ | Total sandfly population |
| $S_S$ | Susceptible sandflies |
| $E_S$ | Exposed sandflies (VL incubation) |
| $I_S$ | VL-infected sandflies |

**Table 2.** Parameter descriptions for the co-infection model.

| Parameter | Description |
| --- | --- |
| $\Lambda$ | Recruitment rate of humans |
| $\mu$ | Natural death rate of humans |
| $c$ | Transmission probability from an infectious human to sandflies |
| $b$ | Transmission probability per bite from infected sandflies to humans [27] |
| $\beta$ | VL transmission rate |
| $\beta_H$ | HIV transmission rate |
| $\lambda$ | Rate at which latent VL progresses to active VL infection |
| $\theta$ | Rate at which VL-infected individuals enter the dormant state |
| $\omega$ | Rate at which dormant VL progresses to PKDL |
| $\gamma_P, \gamma_D, \gamma_L$ | Recovery rates of humans from PKDL, dormant VL, and latent VL, respectively |
| $\Omega$ | Rate of waning immunity (transition from $R$ to $S$) |
| $\xi_1, \xi_2$ | Rates at which VL-dormant individuals with HIV and AIDS enter the co-infected compartment due to VL reactivation |
| $\nu$ | Rate at which HIV infection progresses to AIDS |
| $\psi$ | Rate of ART treatment initiation for HIV-infected individuals |
| $\tau$ | Rate at which individuals discontinue treatment and return to $I_H$ |
| $\sigma_L$ | VL-related death rate |
| $\sigma_A$ | AIDS-related death rate |
| $\Lambda_s$ | Recruitment rate of sandflies |
| $\lambda_s$ | Rate at which VL-exposed sandflies become VL-infected |
| $\mu_s$ | Natural death rate of sandflies |

**Table 3.** Modification parameters and their ranges.

| Parameters | Description | Range |
| --- | --- | --- |
| $\alpha_H$ | Modification parameter for variation in HIV and AIDS transmission | $\alpha_H < 1$ |
| $\alpha_S, \omega_S$ | Modification parameters for transmission from co-infected compartments | $1 < \omega_S < \alpha_S$ |
| $\epsilon_i, i = 1, 2, 3, 4$ | Modification parameters associated with $f_H$ | $1 \leq \epsilon_1 \leq \epsilon_2 \leq \epsilon_3 \leq \epsilon_4$ |
| $\delta_i, i = 1, 2, 3, 4$ | Modification parameters associated with $\nu$ | $1 \leq \delta_1 \leq \delta_2 \leq \delta_3 \leq \delta_4$ |
| $\eta_i, i = 1, 2$ | Modification parameters associated with $f_L$ | $1 \leq \eta_1 \leq \eta_2$ |
| $\alpha_i, i = 1, 2$ | Modification parameters associated with $\lambda$ | $1 \leq \alpha_1 \leq \alpha_2$ |
| $\kappa_i, i = 1, 2$ | Modification parameters associated with $\theta$ | $1 \leq \kappa_1 \leq \kappa_2$ |
| $\rho_i, i = 1, 2$ | Modification parameters associated with $\omega$ | $1 \leq \rho_1 \leq \rho_2$ |
| $\zeta_i, i = 1, 2$ | Modification parameters associated with $\gamma_P$ | $\zeta_2 \leq \zeta_1 \leq 1$ |
| $\varphi_i, i = 1, 2$ | Modification parameters associated with $\gamma_L$ | $\varphi_2 \leq \varphi_1 \leq 1$ |
| $\psi_i, i = 1, 2$ | Modification parameters associated with $\gamma_D$ | $\psi_1 \leq \psi_2 \leq 1$ |
| $\phi_i, i = 1, 2, 3, 4$ | Modification parameters associated with $\psi$ | $\phi_1 \leq \phi_2 \leq \phi_3 \leq \phi_4 \leq 1$ |

We have also incorporated ART treatment for HIV-infected individuals, as evidence suggests that VL mortality is significantly reduced by 64 % *to* 66 % among those receiving ART compared to those without it. This approach is essential for preventing VL relapses, particularly in patients with low CD4+ T-cell counts, who remain at high risk despite secondary prophylaxis and effective initial VL treatment. Considering treatment effects allows our model to more accurately capture the interaction between HIV and VL [25, 26].

Additionally, VL can persist in a dormant state within infected individuals, remaining inactive until they acquire HIV or progress to AIDS, at which point VL is reactivated, leading to relapse. This reactivation in HIV or AIDS patients adds to the complexity of co-infection dynamics, highlighting the necessity of ART and other intervention strategies in our model.

Using the description of parameters in Tables 1, 2 and 3, along with the flow diagram in Figure 1, we formulated a system of differential equations to represent these dynamics as follows:

**Fig 1.**
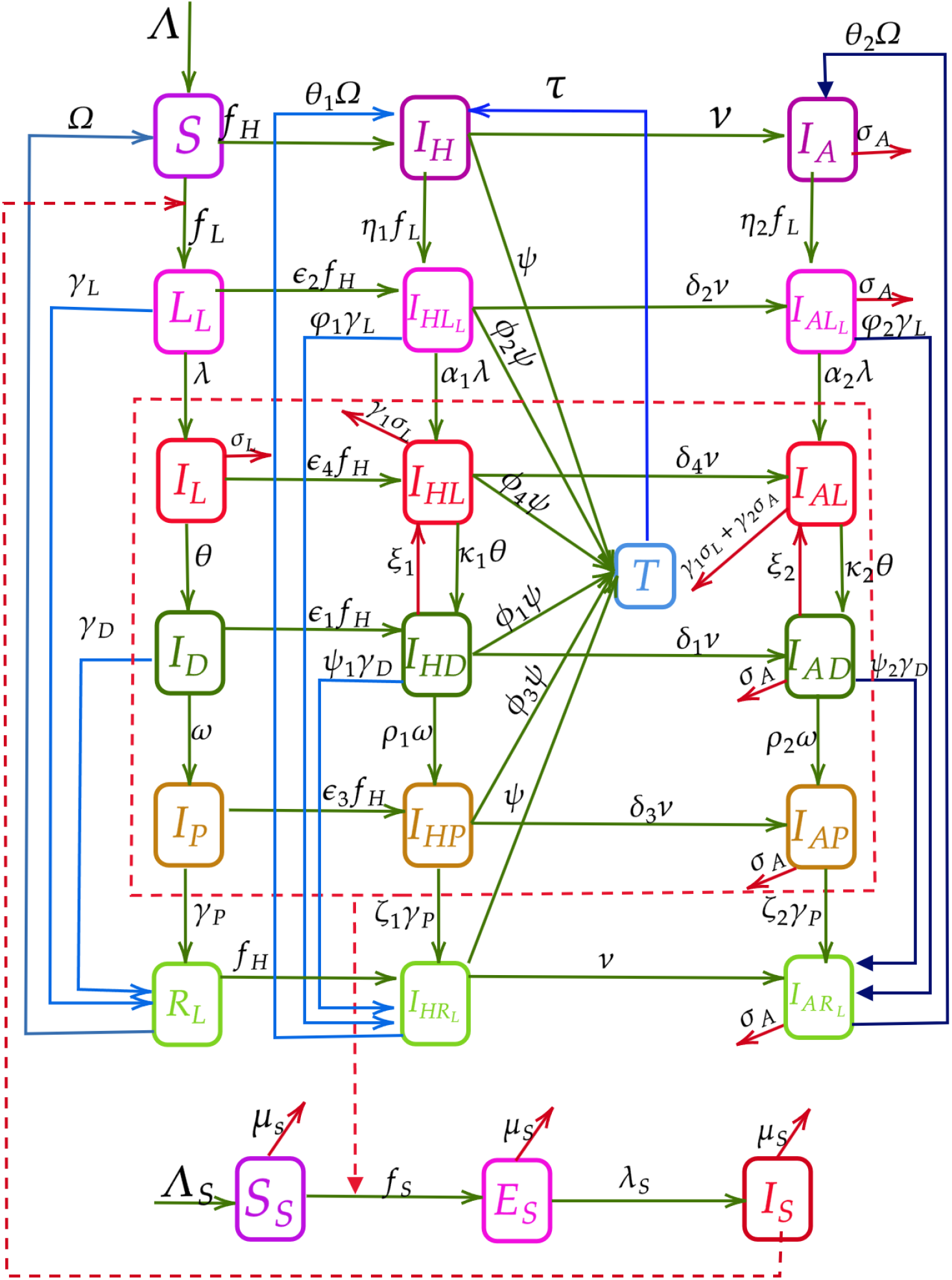
Flow diagram representing the transmission dynamics of HIV and Visceral Leishmaniasis (VL) co-infection.

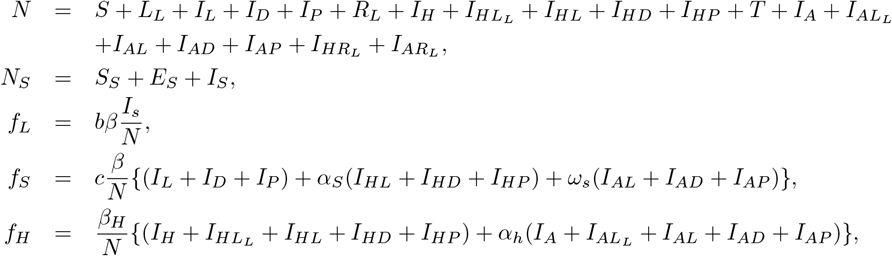

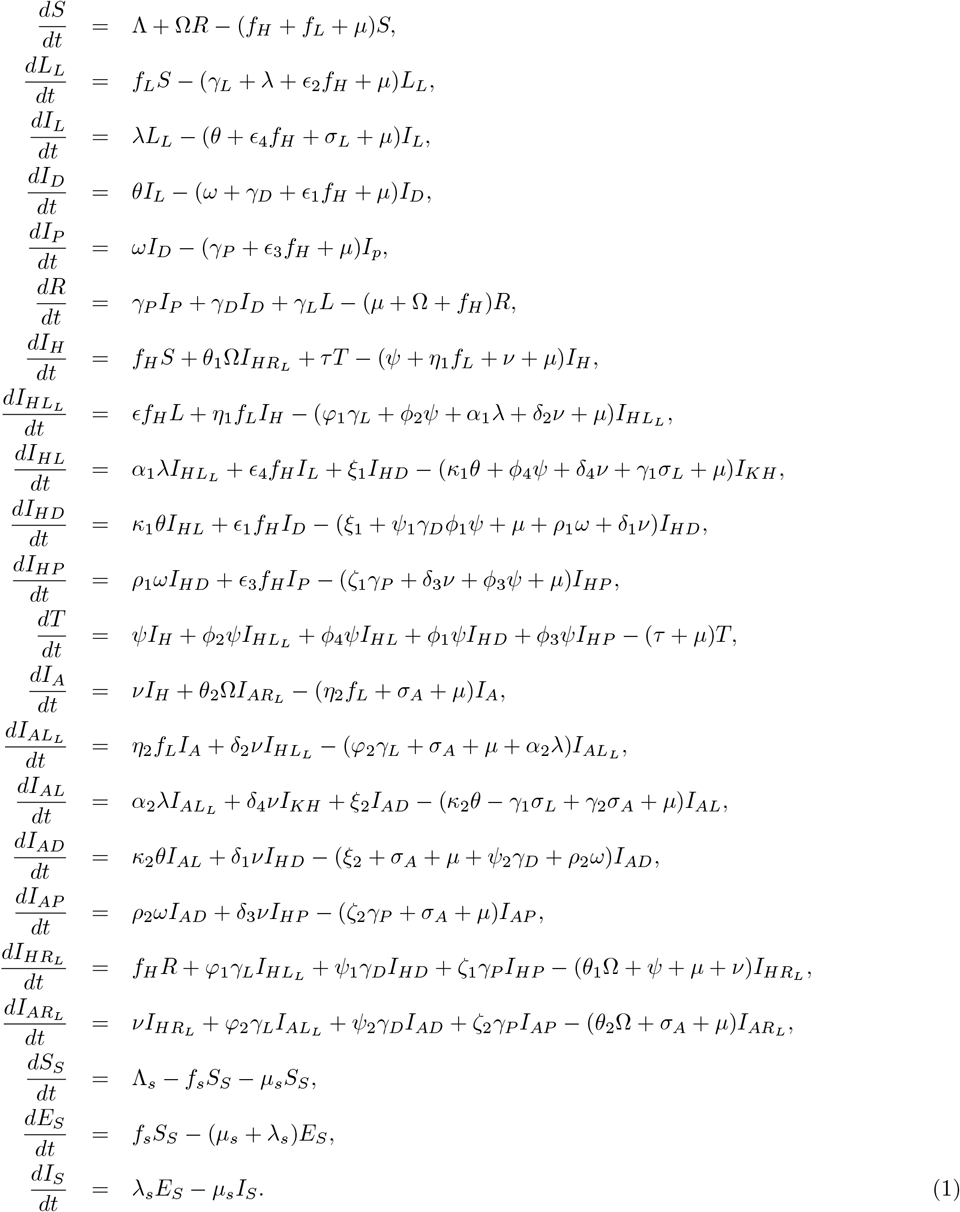

## 2 Analysis of VL only model

### 2.1 Formulation of Model

The VL-only sub-model is derived by setting all HIV-related and co-infected compartments to zero, that is, 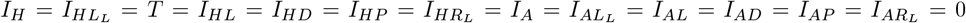, so that only VL transmission dynamics remains active in the system. This reduction isolates the VL-specific pathways by removing HIV progression and co-infection effects, allowing us to analyze the behavior of VL in the absence of HIV. solving the equation for the remaining compartments.

**Flow diagram of VL only Model**

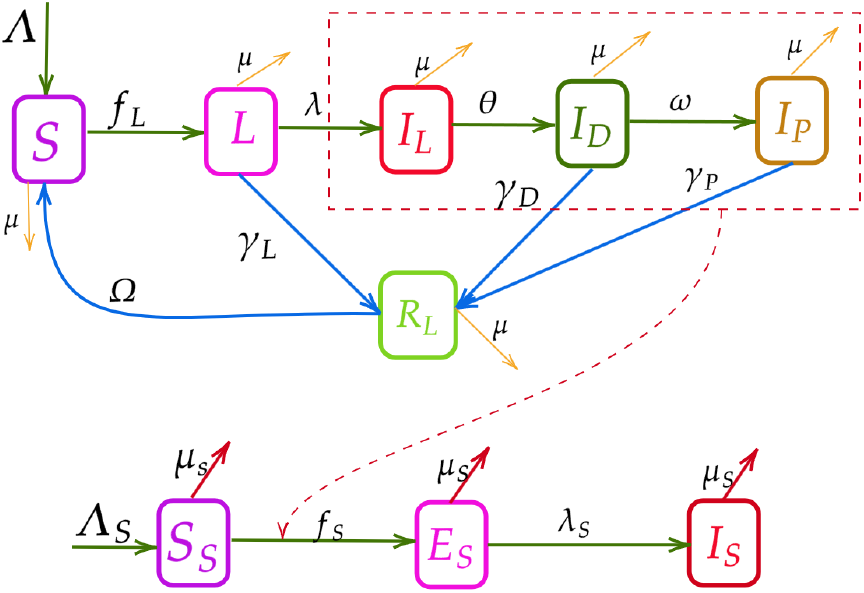

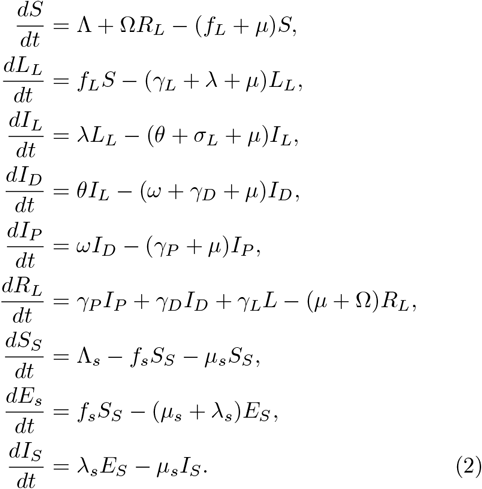

### 2.2 Positivity and the Bounded Nature of the Solution

From (2), we obtain the following

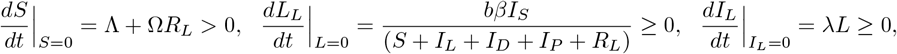

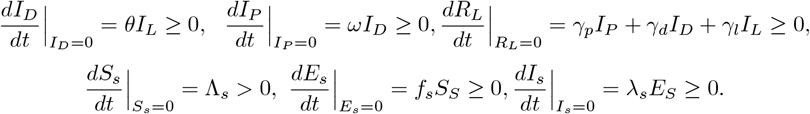

All transition rates in the model are non-negative on the boundary planes. Hence, any solution that begins in the interior of the non-negative bounded cone 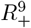, will remain inside this region. This follows from the fact that the vector field points inward along every boundary surface, preventing trajectories from leaving the feasible region. As a result, the state variables remain non-negative for all time. Moreover, consistent with the compartmental structure of the model, the total population *N* = *S* + *L*_*L*_ + *I*_*L*_ + *I*_*D*_ + *I*_*P*_ + *R*_*L*_ satisfies,

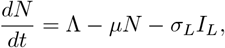

and

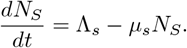

This gives lim 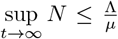, lim 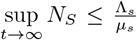. Hence every solutions *S*(*t*), *L*_*L*_(*t*), *I*_*L*_(*t*), *I*_*D*_(*t*), *I*_*P*_ (*t*), *R*_*L*_(*t*) are bounded by 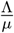 and the solutions *S*_*S*_(*t*), *E*_*S*_(*t*), *I*_*S*_(*t*) are bounded by 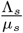 This gives us the biological feasible region of the system (2) by the below positively invariant set is given as

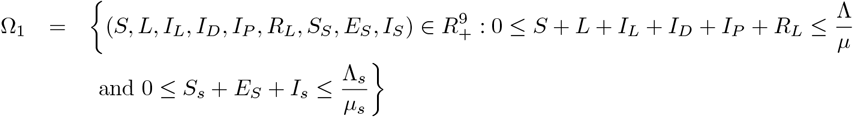

### 2.3 Existence of Equilibrium Points and Basic Reproduction Number

#### 2.3.1 Disease-free Equilibrium

For the subsystem with only VL (2), the disease-free equilibrium (DFE) corresponds to the state in which no individuals are infected with VL and the population remains entirely susceptible. Biologically, the DFE represents a scenario in which VL does not persist in the community and transmission is completely absent.

The disease-free equilibrium for our model is given by

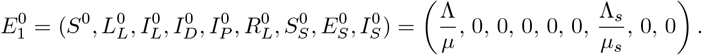

#### 2.3.2 The Basic Reproduction Number

We compute the basic reproduction number (*R*_0*L*_) for the model using the next generation matrix method [28, 29, 30]. In this framework, the new infection and transition matrices, denoted by *F*_1_ and *D*_1_, respectively, are constructed for the infected compartments *L*_*L*_, *I*_*L*_, *I*_*D*_, *I*_*P*_, and *I*_*S*_.

The explicit forms of *F*_1_ and *D*_1_, together with their corresponding Jacobian matrices *F*_1_ and *V*_1_, are provided in the supporting document.

The next generation matrix is then given by 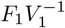, whose dominant eigenvalue represents the basic reproduction number.

After simplification, the basic reproduction number for the VL-only subsystem is obtained as

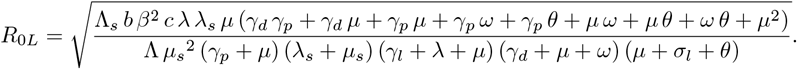

Biologically, *R*_0*L*_ represents the expected number of secondary VL infections generated by a single infected individual in a fully susceptible population. Thus, VL can invade and persist if *R*_0*L*_ > 1, whereas *R*_0*L*_ < 1 implies eventual disease elimination.

##### Theorem 1.

*The disease-free equilibrium* 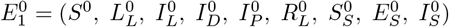 *is locally asymptotically stable if R*_0*L*_ < 1, *under some restriction on the parameters otherwise it is unstable*.

*Proof*. Proof of the theorem is available in supporting information file.

#### 2.3.3 Existence of an Endemic Equilibrium Point

This is the point in the system where the illness still exists. Consequently, 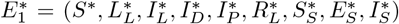 is the endemic equilibrium of the system (2), is obtained by equating system (2) to zero.

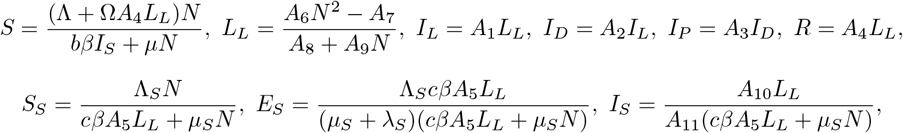

*N* is a positive root of the following equation

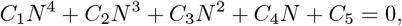

where

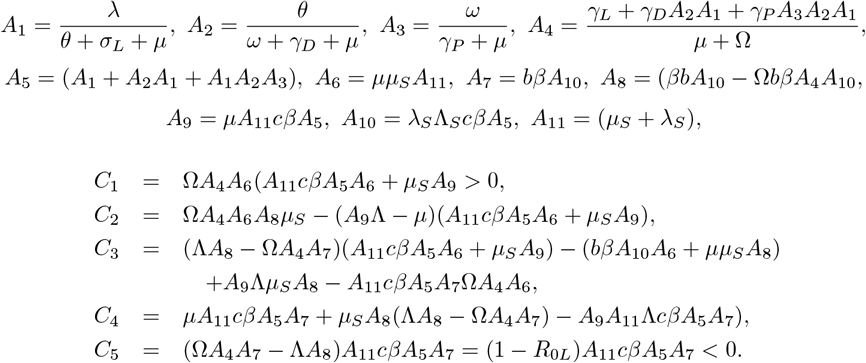

Since the leading coefficient *C*_1_ is strictly positive. By continuity of the polynomial, the function must cross the horizontal axis at least once for some *N >* 0. Therefore, the equation admits at least one positive real root.

### 2.4 Bifurcation and Stability Analysis of the Endemic Equilibrium

The detailed derivations are presented in supporting information file.

According to the theorem discussed in [31], since *a <* 0 and *b >* 0, the system admits a unique VL-only endemic equilibrium point, denoted by 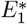, whenever *R*_0*L*_ > 1. Hence, we state the following theorem.

#### Theorem 2.

*The unique endemic equilibrium* 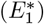 *of system* (2) *is locally asymptotically stable whenever R*_0*L*_ > 1.

### 2.5 Numerical Analysis

This section presents the results of various numerical simulations, including model solutions, data fitting, parameter estimation, and analysis of disease prevalence. A detailed discussion of these findings is also provided. The model has been calibrated using visceral leishmaniasis (VL) data from the state of Bihar.

#### 2.5.1 Data Fitting

Data fitting with optimally estimated parameters enables the identification of key factors governing disease transmission dynamics. In this study, model fitting and parameter estimation were performed using reported visceral leishmaniasis (VL) case data from Bihar, India. Through model calibration, seven key parameters were estimated, including the biting rate of sandflies (*β*), recovery rates from latent (*γ*_*L*_), infected dormant (*γ*_*D*_), and PKDL-infected (*γ*_*P*_) compartments, waning immunity rate (Ω), progression rates from latent to infectious (*λ*) and from infectious to dormant stages (*θ*), and transmission probabilities from sandfly to human and human to sandfly (*b, c*). A few additional parameters were adopted from existing literature and assumed within biologically plausible ranges to ensure epidemiological realism and model consistency. All parameter values are summarized in Table 4.

**Table 4.** Parameter values.

| Parameter | Description | Value | Source |
| --- | --- | --- | --- |
| $\beta$ | Biting rate of sandflies | 3.9384 | Fitted |
| $b$ | Transmission probability per bite from infected sandflies to humans | 0.033 | Fitted |
| $\sigma_L$ | Death due to disease | 0.3 | Assumed |
| $\Omega$ | Rate of waning immunity | 0.4052 | Fitted |
| $\gamma_L$ | Recovery rate of humans from latent VL | 0.4109 | Fitted |
| $\lambda$ | Rate at which $L$ progresses to $I_L$ | 0.049 | Fitted |
| $\theta$ | Rate at which $I_L$ individuals enter into $I_D$ | 0.6 | Fitted |
| $\omega$ | Rate at which individuals in $I_D$ enter into $I_P$ | 0.016 | [33] |
| $\gamma_D$ | Recovery rate of humans from $I_D$ | 0.95 | Fitted |
| $\gamma_P$ | Recovery rate of humans from $I_P$ | 0.7013 | Fitted |
| $c$ | Transmission probability from infectious humans to sandflies | 0.7356 | Fitted |
| $\lambda_s$ | Rate at which sandflies in $E_S$ move to $I_S$ | 0.5 | [34] |
| $\mu_s$ | Death rate of sandflies | 2.17 | [34] |
| $R_{0L}$ | Basic Reproduction number for VL only model | 1.1039 | |

Since the available dataset contains only the number of reported infected cases, the unobserved state variables latent VL (*L*_*L*_), infected dormant (*I*_*D*_), PKDL-infected (*I*_*P*_), and recovered individuals (*R*) were indirectly estimated through model simulations. The initial conditions for the VL-only model compartments were set as *L*(0) = 449.443, *I*_*l*_(0) = 204.000 (fixed from data), *I*_*d*_(0) = 264.919, *I*_*p*_(0) = 202.877, and *R*_*l*_(0) = 1033.764. The model was numerically implemented and fitted using the Python programming language. Parameter values were optimized through the Maximum Likelihood Estimation (MLE) [32] method to ensure that the simulated outputs closely matched the observed active case data. The fitted curve, presented in Figure 2, shows strong agreement between the observed and model-predicted values, indicating that the estimated parameters effectively capture the underlying VL transmission dynamics.

**Fig 2.**
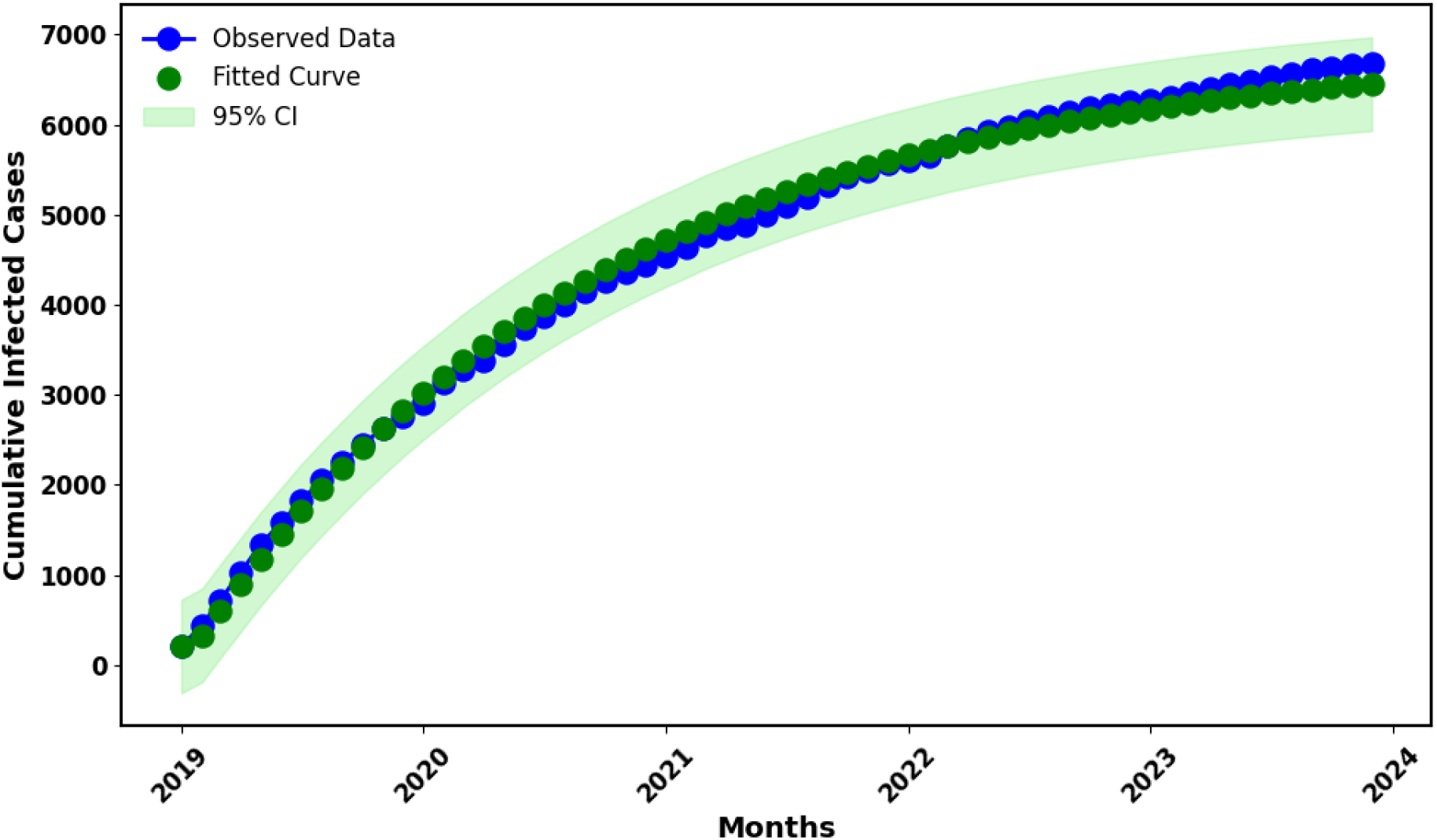
Data fitting plot for the VL-only model. The green dots represents the model fit, while the blue dotted line indicate the observed cumulative monthly visceral leishmaniasis cases of Bihar.

### 2.6 Sensitivity Analysis

Sensitivity analysis is an essential method for assessing the reliability of the findings of an epidemiological study. In order to determine how significant parameters affect the fundamental reproduction number (*R*_0*L*_), they must be changed within a specific range. Important factors in the VL model that have a substantial impact on (*R*_0*L*_) include *β, µ*_*S*_. We determined the ratio of the relative change in the variable to the relative change in the parameter [35] in order to create the normalized forward sensitivity index of a variable with respect to a parameter.

With regard to the basic reproduction number (*R*_0*L*_), Figure 3 demonstrates that the parameters *β, b, µ*, Λ_*s*_, *c, λ*_*S*_, *λ* have positive sensitivity indices, indicating that they play a significant role in increasing *R*_0*L*_. Conversely, the other parameters have negative sensitivity indices, meaning they contribute to a decrease in *R*_0*L*_. Biologically, this implies that an increase in the biting rate, transmission probabilities, or recruitment of susceptible individuals enhances disease transmission. Conversely, parameters associated with recovery or natural mortality reduce the potential for visceral leishmaniasis spread in the population.

**Fig 3.**
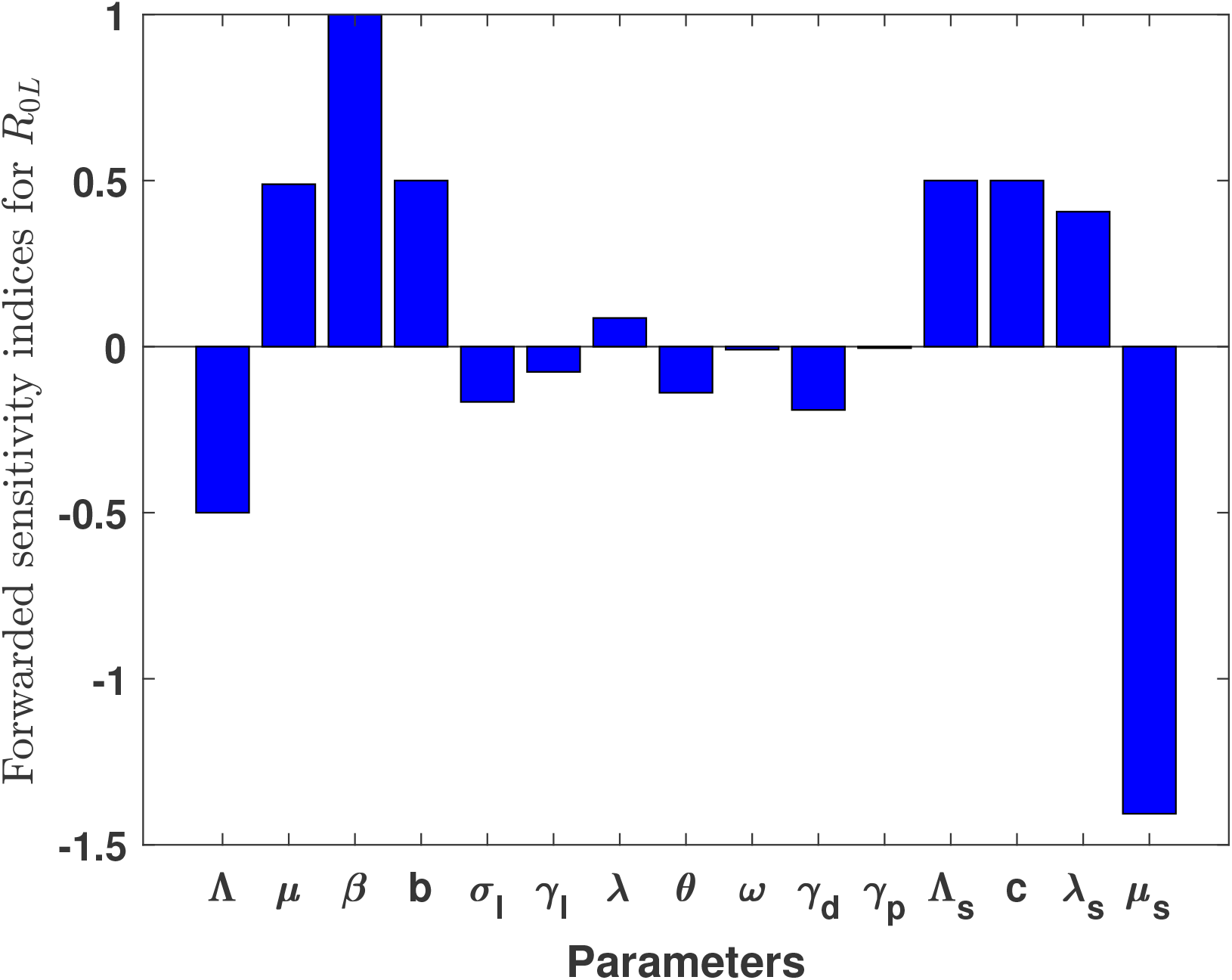
Forward sensitivity analysis of the VL only model with respect to the basic reproduction number (*R*_0*L*_) using the parameter values in Table4.

**Fig 4.**
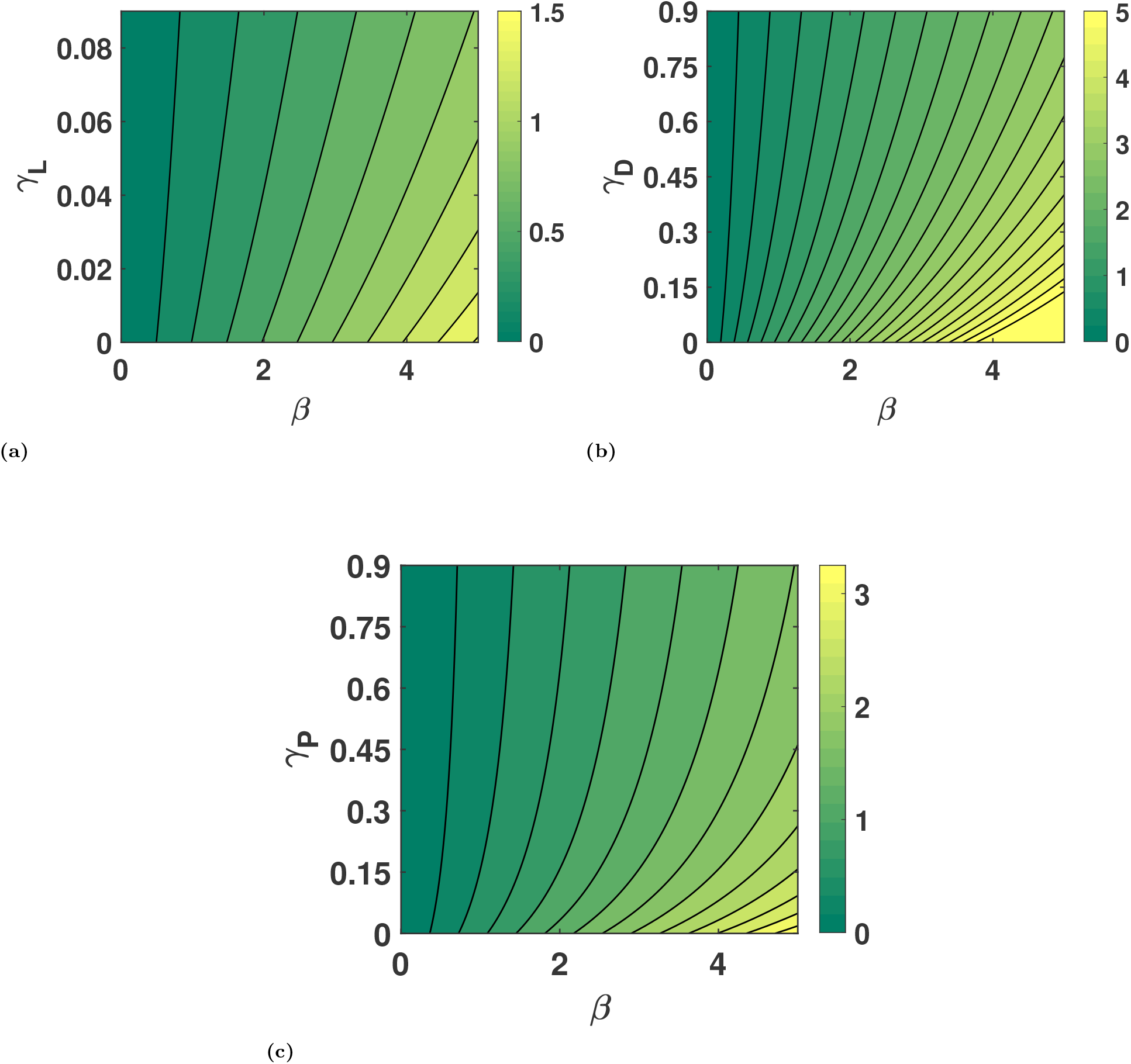
Contour plots of *R*_0*L*_ for different combinations of parameter pairs (a) (*β, γ*_*L*_), (b) (*β, γ*_*D*_), and (c) (*β, γ*_*P*_), with all other parameters fixed as shown in Table 4.

For system (2), we perform a global sensitivity analysis using the technique described in [36, 37]. The response functions are VL only infected *I*_*L*_, VL dormant *I*_*D*_, PKDL infected *I*_*P*_ and Infected sandflies *I*_*S*_. We used the following parameters as inputs:Λ, *µ, β, b, c, σ*_*L*_, Ω, *γ*_*L*_, *λ, θ, ω, γ*_*D*_, *γ*_*P*_, Λ_*S*_, *µ*_*s*_, *λ*_*s*_. We used partial rank correlation coefficients (PRCCs) and Latin Hypercube Sampling (LHS) as our two statistical approaches. While the latter links the input parameters and response function, assigning values between −1 and 1, the former allows variations of several parameters together in a systematic manner. The PRCC sign indicates the kind of correlations that exist between the response function and the input parameters, while the values of the correlation indicate their strength. As a prerequisite for computing PRCCs, nonlinear and monotone relationships were identified between the input parameters and the symptomatic infectious population. 1000 simulations are conducted for each LHS, taking into account a uniform distribution for each input parameter. 25% of the nominal values of the parameters are expected to be exceeded.

The Partial Rank Correlation Coefficient (PRCC) values were estimated to measure the sensitivity of each model parameter with respect to the corresponding response function while accounting for the effects of other parameters. When evaluating a parameter’s role in model prediction and its accuracy, it is important to consider both the magnitude and direction of the PRCC values. PRCC values [38] are significant when they are greater than 0.5 (indicating a strong positive influence) or less than −0.5 (indicating a strong negative influence). Figure 5a illustrates that the parameters Λ, *σ*_*L*_, *λ*, and *µ*_*S*_ exert a strong influence on the dynamics of *I*_*L*_. Similarly, Figure 5b indicates that *θ, γ*_*D*_, and *µ*_*S*_ significantly affect the behavior of *I*_*D*_. In Figure 5c, the parameters *β, b, λ, ω, γ*_*D*_, *µ*_*S*_, and *λ*_*S*_ again exhibit a pronounced impact on *I*_*P*_. Furthermore, Figure 5d demonstrates that *β, c, λ, γ*_*D*_, Λ_*S*_, *µ*_*S*_, and *λ*_*S*_ strongly influence *I*_*S*_ as their values vary. Collectively, these results highlight the key parameters that drive variations in the infection-related compartments of the model. Biologically, these findings suggest that parameters linked to transmission intensity, host recruitment, and recovery dynamics play a central role in shaping the prevalence of different infection stages. Controlling factors such as the sandfly biting rate, transmission probabilities, and host mortality could therefore substantially reduce the overall disease burden.

**Fig 5.**
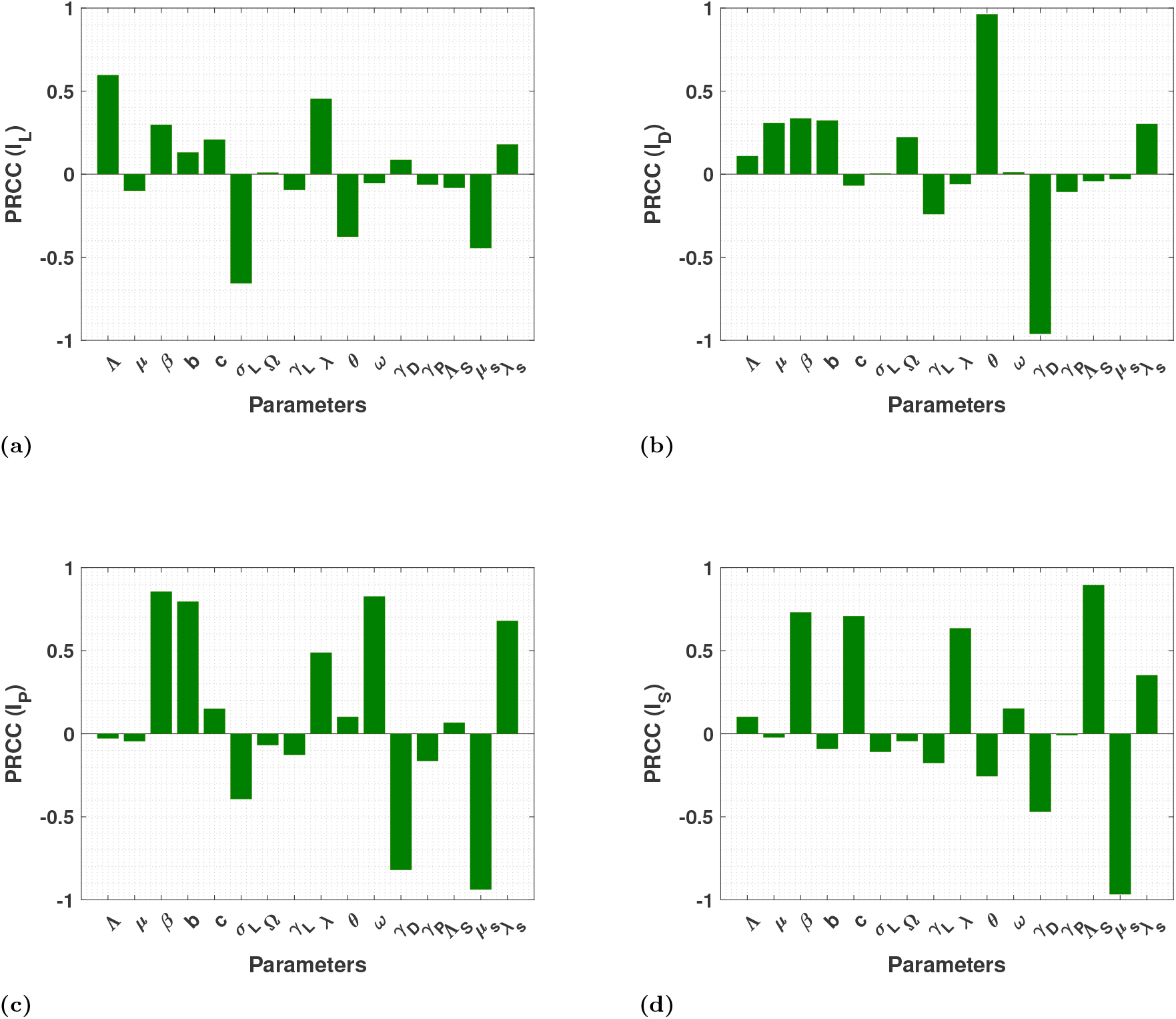
Results from the PRCC indicating the sensitivity indices of the model parameters with (a) VL only infected (*I*_*L*_), (b) VL dormant (*I*_*D*_), (c) PKDL infected (*I*_*P*_) and (d) Infected sandflies (*I*_*S*_).

## 3 Analysis of HIV only Model

### 3.1 Formulation of Equation

The HIV-only sub-model is obtained by setting all VL-related and co-infected compartments to zero, that is, 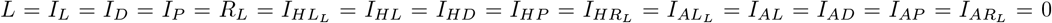 and solving the equation for the remaining compartments.

This reduction allows us to focus exclusively on HIV transmission and treatment dynamics by removing all VL-related and co-infected compartments. Biologically, the HIV-only subsystem represents a setting where VL does not circulate, enabling the assessment of HIV progression and the impact of treatment interventions in isolation.

**Flow diagram of HIV only model**

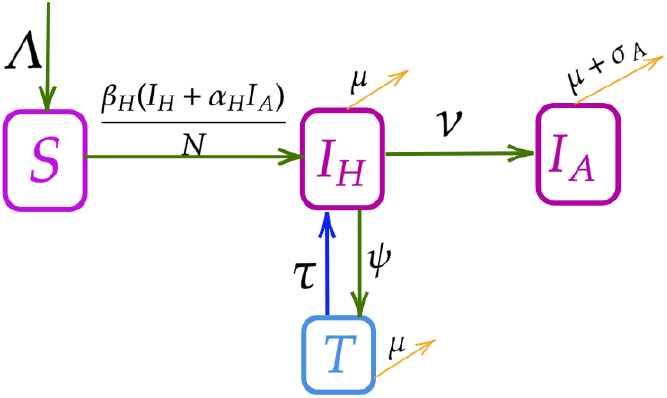

**HIV-only Model**

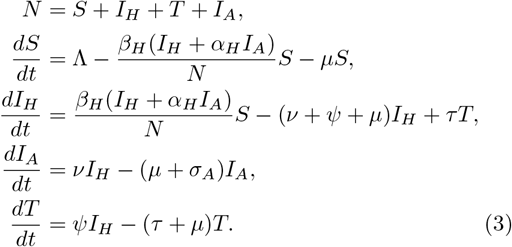

### 3.2 Positivity and the Bounded Nature of the Solution

From (3), we obtain the following

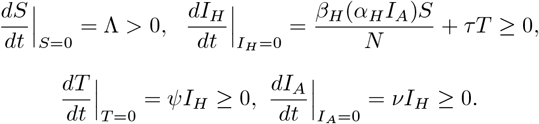

All transition rates defined in the model remain non-negative on the boundary hyperplanes. Therefore, any trajectory that originates in the interior of the non-negative bounded cone 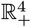 will remain within this region for all future time. This is due to the fact that the vector field points inward along each boundary surface, preventing solutions from leaving the feasible domain. Hence, all state variables of the system remain non-negative. Furthermore, in accordance with the compartmental structure of the model, the total population *N* = *S* + *I*_*H*_ + *I*_*A*_ + *T* satisfies

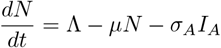

This gives lim 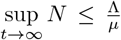. Hence every solutions *S*(*t*), *I*_*H*_ (*t*), *I*_*A*_(*t*), *T* are bounded by 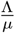. This gives us the biological feasible region of the system (3) by the below positively invariant set is given as

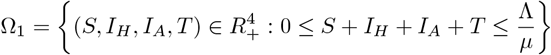

### 3.3 Existence of Equilibrium Points and Basic Reproduction Number

#### 3.3.1 Disease-free Equilibrium

For the HIV-only subsystem (3), the disease-free equilibrium (DFE) corresponds to the state in which no individuals are infected with HIV and the entire population remains susceptible. Biologically, the DFE represents a scenario where HIV transmission is absent and the infection cannot persist in the community. The disease-free equilibrium for the HIV-only model is therefore given by

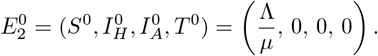

#### 3.3.2 The Basic Reproduction Number

The basic reproduction number (*R*_0*H*_) for the human-to-human transmission subsystem is derived using the next generation matrix approach [28, 29, 30]. In this formulation, the matrices *F*_2_ and *D*_2_ represent the new infection and transition terms, respectively, associated with the infected compartments *I*_*H*_, *I*_*A*_, and *T*.

The detailed forms of *F*_2_, *D*_2_, and their Jacobians *F*_2_ and *V*_2_ are provided in Supporting document. The next generation matrix is defined as 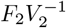, and its dominant eigenvalue yields the basic reproduction number *R*_0*H*_.

Hence, the basic reproduction number for the human infection subsystem is obtained as

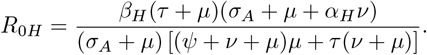

Biologically, *R*_0*H*_ quantifies the average number of secondary HIV infections produced by a single infected individual in an entirely susceptible population. HIV can invade and persist only when *R*_0*H*_ > 1, whereas *R*_0*H*_ < 1 implies that the infection will eventually die out.

##### Theorem 3.

*When the disease-free equilibrium* 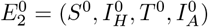 *is locally asymptotically stable if R*_0*H*_ < 1, *under some restriction on the parameters otherwise it is unstable*.

*Proof*. Proof of the theorem is available in supporting information file.

### 3.4 Existence of Endemic Equilibrium Points

This is the point in the system where the illness still exists. Consequently 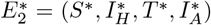 is the endemic equilibrium of the system (3), is obtained by equating the system (3) to zero

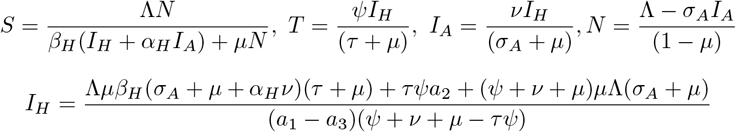

where

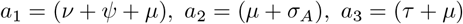

### 3.5 Bifurcation and Stability Analysis of the Endemic Equilibrium

The detailed derivations are provided in Supporting document.

According to the theorem presented in [31], since *a <* 0 and *b >* 0, the system admits a unique HIV-only endemic equilibrium point, denoted by 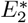, which exists whenever the corresponding reproduction number *R*_0*H*_ > 1. Therefore, we establish the following theorem.

#### Theorem 4.

*The unique endemic equilibrium* (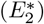) *of the system* (2) *is locally asymptotically stable when R*_0*H*_ > 1.

### 3.6 Numerical Analysis

This section presents the numerical simulations and analysis of the HIV-only model. Model validation is performed by fitting the simulated results to the yearly HIV incidence data from Bihar. Parameter estimation is carried out to obtain the best-fit values, followed by sensitivity and PRCC analyses to identify the most influential parameters affecting disease dynamics.

#### 3.6.1 Data Fitting

Data fitting for the HIV-only model is carried out using yearly HIV incidence data from Bihar, India. All model parameters are estimated, except for the constant recruitment rate and the natural death rate, which are obtained from demographic statistics. The yearly cumulative HIV case data for Bihar, covering the period from 2010 to 2023, are obtained from the official factsheets of the National AIDS Control Organization (NACO) https://naco.gov.in/.

The initial conditions for the HIV-only model compartments were set as *I*_*h*_(0) = 5183.974699, *I*_*a*_(0) = 4666.946449, and *T* (0) = 4146.675894, corresponding to the estimated values for the base year 2010. Parameter estimation is performed using the Maximum Likelihood Estimation (MLE) [32] method implemented in Python to achieve the best possible agreement between model predictions and observed data. Variables such as *I*_*A*_ and *T* are also estimated during the fitting process, since direct observational data for these compartments are not available. The estimated parameter values are summarized in Table 5 and data fitting graph is provided in Figure 6.

**Table 5.** Parameter values for HIV dynamics.

| Parameter | Description | Values | Reference |
| --- | --- | --- | --- |
| $\beta_H$ | HIV transmission rate | 0.0605 | Fitted |
| $\alpha_H$ | Modification parameter | 0.0270 | |
| $\tau$ | Rate at which individuals discontinue treatment and return to $I_H$ | 0.03 | |
| $\psi$ | Rate of ART treatment initiation for HIV-infected individuals | 0.0381 | |
| $\nu$ | Rate at which HIV infection progresses to AIDS | 0.0557 | |
| $\sigma_A$ | AIDS-related death rate | 0.0801 | |
| $R_{0H}$ | Basic Reproduction number for HIV only model | 1.0708 | |

**Fig 6.**
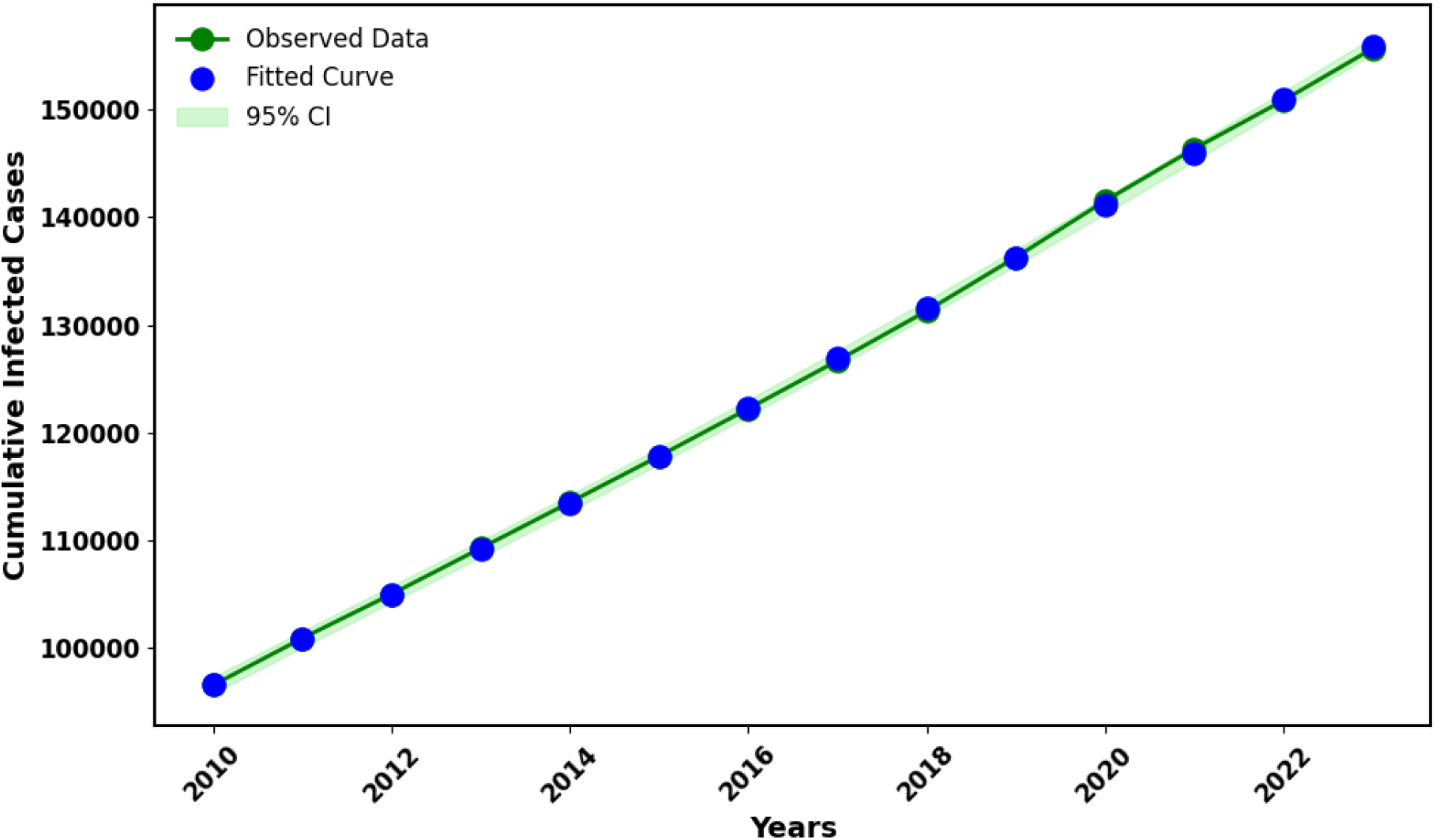
Data fitting plot for the HIV-only model. The green line represents the model fit, while the blue dots indicate the observed cumulative yearly HIV cases of Bihar.

### 3.7 Sensitivity Analysis

Sensitivity analysis is a crucial technique for evaluating the validity of an epidemiological study’s conclusions. Changes to major parameters must be made within a certain range in order to ascertain their impact on the fundamental reproduction number *R*_0*H*_). The HIV model contains significant elements that affect (*R*_0*H*_), such as *β*_*H*_, *α*_*H*_, *ν*. To produce the normalized forward sensitivity index of a variable with regard to a parameter, we calculated the ratio of the relative change in the variable to the relative change in the parameter [35]. With regard to the basic reproduction number (*R*_0*H*_), Figure7 demonstrates that the parameters *β*_*H*_, *α*_*H*_ have positive sensitivity indices, indicating that they play a significant role in increasing (*R*_0*H*_). Conversely, the other parameters have negative sensitivity indices, meaning they contribute to a decrease in (*R*_0*H*_). Biologically, this indicates that higher HIV transmission and progression rates enhance the potential for disease spread, thereby increasing the basic reproduction number. In contrast, parameters associated with recovery, treatment, or natural mortality reduce (*R*_0*H*_), highlighting their importance in controlling HIV transmission within the population.

**Fig 7.**
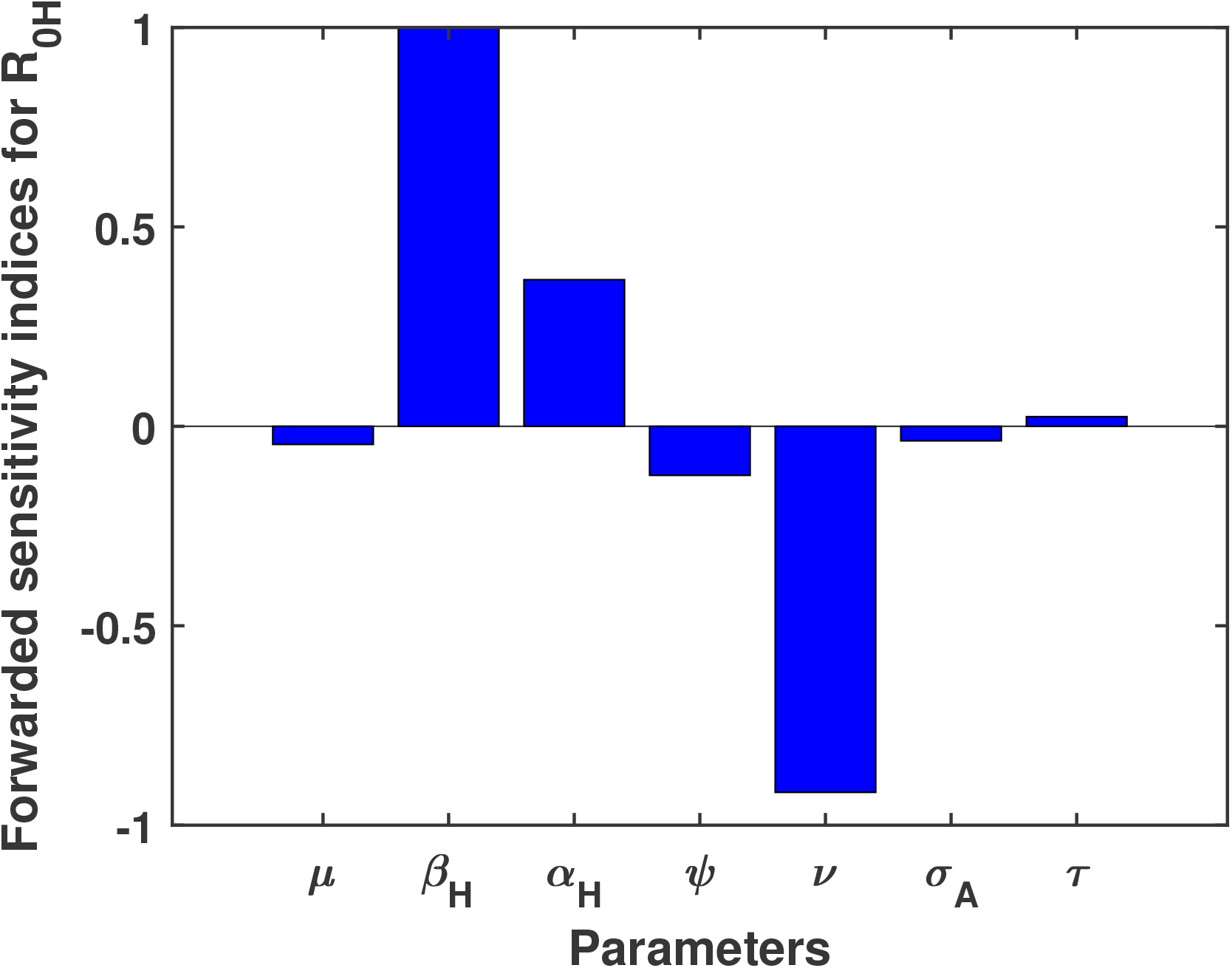
Forward sensitivity analysis of the HIV-only model with respect to the basic reproduction number (*R*_0*H*_) using the parameter values in Table5

**Fig 8.**
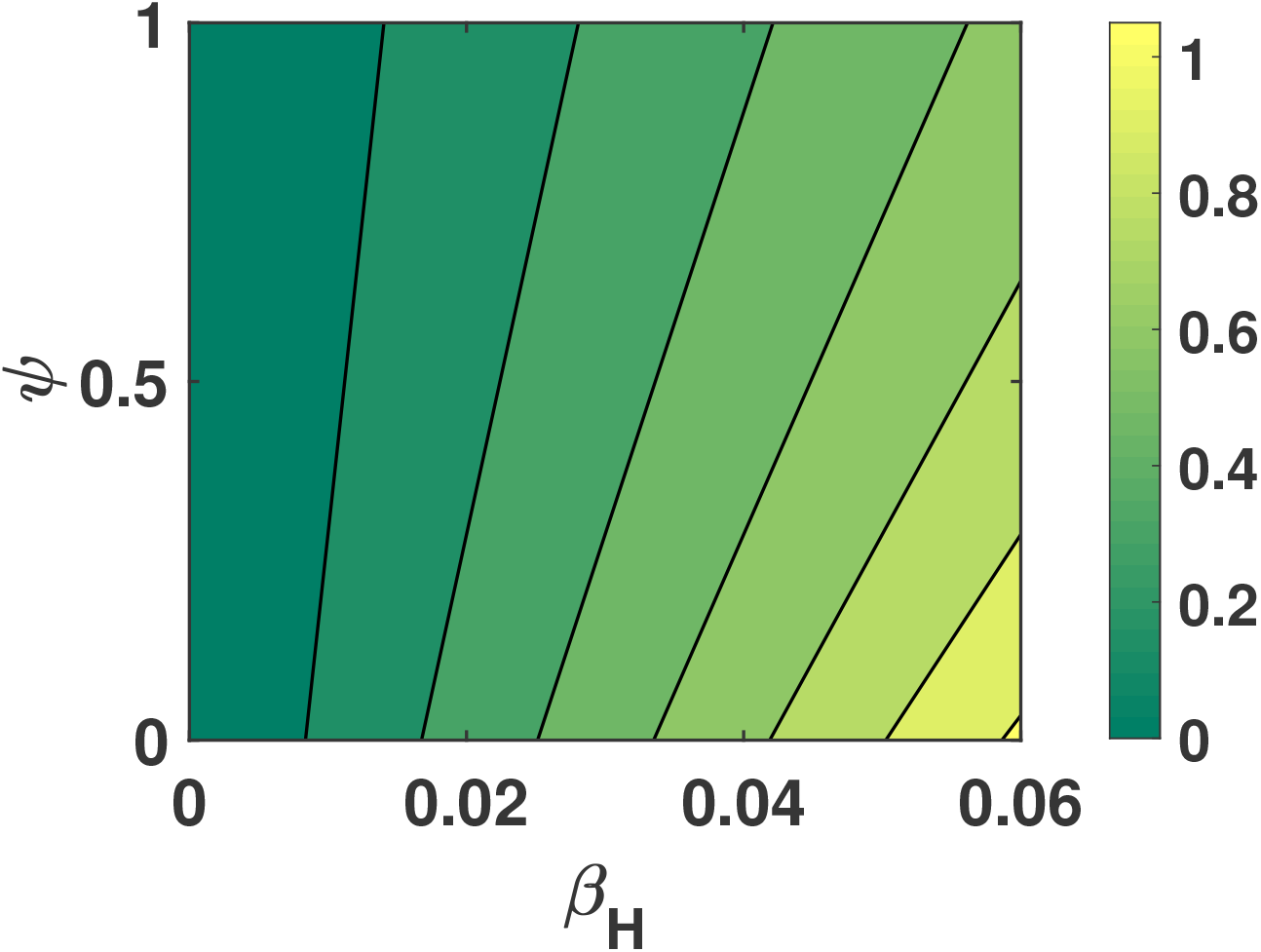
The contour plot of *β*_*H*_ and *ψ* illustrates how variations in these parameters influence the value of *R*_0*H*_, while all other parameters remain fixed, as shown in Table 5.

For system (3), we perform a global sensitivity analysis using the technique described in [36, 37]. The response function was HIV infected, AIDS infected, *I*_*H*_, *I*_*A*_ infected. We used the following parameters as inputs:Λ, *µ, β*_*H*_, *α*_*H*_, *τ, ψ, ν, σ*_*A*_. We used partial rank correlation coefficients (PRCCs) and Latin Hypercube Sampling (LHS) as our two statistical approaches. While the latter links the input parameters and response function, assigning values between −1 and 1, the former allows variations of several parameters together in a systematic manner. The PRCC sign indicates the kind of correlations that exist between the response function and the input parameters, while the values of the correlation indicate their strength. As a prerequisite for computing PRCCs, nonlinear and monotone relationships were identified between the input parameters and the symptomatic infectious population. 1000 simulations are conducted for each LHS, taking into account a uniform distribution for each input parameter. 25% of the nominal values of the parameters are expected to be exceeded.

The Partial Rank Correlation Coefficient (PRCC) values for each parameter in relation to the corresponding response function. When evaluating a parameter’s role in model prediction and its accuracy, it is important to consider both the magnitude and direction of the PRCC values. PRCC values [37] are significant when they are greater than 0.5 (indicating a strong positive influence) or less than −0.5 (indicating a strong negative influence). Figure 9a illustrates that the parameter Λ, *β*_*H*_, *ν* has a strong influence on *I*_*H*_. Similarly, Figure 9b shows that the parameter *β, ν, σ*_*A*_ has a significant impact on *I*_*A*_ with changes in its values. These figures highlight the key parameters that notably affect the outcome measures in the model

**Fig 9.**
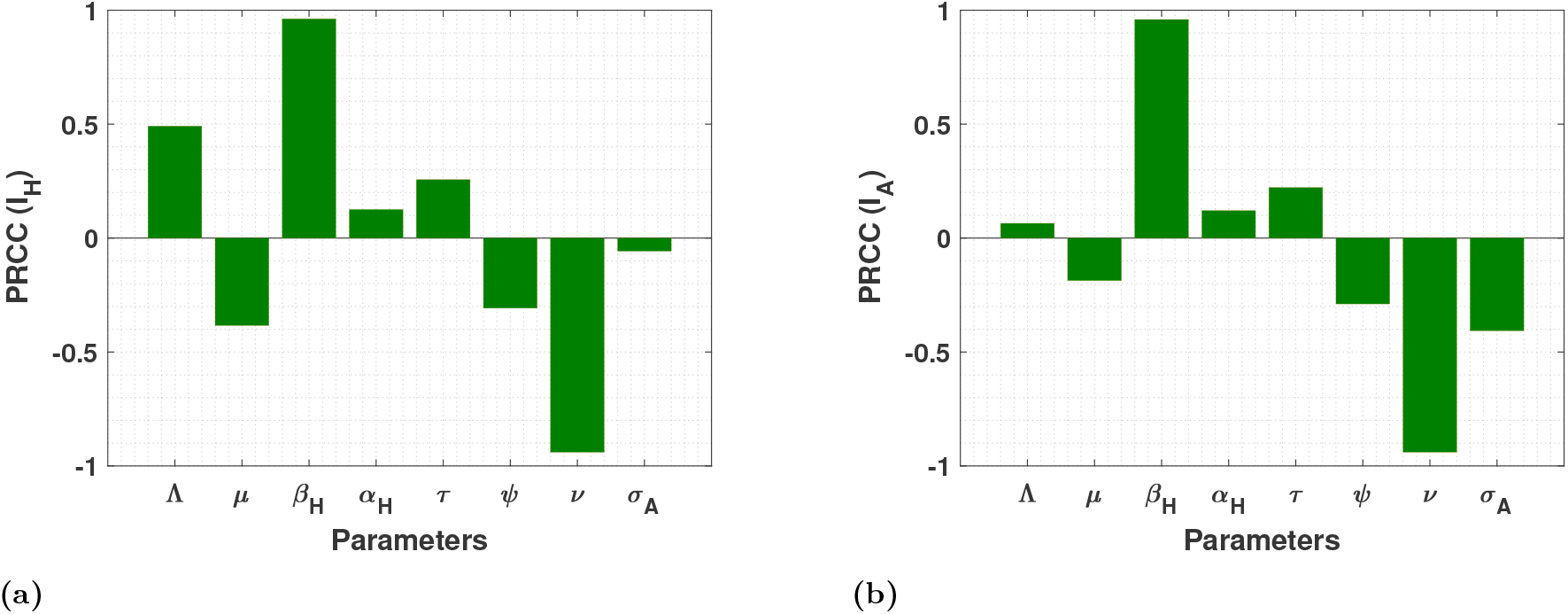
Results from the PRCC analysis showing the sensitivity indices of the model parameters for (a) HIV-infected individuals (*I*_*H*_) and (b) AIDS-infected individuals (*I*_*A*_).

## 4 Complete VL–HIV Co-infection Model

This section introduces the full visceral leishmaniasis–HIV (VL–HIV) co-infection model, which integrates the dynamics of both the human (host) and sandfly (vector) populations. The formulation captures the interaction between VL transmission through the sandfly vector and HIV transmission within the human population, while also accounting for the progression of individuals into co-infected states. By linking single-infection compartments with co-infected classes, the model provides a comprehensive framework for analyzing transmission pathways, disease progression, and the epidemiological feedback between VL and HIV.

### 4.1 Invariant Region and Boundedness of Solutions

From Equation (1), we acquire the following.

On the boundary planes, every rate listed above is non-negative. Consequently, if we begin in the non-negative bounded cone’s interior 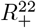, We will remain within this cone because the direction of the vector field is inward on all bounding planes. Thus, the solutions of our formulated model will be non-negative, is guaranteed. Furthermore, from the compartmental model, we conclude that the Entire population is 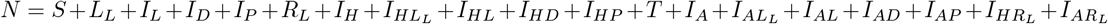 and *N*_*s*_ = *S*_*S*_ + *E*_*S*_ + *I*_*S*_ satisfies

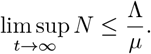

And

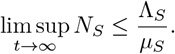

Every solutions *S*, 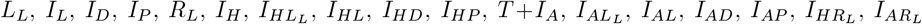 are bounded above by 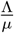, and *S* (*t*), *E*_*S*_, *I*_*S*_ (*t*) are bounded by 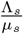 this gives feasible regions of the system (1) by the below positively invariant set;

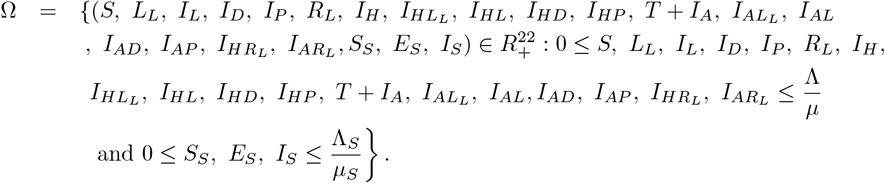

### 4.2 Existence of Equilibrium Points and the Basic Reproduction Number of Complete Co-infection Model

#### 4.2.1 Disease-free Equilibrium

For the complete VL–HIV co-infection system (1), the disease-free equilibrium (DFE) corresponds to the state in which neither VL nor HIV is present in the human population, and the sandfly vector population contains no infected individuals. Biologically, the DFE represents a scenario where both infections fail to persist in the community, and all individuals in both the host and vector populations remain susceptible.

For our formulated model, we have disease-free equilibrium as

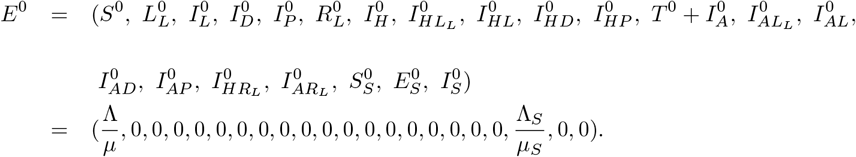

#### 4.2.2 The Basic Reproduction Number

The basic reproduction number of complete co-infection model is given by *R*_0_ = max {*R*_0*L*_, *R*_0*H*_} by theorem in[30] and using previous section we know that

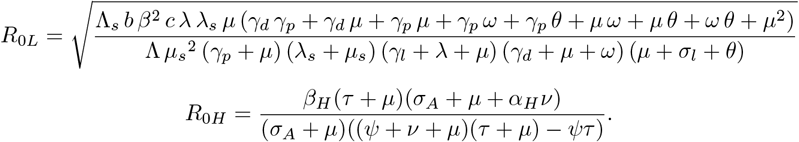

##### Theorem 5.

*The disease-free Equilibrium given by R*_0_ *is locally asymptotically stable when R*_0_ < 1 *and is unstable otherwise*.

*Proof*. Proof of theorem is available in supporting information file.

### 4.3 Numerical Analysis

This section presents the outcomes of multiple simulations, including model solutions, data fitting, parameter estimations, and disease prevalence. A detailed discussion of the findings is provided, along with model calibration for the Bihar visceral leishmaniasis and HIV co-infection data. 41

#### 4.3.1 Data Fitting

Data fitting with parameter values helps in identifying key parameters essential for understanding the dynamics of VL-HIV co-infection. In this study, model fitting and optimal parameter estimation were performed using co-infected case data from Bihar, India. The initial conditions for the model compartments were set as follows: *LI*_*h*_(0) = 480.770, *I*_*dh*_(0) = 46.314, *I*_*ph*_(0) = 5.841, *LI*_*a*_(0) = 42.760, *I*_*la*_(0) = 121.432, *I*_*da*_(0) = 3.699, *I*_*pa*_(0) = 3.204, *R*_*lIh*_(0) = 4.071, and *R*_*lIa*_(0) = 4.096. Through model calibration, four key parameters *ξ*_1_, *ξ*_2_, *α*_*S*_, and *ω*_*S*_ were estimated using the Maximum Likelihood Estimation (MLE) method. The estimated values of these parameters are summarized in Table 6.

**Table 6.** Parameter values for VL HIV co-infection dynamics.

| Parameter | Description | Value | Source |
| --- | --- | --- | --- |
| $\alpha_s$ | Modification parameter | 5.2205 | Fitted |
| $\omega_s$ | Modification parameter | 4.2958 | |
| $\xi_1$ | Relapse rate of VL infection from $I_{HD}$ | 0.027 | |
| $\xi_2$ | Relapse rate of VL infection from $I_{AD}$ | 0.6517 | |

Additionally, modification parameter values were chosen based on the conditions specified in Table 3, ensuring consistency with observed disease dynamics. The numerically simulated model was implemented in Python, and the fitted curve illustrates how well the observed data align with theoretical predictions, as shown in Figure 10. This figure highlights the effectiveness of the estimated parameters in capturing the transmission dynamics of VL-HIV co-infection.

**Fig 10.**
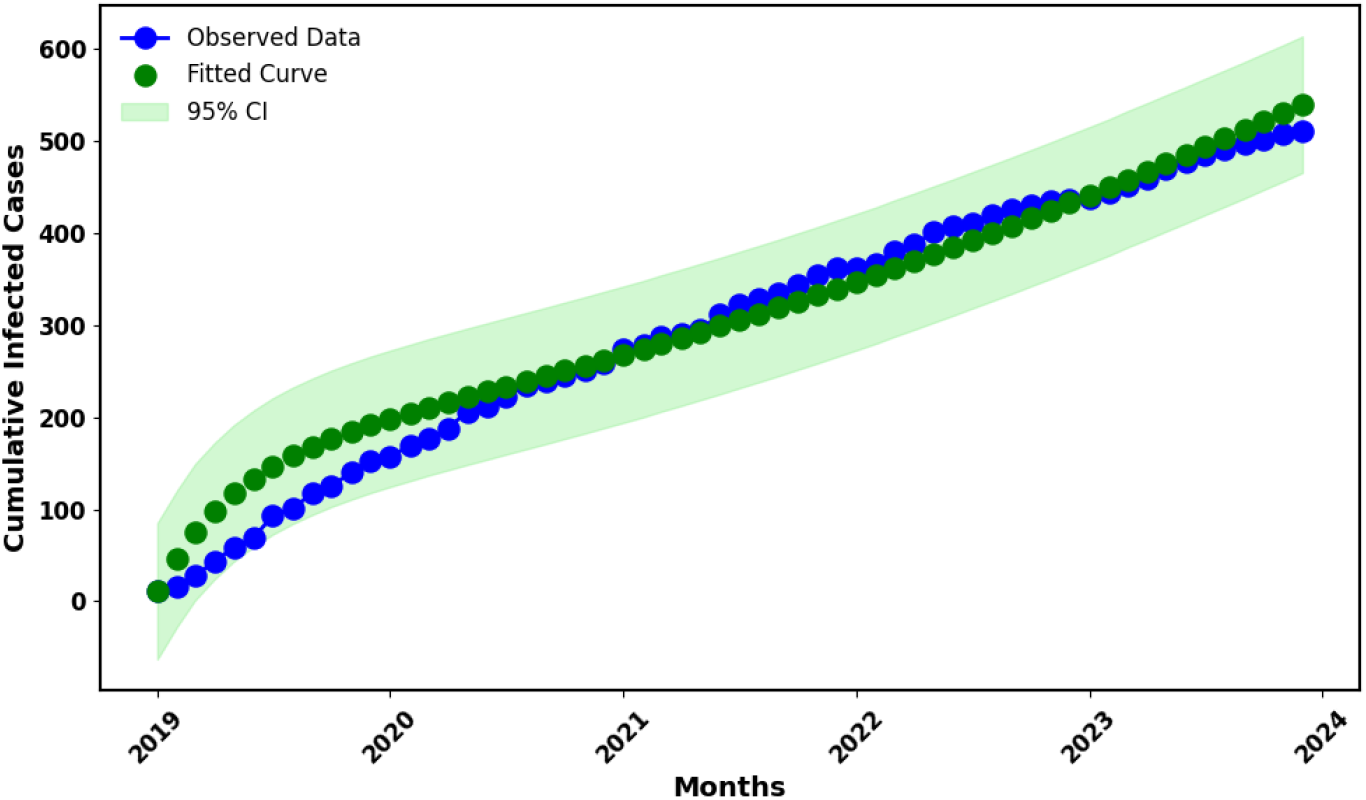
Data fitting plot for the co-infection model. The green dotted line represents the model fit, while the blue dots indicate the observed monthly infected VL HIV co-infected cases of Bihar.

Modification parameters:

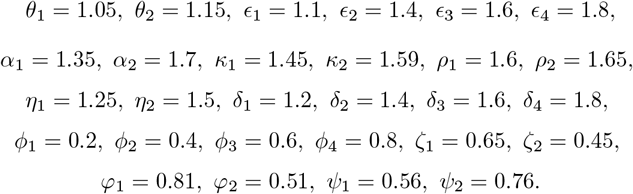

### 4.4 VL Resurgence Driven by HIV Co-infection

When the biting rate decreases to *β* = 1.9384, the basic reproduction number for the VL-only model (*R*_0*L*_) is reduced to *R*_0*L*_ = 0.5433. Figure 11 illustrates the dynamics of the infected population in the VL-only model; when *R*_0*L*_ < 1, the red dotted line tends toward zero, indicating that the VL infection eventually dies out.

**Fig 11.**
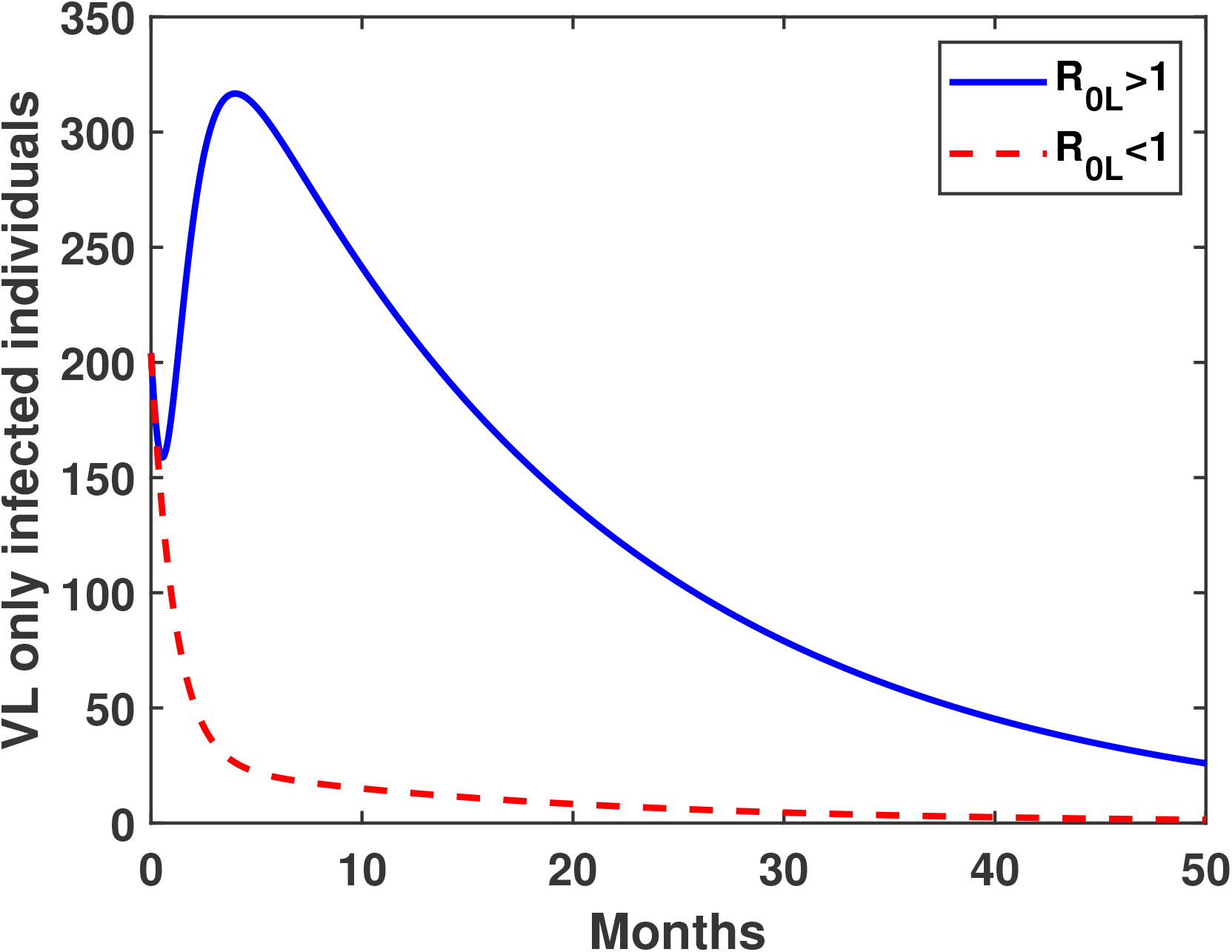
Time evolution of VL-only infected individuals for two different sandfly biting rates, illustrating the cases *R*_0*L*_ > 1 and *R*_0*L*_ < 1.

Figure 12a shows the number of VL-infected individuals in the full co-infection model. Even when the VL reproduction number is below the epidemic threshold (*R*_0*L*_ < 1), VL continues to persist in the population when HIV is present (*R*_0*H*_ > 1). This indicates that HIV infection promotes the resurgence and continued circulation of VL in the populations.

**Fig 12.**
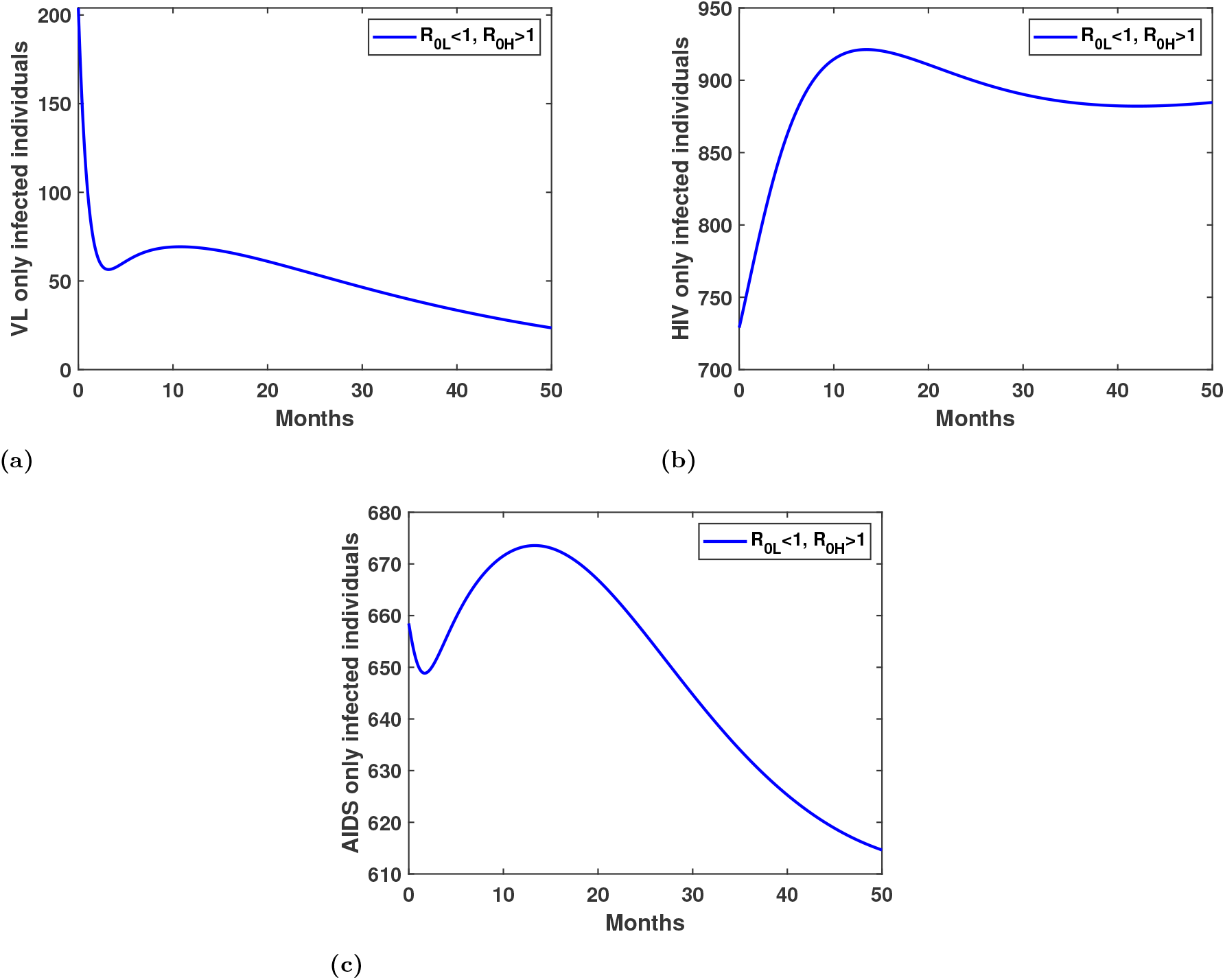
Variation of (a) VL-only infected individuals (b) HIV-only infected individuals and (c) AIDS-only infected individuals over time when *R*_0*H*_ > 1 but *R*_0*L*_ < 1.

Similarly, Figures 12b and 12c illustrate the dynamics of HIV-only infected individuals and AIDS-only infected individuals within the co-infection model. These plots highlight how HIV progresses in the presence of VL and demonstrate the interaction between the two diseases.

Figure 13a illustrates the dynamics of the co-infected population with VL-HIV. Even in scenarios where VL alone cannot persist in the community, co-infection continues to exist when HIV remains endemic. This highlights the role of HIV in sustaining VL among co-infected individuals.

**Fig 13.**
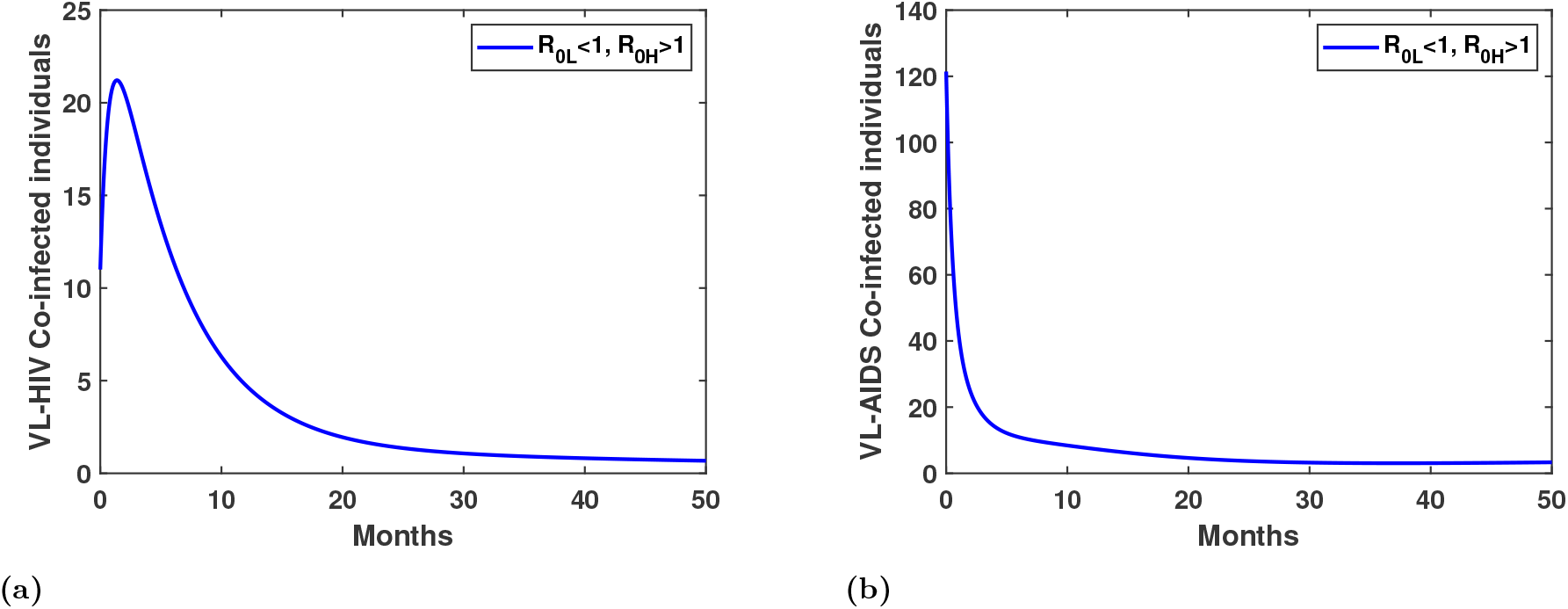
Variation of (a) VL HIV co-infected individuals (b) VL AIDS co-infected individuals individuals over time when *R*_0*H*_ > 1 but *R*_0*L*_ < 1.

Similarly, Figure 13b shows the dynamics of individuals co-infected with VL and AIDS. These results demonstrate that all co-infection compartments persist, even when the standalone VL infection would otherwise die out, underscoring the strong influence of HIV on VL persistence.

### 4.5 Impact of Treatment Rate on Disease Prevalence

HIV treatment plays a pivotal role not only in reducing HIV transmission but also in mitigating the burden of VL–HIV co-infection. As shown earlier, HIV infection contributes to the resurgence of VL by increasing parasite persistence and relapse among co-infected individuals. Increasing the HIV treatment rate therefore weakens this effect by improving immune function and reducing the likelihood of VL reactivation.

Figure 14 demonstrates that as the treatment rate rises, the number of VL–HIV co-infected individuals consistently decreases. This highlights the indirect yet significant impact of HIV treatment in controlling VL transmission in co-infected populations.

**Fig 14.**
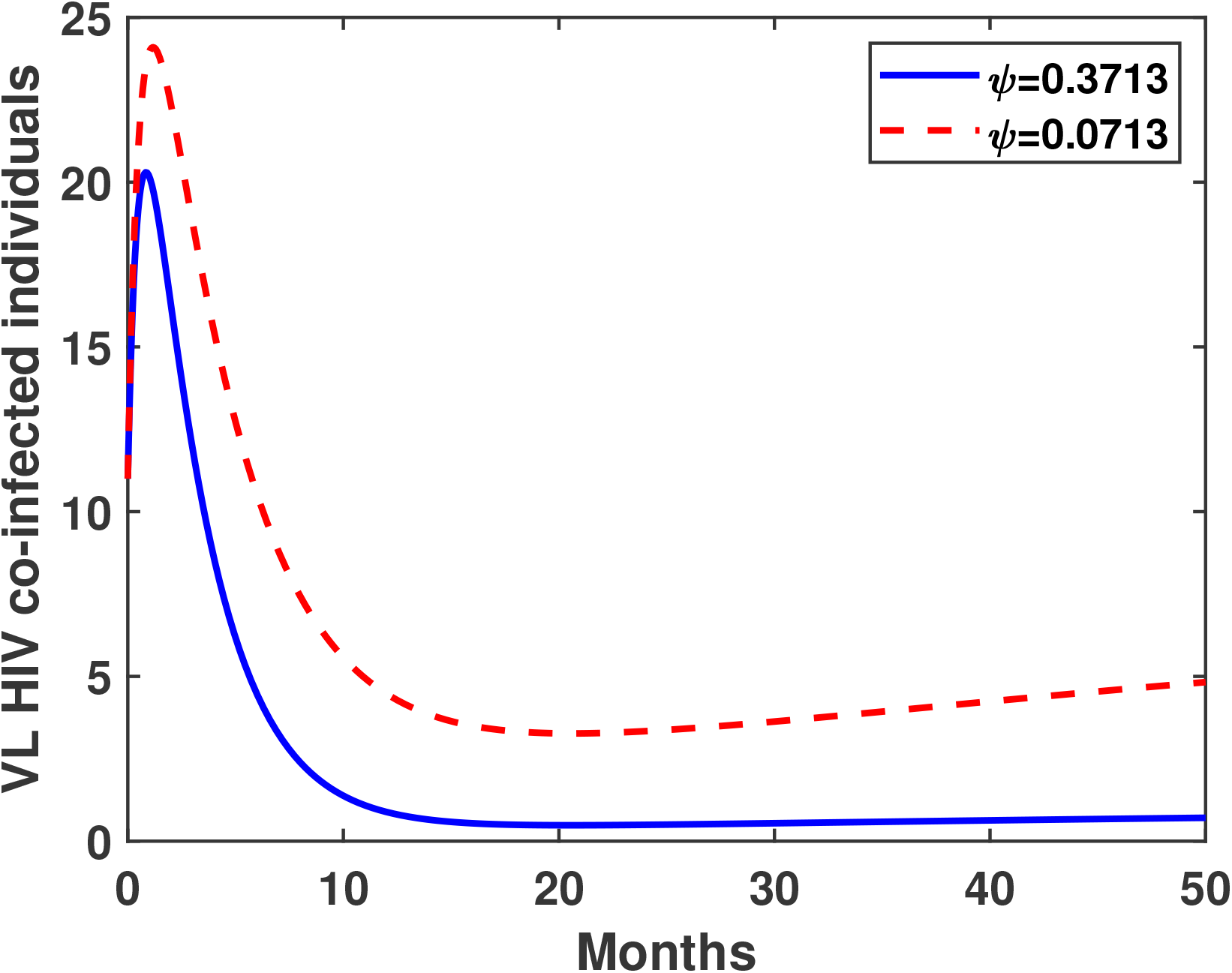
Variation of VL HIV co-infected individuals with time for different treatment rates.

### 4.6 Sensitivity Analysis

For system (1), we performed a global sensitivity analysis using the technique described in [38, 36]. In Section 2,3, we presented PRCC analysis for both the VL-only model and the HIV-only model, assessing the impact of parameters on *I*_*L*_, *I*_*D*_, *I*_*P*_, *I*_*s*_, *I*_*H*_, *I*_*A*_. We now extend this analysis to the co-infection model. Here we are going to study the impact of parameters on co-infected compartments like (*I*_*L*_, *I*_*H*_, *I*_*A*_, *I*_*HL*_, *I*_*AL*_, *I*_*S*_), in order to understand how changes in these parameters affect the dynamics of the co-infection model. By performing a PRCC on these compartments, we can gain insight into which parameters play a significant role in driving the behavior of the system. This information will be crucial for developing strategies to control and manage co-infections of VL and HIV in the population. The response function was (*I*_*KH*_, *P*_*KH*_, *I*_*KA*_, *I*_*s*_). We used the following parameters as inputs:Λ, *µ, β, b, c, σ*_*L*_, Ω, *γ*_*L*_, *λ, θ, ω, γ*_*D*_, *γ*_*P*_, Λ_*S*_, *µ*_*S*_, *λ*_*S*_, *β*_*H*_, *α*_*H*_, *τ, ψ, ν, σ*_*A*_, *α*_*S*_, *ω*_*S*_, *ξ*_1_, *ξ*_2_. We used partial rank correlation coefficients (PRCCs) and Latin Hypercube Sampling (LHS) as our two statistical approaches. While the latter links the input parameters and response function, assigning values between −1 and 1, the former allows variations of several parameters together in a systematic manner. The PRCC sign indicates the kind of correlations that exist between the response function and the input parameters, while the values of the correlation indicate their strength. As a prerequisite for computing PRCCs, nonlinear and monotone relationships were identified between the input parameters and the symptomatic infectious population. 1000 simulations are conducted for each LHS, taking into account a uniform distribution for each input parameter. 25% of the nominal values of the parameters are expected to be exceeded.

The Partial Rank Correlation Coefficient (PRCC) values indicate the influence of each parameter on the corresponding response function. When assessing a parameter’s role in model predictions, both the magnitude and direction of PRCC values are essential. A value greater than 0.5 suggests a strong positive influence, while a value less than −0.5 indicates a strong negative effect [37].

Figure 15a highlights that parameters *β, b, c, θ*, Λ_*s*_, *µ*_*S*_, *λ*_*S*_, *β*_*H*_ and *ν* strongly influence *I*_*L*_. Similarly, Figure 15b shows that *β, b, c, ν, λ*, Λ_*S*_, *µ*_*S*_, *β*_*H*_ significantly impacts *I*_*H*_, while Figure 15c identifies *α*_*H*_, and *ν* as key factors affecting *I*_*A*_. The infected sandfly population (*I*_*S*_) is notably influenced by *β, b*, Λ_*S*_, *µ*_*S*_, as illustrated in Figure 15d.

**Fig 15.**
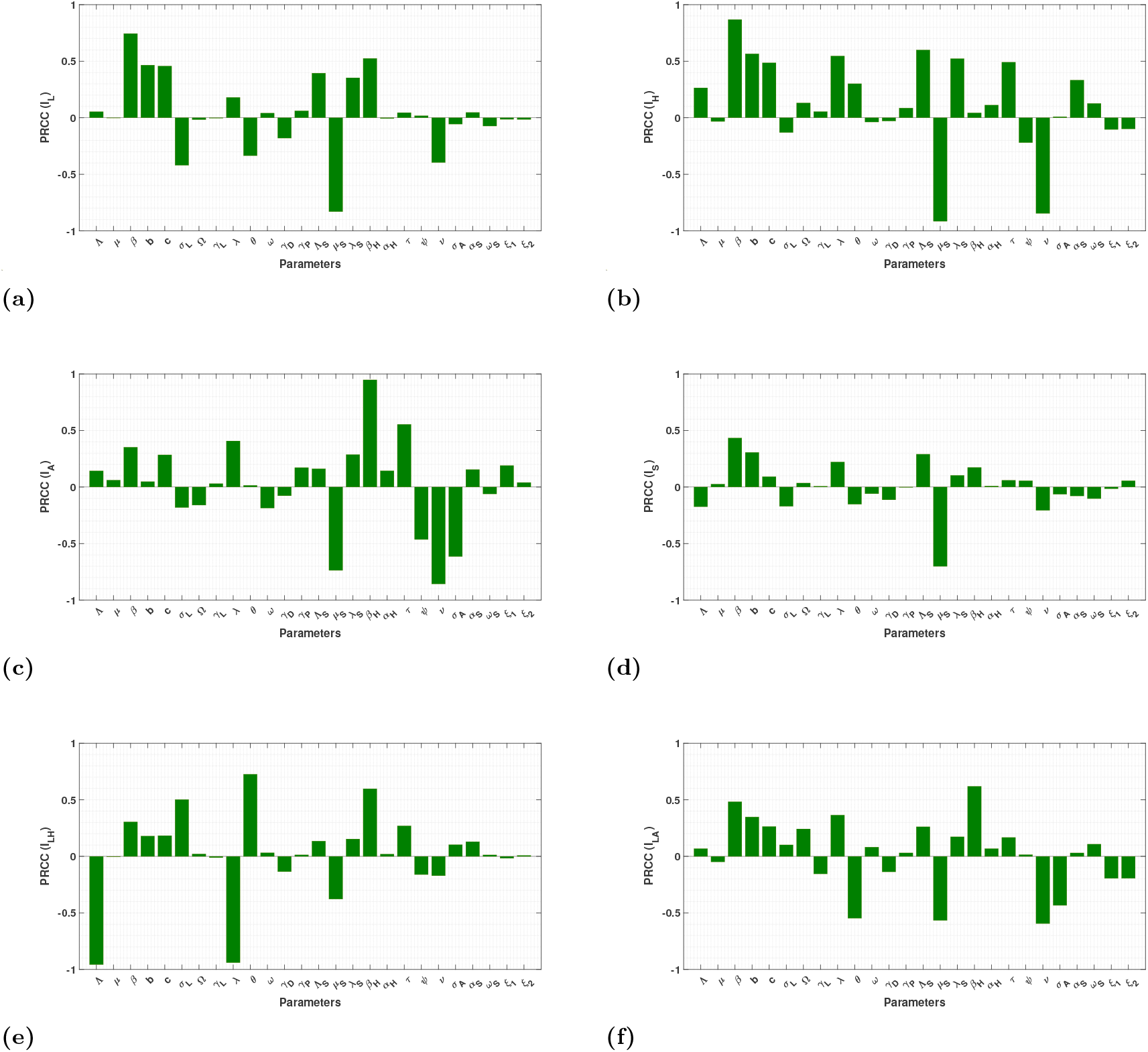
Results from the PRCC analysis showing the sensitivity indices of the model parameters for (a) VL infected individuals (*I*_*L*_) (b) HIV infected (*I*_*H*_) (c) AIDS-infected individuals (*I*_*A*_), (d) Infected sandflies (*I*_*S*_), (e) VL HIV co-infected (*I*_*HL*_) and (f) VL AIDS co-infected (*I*_*AL*_).

For the co-infected compartments, Figure 15e demonstrates that Λ, *σ*_*L*_, *λ, θ, β*_*H*_, play a significant role in *I*_*HL*_. Similarly, Figure 15f shows that *β, b, c, θ, µ*_*S*_, strongly impact *I*_*AL*_.

The PRCC analysis provides valuable insights into the key parameters driving the dynamics of the co-infection model. Our findings highlight the significant influence of parameters such as Λ, *σ*_*L*_, Ω, *θ, β*_*H*_, and *α*_*H*_ across multiple infected compartments, emphasizing their role in disease progression. Additionally, factors like *β, c*, Λ_*S*_, *µ*_*S*_, and *λ*_*S*_ critically impact the infected sandfly population (*I*_*S*_), reinforcing their importance in vector control strategies.

By identifying these influential parameters, our analysis offers a deeper understanding of the factors governing VL-HIV co-infection dynamics. This knowledge is essential for developing effective intervention strategies, optimizing resource allocation, and improving disease management efforts in affected populations.

## 5 Optimal Control

In order to manage HIV and visceral leishmaniasis co-infection, optimal control analysis is essential. We can successfully lower transmission rates and enhance patient outcomes by identifying critical elements that contribute to the spread of these diseases and putting focused treatments into place. Moreover, the implementation of optimal control strategies facilitates the more efficient allocation of resources by healthcare professionals, resulting in a more economical and long-lasting method of controlling co-infections. Here we extend our model (1) by introducing three optimal control parameters, namely *u*_1_, *u*_2_, *u*_3_

1. *u*_1_: This control variable is designed to reduce the transmission rate of visceral leishmaniasis. By implementing *u*_1_, we aim to decrease the number of new infections within the population. This can be achieved through a combination of interventions, including enhanced vector control measures such as the distribution of insecticide-treated nets and the use of protective body lotions to prevent sandfly bites. Additionally, improvements in treatment and prevention strategies, as well as public awareness campaigns, contribute to reducing the transmission coefficient associated with VL. The ultimate goal of *u*_1_ is to lower the overall incidence and spread of VL within the community, thereby controlling the disease more effectively.
2. *u*_2_: This control variable aims to reduce the transmission rate of HIV. The implementation of *u*_2_ involves a range of strategies to minimize new HIV infections. Key interventions include public education on safe sex practices, promoting condom use, and reducing the risk associated with the use of shared needles. Efforts to provide clean needles and needle exchange programs are crucial in preventing transmission among individuals who inject drugs. The goal of *u*_2_ is to lower the HIV transmission coefficient, thereby decreasing the incidence of new infections and improving public health outcomes related to HIV.
3. *u*_3_: This control variable is designed to increase the mortality rate of sandflies to reduce the incidence of visceral leishmaniasis (VL). Specifically, the natural death rate of sandflies, *µ*, is modified to *µ* + *du*_3_(*t*), where *d* represents the effectiveness of the control intervention. Controlling the sandfly population is a key component of *u*_3_ implementation, as it helps curb the spread of VL. Interventions such as insecticides, biological control agents, and environmental management strategies play a crucial role in minimizing sandfly breeding sites. By effectively increasing the sandfly death rate through *u*_3_, the transmission of VL is reduced, ultimately lowering the overall incidence and improving public health outcomes related to the disease.

These three control functions are bounded and Lebesgue integrable over the interval [0, *t*_*f*_], where *t*_*f*_ represents the predefined time period during which the controls are applied. It is assumed that all control functions *u*_1_, *u*_2_, *u*_3_ range between 0 and 1. A value of 0 indicates that no effort is applied to the corresponding control, while a value of 1 signifies maximum effort. Based on this understanding, we incorporate these five controls into the model (1), resulting in the following optimal control model:

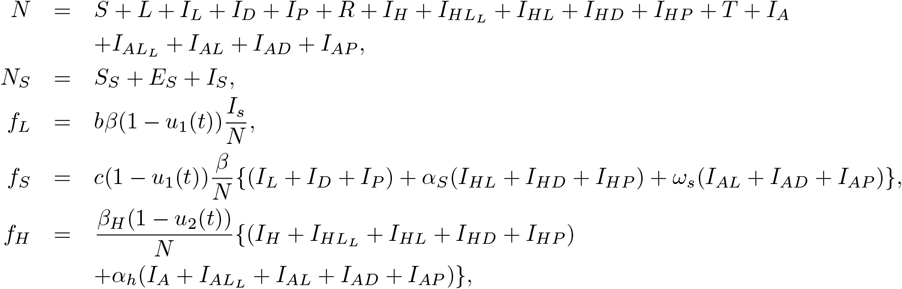

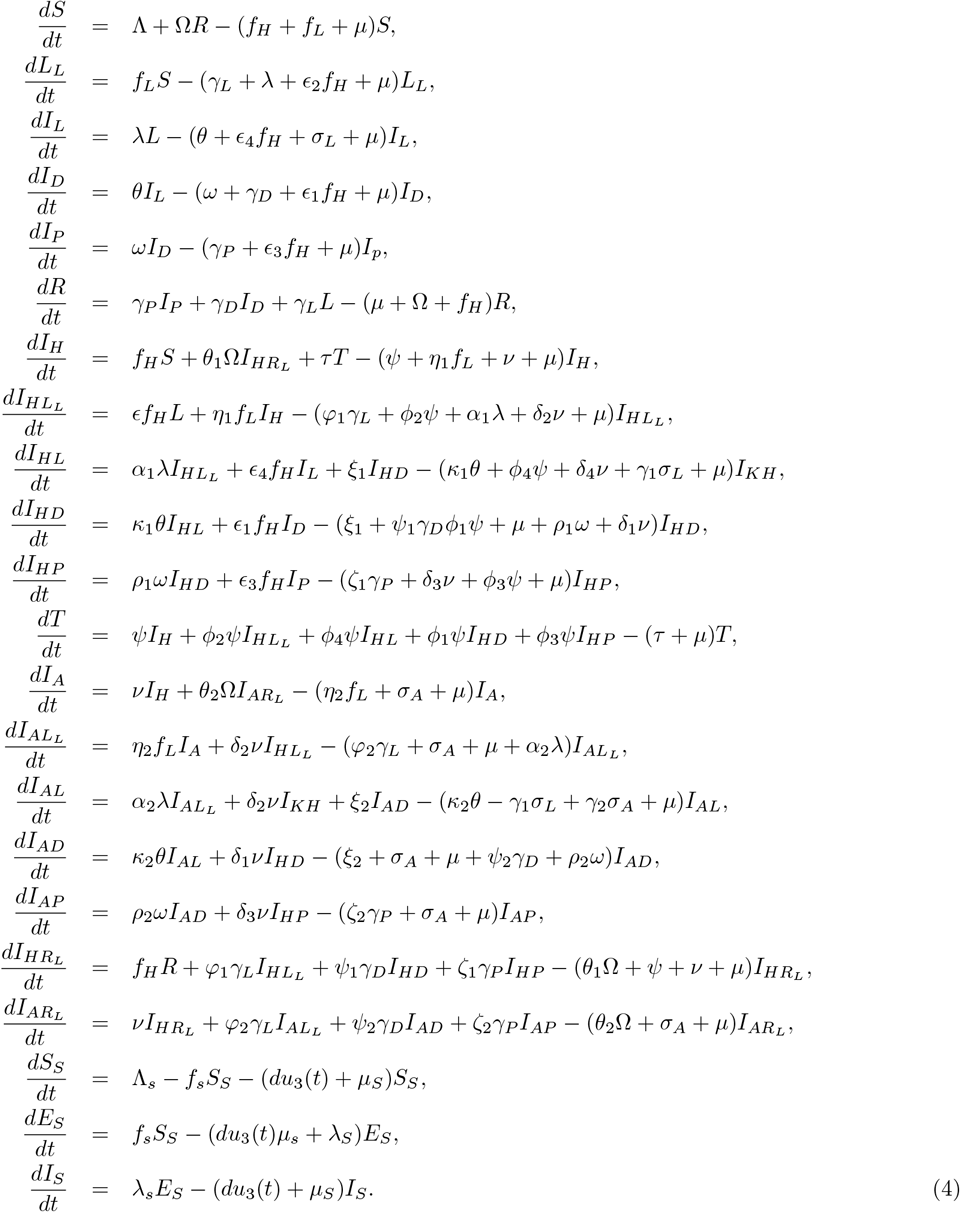

For fixed *t*_*p*_, the objective functional is provided by

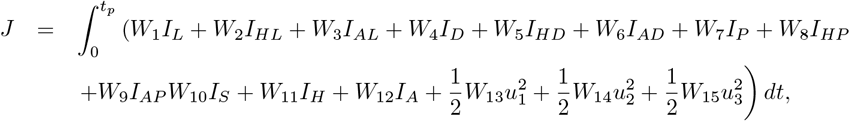

where *W*_1_, *W*_2_, *W*_3_, *W*_4_, *W*_5_, *W*_6_, *W*_7_, *W*_8_, *W*_9_, *W*_10_, *W*_11_, *W*_12_, *W*_13_, *W*_14_, *W*_15_ ≥ 0 are the weight constants.

The goal is to determine the control parameters 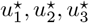 such that

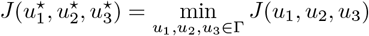

where Γ is the control set, defined as

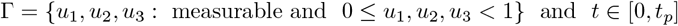

The Lagrangian of this problem is:

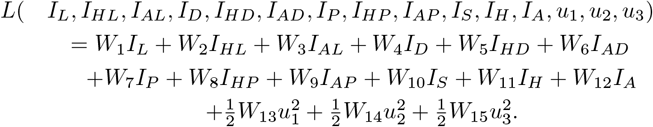

The Hamiltonian ℋ_*C*_ formed for our problem is:

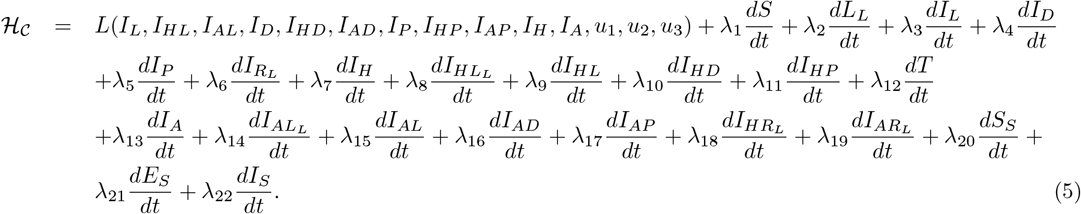

where 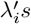 are the adjoint variables (*i* = 1 *to* 22). The corresponding different equations representing the adjoint variables are provided in supporting information file.

Let

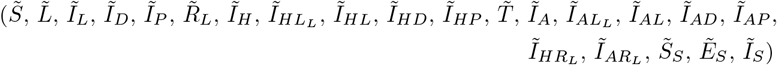

be the optimal solution corresponding to

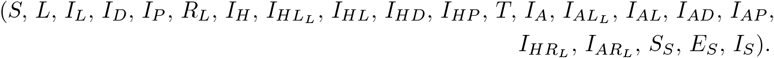

be a solution of system (4). By using [39, 40, 41], we state and prove the below theorem.

### Theorem 6.

*There exist optimal controls* 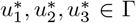, *such that* 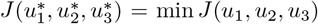 *subject to extended system of equations*(4)

*Proof*. Proof of the Theorem is available in supporting information file.

### Theorem 7.

*The optimal controls u*_1_∗, *u*_2_∗, *u*_3_∗ *which minimize J over the region* Γ *is given by:*

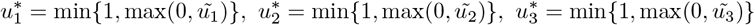

*where*

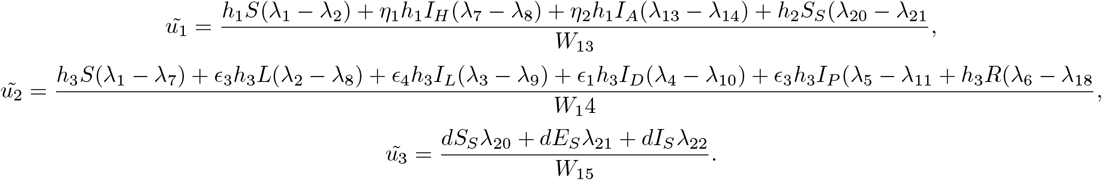

*Proof*. Proof of the Theorem is available in supporting information file.

### 5.1 Numerical Simulation

This section presents a detailed numerical analysis of optimal control using MATLAB, conducted over 60 months. The simulations are based on the parameter set provided in Table 5,4,6. As in [41], an iterative approach employing forward and backward difference approximations is used to solve the expanded system of equations (4). The state equations (4) are solved first using the forward difference approximation, and the adjoint equations are solved next using the backward difference approximation. The weights that balance off the infection compartments are represented by the positive valued constants *W*_1_, *W*_2_, *W*_3_, *W*_4_, *W*_5_, *W*_6_, *W*_7_, *W*_8_, *W*_9_, *W*_10_, *W*_11_*andW*_12_. The positive constants *W*_13_, *W*_14_, *W*_15_ represent the weight constants for optimal control *u*_*i*_’s respectively.

### 5.2 Scenario A: Single-Control Strategies

1. **Strategy I:** Only the control *u*_1_ is applied. This strategy focuses solely on reducing the transmission rate of visceral leishmaniasis (VL) through interventions such as insecticide-treated nets, protective lotions, and public awareness campaigns.
2. **Strategy II:** Only the control *u*_2_ is applied. This strategy targets the reduction of HIV transmission by promoting safe sex practices, condom use, and needle exchange programs.
3. **Strategy III:** Only the control *u*_3_ is applied. This strategy aims to increase the mortality rate of sand-flies by modifying their natural death rate to *µ* + *du*_3_(*t*) using insecticides, biological control agents, and environmental management strategies.

Figure 16 illustrates the effect of each single-control strategy on the co-infected populations *I*_*LH*_ and *I*_*LA*_. Among the individual control strategies, the implementation of *u*_1_(*t*) demonstrates the greatest reduction in the number of co-infected individuals compared to *u*_2_(*t*) and *u*_3_(*t*).

**Fig 16.**
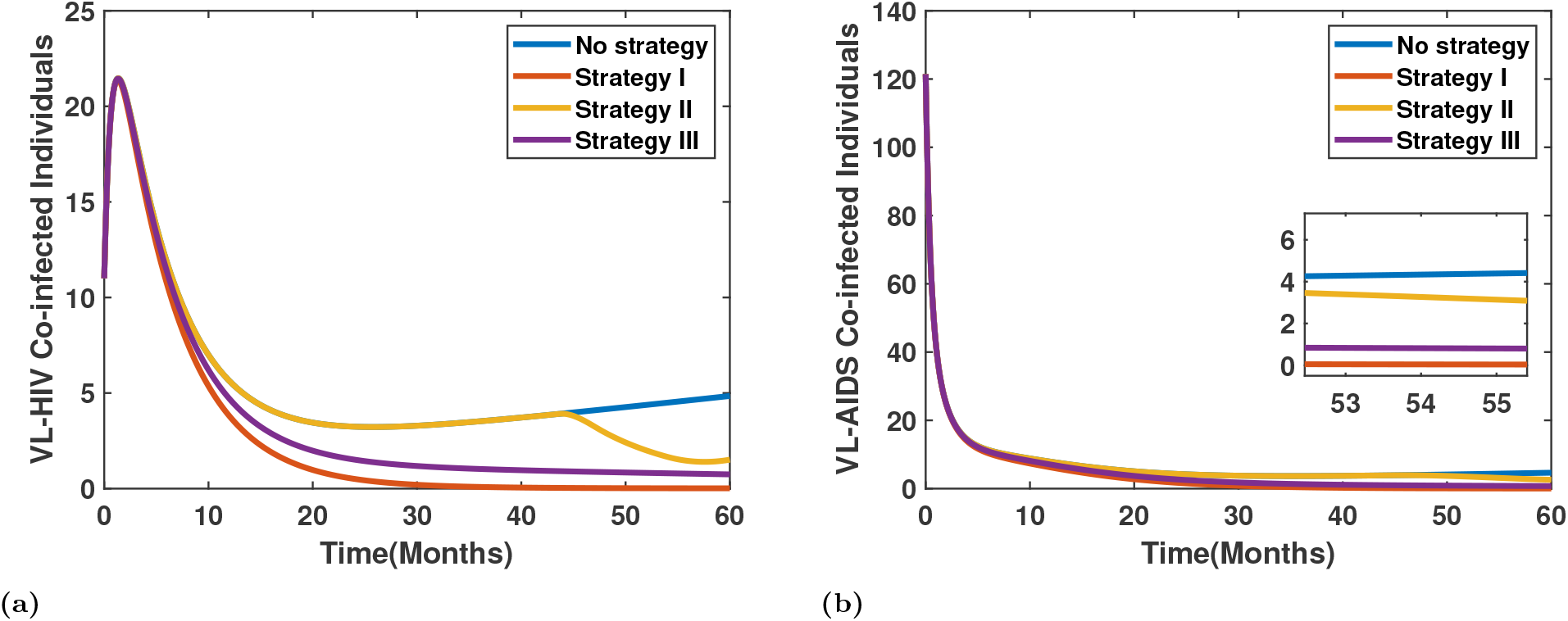
Variation in the infected population under Scenario A.

Figure 17 presents the optimal time-dependent control profiles when each control (*u*_1_, *u*_2_, *u*_3_) is applied individually. For *u*_1_(*t*) and *u*_3_(*t*), the control efforts start at their maximum intensity and gradually decrease over time, indicating the importance of early and sustained intervention in reducing infection levels. In contrast, the *u*_2_(*t*) profile remains low during the initial period and becomes active only toward the later stage of the simulation, suggesting that this intervention is most effective when applied in the advanced phase of the epidemic.

**Fig 17.**
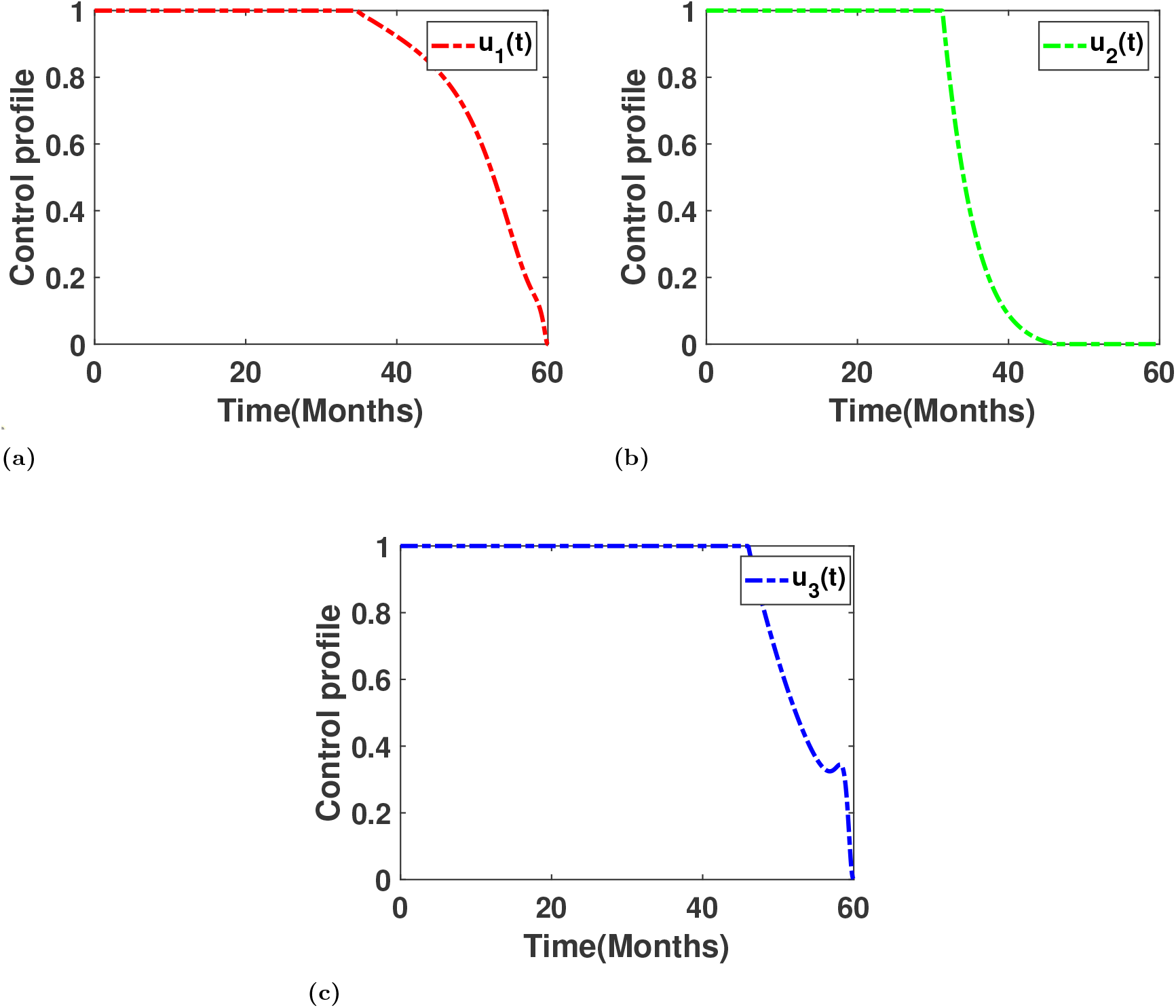
Control profile when only one control applied

### 5.3 Scenario B: Combined-Control Strategies

1. **Strategy IV:** Both controls *u*_1_ and *u*_2_ are applied. This strategy simultaneously reduces the transmission rates of visceral leishmaniasis (VL) and HIV through interventions such as insecticide-treated nets, protective lotions, public awareness campaigns, safe sex education, condom use, and needle exchange programs.
2. **Strategy V:** Both controls *u*_1_ and *u*_3_ are applied. This strategy focuses on reducing the transmission of VL while increasing the mortality rate of sandflies. It combines interventions such as vector control measures and insecticide-treated nets with sandfly population control through insecticides, biological agents, and environmental management.
3. **Strategy VI:** Both controls *u*_2_ and *u*_3_ are applied. This strategy targets the reduction of HIV transmission while simultaneously increasing the mortality rate of sandflies. It integrates HIV prevention measures, such as safe sex education and needle exchange programs, with sandfly control strategies to mitigate VL transmission.

Figure 18 illustrates the impact of each combined control strategy on the dynamics of *I*_*HL*_ and *I*_*AL*_. Among all pairwise strategies, Strategy IV (*u*_1_(*t*), *u*_2_(*t*)) demonstrates the most significant reduction in co-infection levels, highlighting its effectiveness in controlling disease transmission.

**Fig 18.**
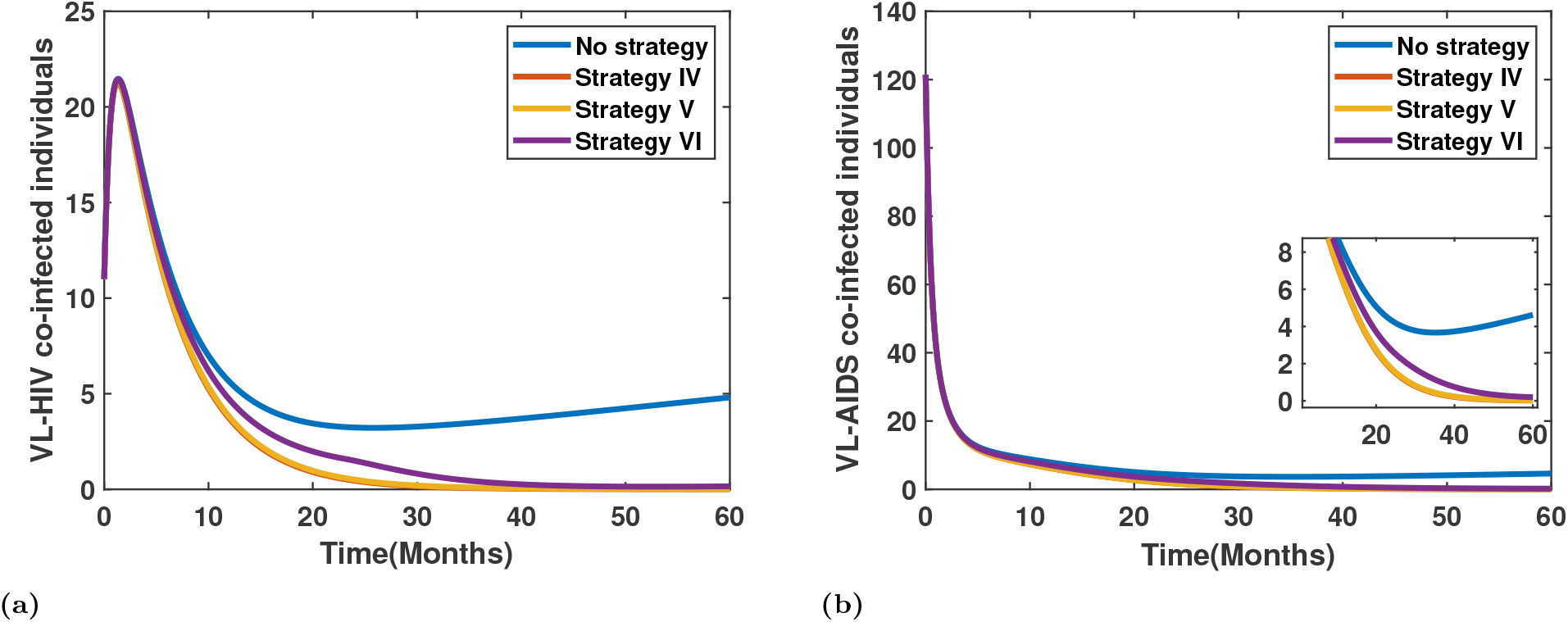
Variation in the infected population under Scenario B.

Figure 19 presents the optimal control profiles for the combined control strategies applied in pairs: (a) *u*_1_(*t*) and *u*_2_(*t*), (b) *u*_1_(*t*) and *u*_3_(*t*), and (c) *u*_2_(*t*) and *u*_3_(*t*).

**Fig 19.**
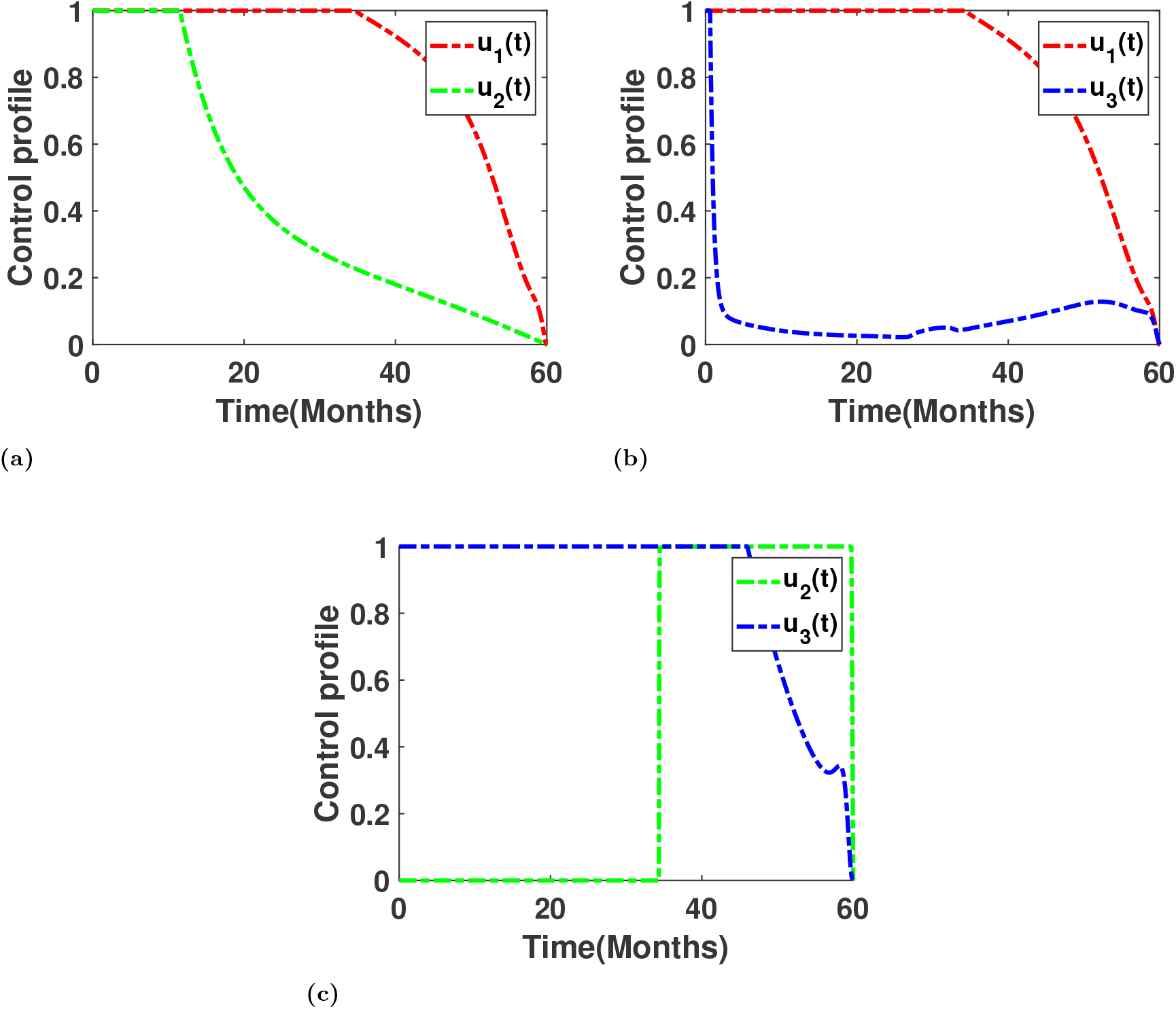
Control Profile when two controls applied

In all cases, the controls *u*_1_(*t*), *u*_2_(*t*), and *u*_3_(*t*) start at their maximum level, indicating the need for strong initial intervention to curb disease spread. For Strategy IV (*u*_1_(*t*), *u*_2_(*t*)), both controls begin at full strength, but *u*_1_(*t*) gradually declines after approximately 40 months, while *u*_2_(*t*) decreases smoothly throughout the period. This reflects that early and continuous prevention, coupled with gradual treatment or behavioral intervention, effectively reduces co-infection prevalence.

In Strategy V (*u*_1_(*t*), *u*_3_(*t*)), *u*_1_(*t*) maintains its maximum level for a longer period, while *u*_3_(*t*) remains steady before gradually decreasing, suggesting that sustained preventive efforts combined with targeted recovery or treatment control yield long-term benefits.

For Strategy VI (*u*_2_(*t*), *u*_3_(*t*)), *u*_2_(*t*) stays inactive for most of the initial period and activates only toward the later phase, whereas *u*_3_(*t*) follows a delayed decline pattern. This indicates that interventions associated with *u*_2_(*t*) are most beneficial in the later stages of the epidemic when co-infection levels are lower.

Overall, the figure highlights that early, aggressive, and sustained interventions particularly those combining *u*_1_(*t*) and *u*_2_(*t*) are most effective in minimizing co-infection levels.

### 5.4 Scenario C: All Three Controls Applied

#### 1. Strategy VII

All three controls *u*_1_, *u*_2_, and *u*_3_ are applied. This strategy implements a comprehensive approach to simultaneously reduce the transmission rates of visceral leishmaniasis (VL) and HIV while increasing the mortality rate of sandflies. It integrates vector control measures, public awareness campaigns, safe sex education, needle exchange programs, and sandfly population control through insecticides, biological agents, and environmental management. By combining all three interventions, this strategy aims to achieve the most effective reduction in disease transmission and improve overall public health outcomes.

Figure 20 illustrates the effect of applying all three optimal controls simultaneously on the dynamics of *I*_*HL*_ and *I*_*AL*_. The comparison between the scenarios with and without control clearly shows a substantial reduction in infection levels under the controlled case, confirming the effectiveness of the combined intervention strategy. The simultaneous application of all controls significantly suppresses co-infection prevalence, demonstrating strong synergistic effects between the interventions.

**Fig 20.**
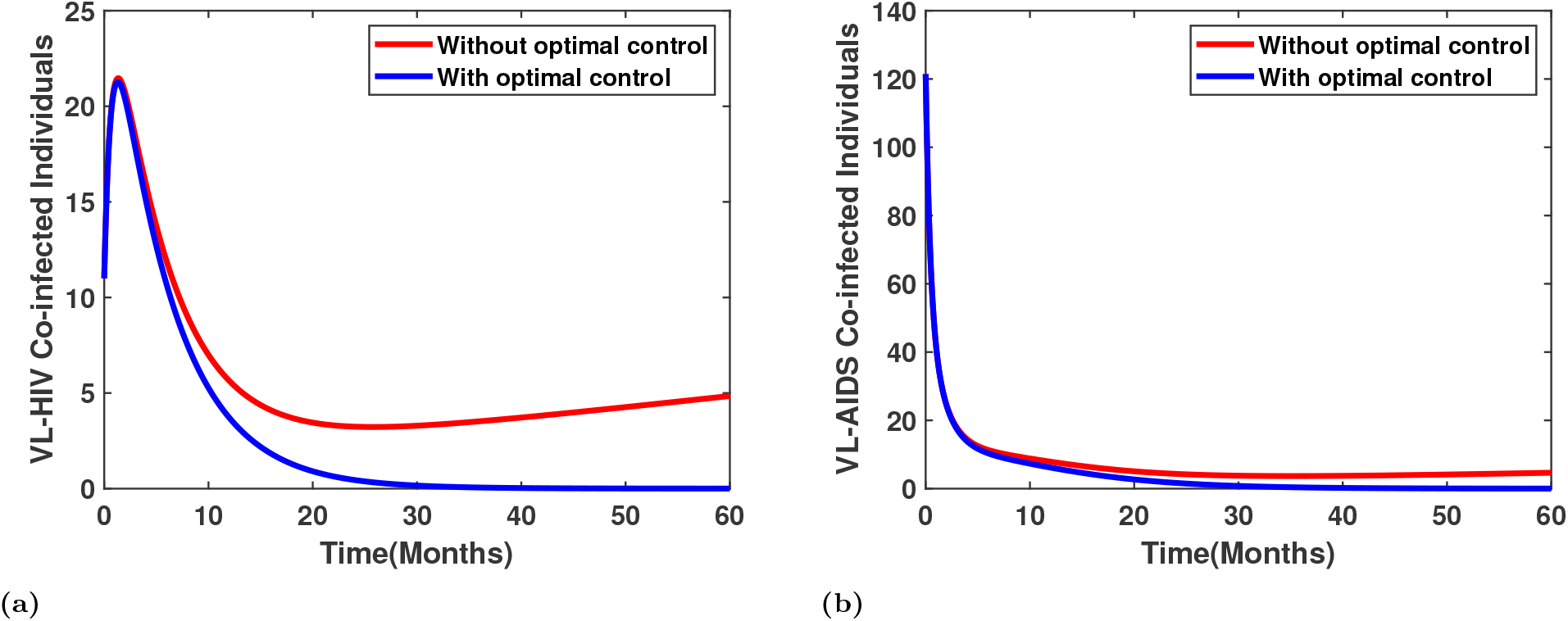
Variation in the infected population under Scenario C.

Figure 21 presents the corresponding optimal control profiles for *u*_1_(*t*), *u*_2_(*t*), and *u*_3_(*t*). The control efforts *u*_1_(*t*) and *u*_2_(*t*) remain at their maximum levels for most of the simulation period, showing the necessity of sustained intervention to curb disease transmission. In contrast, *u*_3_(*t*) stays relatively low with slight fluctuations before tapering off near the end, suggesting that moderate vector control is sufficient when preventive and educational measures are strongly enforced. Overall, the combined control approach proves to be the most efficient in reducing the burden of co-infection.

**Fig 21.**
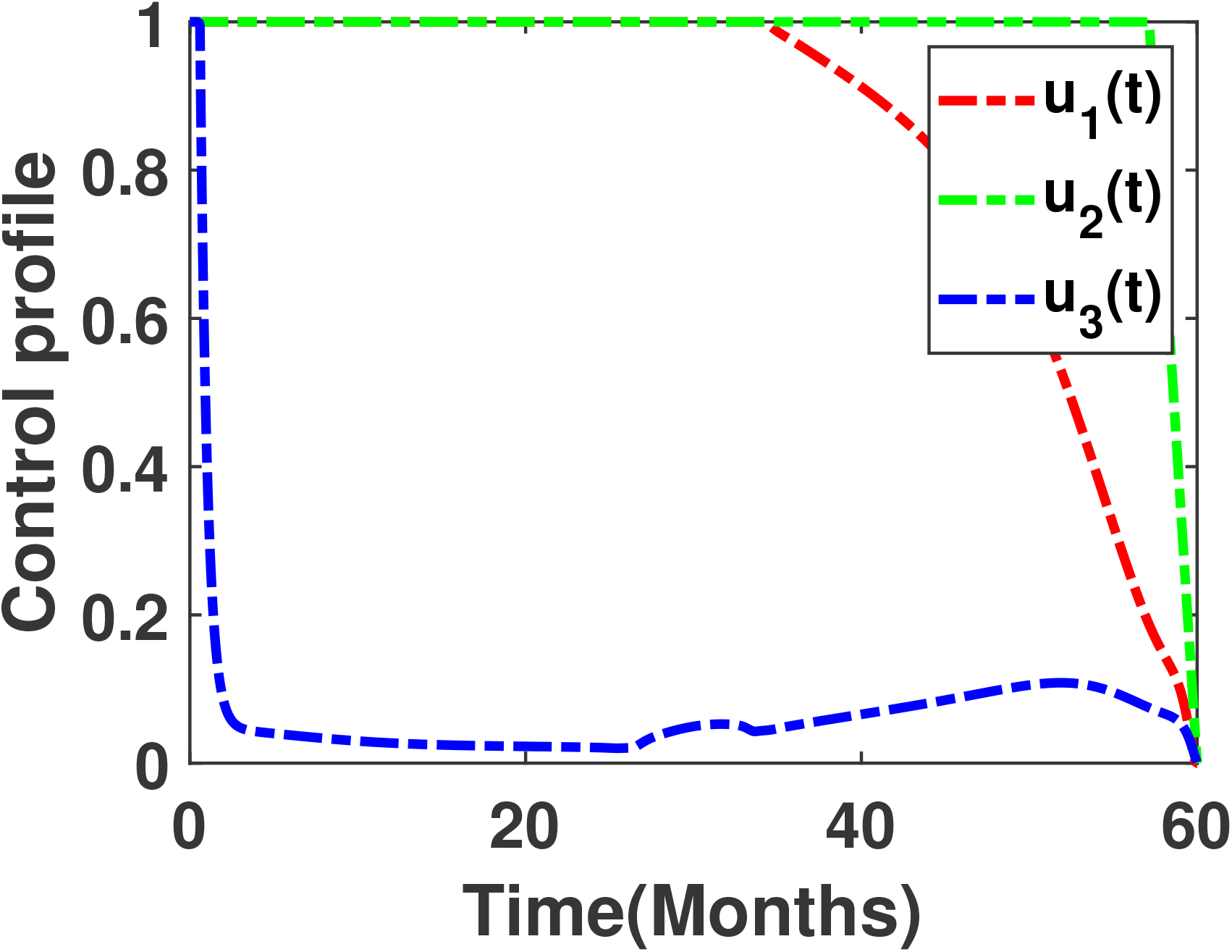
Control Profile when all three controls applied

### 5.5 Results of Optimal Control

This study examined the effectiveness of different optimal control strategies on the dynamics of VL–HIV co-infection. Three intervention scenarios were considered: **Scenario A**, where each control (*u*_1_, *u*_2_, and *u*_3_) was applied individually; **Scenario B**, where pairwise combinations of controls were implemented; and **Scenario C**, where all three controls were applied simultaneously.

The results from Scenario A revealed that the individual application of controls *u*_1_ achieved a greater reduction in co-infection prevalence compared to *u*_3_, *u*_2_, highlighting the stronger influence of preventive and vector-targeted interventions. In Scenario B, the combination of *u*_1_ and *u*_2_ proved to be the most effective pairwise strategy, substantially lowering the number of co-infected individuals. Finally, Scenario C demonstrated that the simultaneous implementation of all three controls—targeting VL transmission reduction, HIV transmission reduction, and increased sandfly mortality—produced the most pronounced decrease in infection levels. This comprehensive approach emphasizes the importance of integrating multiple intervention measures to effectively mitigate the burden of VL–HIV co-infection.

## 6 Conclusion

This study developed a comprehensive compartmental model to investigate the transmission dynamics of visceral leishmaniasis (VL), HIV, and their combined co-infection. The modeling framework was structured into three components: a VL-only subsystem, an HIV-only subsystem, and the complete VL-HIV co-infection model. For the VL-only and co-infection models, monthly VL and VL-HIV case data from Bihar, India obtained through the National Vector Borne Disease Control Programme (NVBDCP) were used for parameter estimation and model calibration. Sensitivity and partial rank correlation coefficient (PRCC) analyses were performed to identify the key parameters influencing the basic reproduction number (*R*_0*L*_) for VL transmission. For the HIV-only subsystem, yearly HIV incidence data from the National AIDS Control Organization (NACO), together with literature-based parameter values, were employed to examine the factors governing disease spread.

The analysis of the complete VL-HIV co-infection model revealed important epidemiological interactions between the two infections. Notably, even when VL cannot persist on its own (*R*_0*L*_ < 1), it can re-emerge and remain endemic in the presence of HIV (*R*_0*H*_ > 1), indicating that HIV infection drives the resurgence and sustained transmission of VL among co-infected individuals. Incorporating HIV treatment into the model further showed a substantial decrease in VL relapse rates and a significant reduction in co-infection prevalence, highlighting the crucial role of strengthening HIV treatment coverage in mitigating the overall VL burden in co-endemic regions.

The optimal control analysis showed that single interventions provide only limited reductions in disease prevalence, whereas combined strategies—particularly those that simultaneously reduce VL and HIV transmission and increase sandfly mortality—achieve substantial declines in both infections. These results underscore the importance of integrated vector management, sustained HIV treatment efforts, and community awareness interventions for effectively controlling and preventing VL-HIV co-infection in endemic settings.

Overall, this study provides a comprehensive understanding of the complex interaction between VL and HIV infections. Biologically, the results show that HIV-induced immune suppression facilitates the persistence and resurgence of VL, even in scenarios where VL would otherwise die out in isolation. This synergistic interaction increases disease severity, elevates relapse frequency, and complicates clinical management, emphasizing the importance of early diagnosis and integrated treatment strategies. The findings further demonstrate that strengthening HIV treatment coverage, together with effective vector control, can substantially reduce co-infection prevalence. From both epidemiological and public health perspectives, a coordinated approach that combines medical interventions, behavioral change initiatives, and vector-control measures is essential for mitigating the dual burden of VL-HIV co-infection in endemic regions.

## Data Availability

HIV data used are available online from National AIDS Control Organization (NACO) at https://naco.gov.in/.

## Funding

Funding for this study was provided by the Gates Foundation (INV-044445).

## Disclaimer

The work/opinion is based on research findings by the authors and not the opinion of the government.

## Notes

### Competing Interest Statement

The authors have declared no competing interest.

