## Supplemental File for "A Deterministic Approach to the Dynamics of Visceral Leishmaniasis and HIV Co-infection with Optimal Control"

### Supporting information

#### S.1 Derivation of the basic reproduction number

Construction of $\mathfrak{F}$ and $\mathfrak{V}$ matrices

The infection and transition terms are defined as

$$\mathfrak{F} = \begin{pmatrix} \frac{b\beta I_S S}{N} \\ 0 \\ 0 \\ 0 \\ \frac{c\beta S_S(I_L+I_D+I_P)}{N} \\ 0 \end{pmatrix}, \quad \mathfrak{V} = \begin{pmatrix} (\gamma_L+\lambda+\mu)L \\ (\theta+\sigma_L+\mu)I_L - \lambda L \\ (\omega+\gamma_D+\mu)I_D - \theta I_L \\ (\gamma_P+\mu)I_P - \omega I_D \\ (\mu_S+\lambda_S)E_S \\ \mu_S I_S - \lambda_S E_S \end{pmatrix}.$$

The Jacobian matrices of $\mathfrak{F}$ and $\mathfrak{V}$ with respect to the infected compartments are denoted by $F_1$ and $V_1$ respectively. The next generation matrix $F_1V_1^{-1}$ is given by

$$F_1V_1^{-1} = \begin{pmatrix} 0 & 0 & 0 & 0 & \frac{b\,\beta\,\lambda_s}{\mu_s\,(\lambda_s+\mu_s)} & \frac{b\,\beta}{\mu_s} \\ 0 & 0 & 0 & 0 & 0 & 0 \\ 0 & 0 & 0 & 0 & 0 & 0 \\ 0 & 0 & 0 & 0 & 0 & 0 \\ a_1 & a_2 & a_3 & \frac{\Lambda_s\,\beta\,c\,\mu}{\Lambda\,\mu_s\,(\gamma_p+\mu)} & 0 & 0 \\ 0 & 0 & 0 & 0 & 0 & 0 \end{pmatrix},$$

where

$$\begin{aligned} a_1 &= \frac{\Lambda_s\,\beta\,c\,\lambda\,\mu}{\Lambda\,\mu_s\,(\gamma_l+\lambda+\mu)\,(\mu+\sigma_l+\theta)} + \frac{\Lambda_s\,\beta\,c\,\lambda\,\mu\,\theta}{\Lambda\,\mu_s\,(\gamma_l+\lambda+\mu)\,(\gamma_d+\mu+\omega)\,(\mu+\sigma_l+\theta)} \\ &\quad + \frac{\Lambda_s\,\beta\,c\,\lambda\,\mu\,\omega\,\theta}{\Lambda\,\mu_s\,(\gamma_p+\mu)\,(\gamma_l+\lambda+\mu)\,(\gamma_d+\mu+\omega)\,(\mu+\sigma_l+\theta)}, \\ a_2 &= \frac{\Lambda_s\,\beta\,c\,\mu}{\Lambda\,\mu_s\,(\mu+\sigma_l+\theta)} + \frac{\Lambda_s\,\beta\,c\,\mu\,\theta}{\Lambda\,\mu_s\,(\gamma_d+\mu+\omega)\,(\mu+\sigma_l+\theta)} \\ &\quad + \frac{\Lambda_s\,\beta\,c\,\mu\,\omega\,\theta}{\Lambda\,\mu_s\,(\gamma_p+\mu)\,(\gamma_d+\mu+\omega)\,(\mu+\sigma_l+\theta)}, \\ a_3 &= \frac{\Lambda_s\,\beta\,c\,\mu}{\Lambda\,\mu_s\,(\gamma_d+\mu+\omega)} + \frac{\Lambda_s\,\beta\,c\,\mu\,\omega}{\Lambda\,\mu_s\,(\gamma_p+\mu)\,(\gamma_d+\mu+\omega)}. \end{aligned}$$

#### S.2 Proof of Theorem 2.1

The Jacobian of the system **??** at the disease-free equilibrium point $E_1^0$ is obtained as follows.

$$J_{E_1^0} = \begin{pmatrix} -\mu & 0 & 0 & 0 & 0 & \Omega & 0 & 0 & -b\beta \\ 0 & -a_1 & 0 & 0 & 0 & 0 & 0 & 0 & b\beta \\ 0 & \lambda & -a_2 & 0 & 0 & 0 & 0 & 0 & 0 \\ 0 & 0 & \theta & -a_3 & 0 & 0 & 0 & 0 & 0 \\ 0 & 0 & 0 & \omega & -a_4 & 0 & 0 & 0 & 0 \\ 0 & \gamma_L & 0 & \gamma_D & \gamma_P & -a_5 & 0 & 0 & 0 \\ 0 & 0 & \frac{-c\beta\Lambda_S\mu}{\mu_S\Lambda} & \frac{-c\beta\Lambda_S\mu}{\mu_S\Lambda} & \frac{-c\beta\Lambda_S\mu}{\mu_S\Lambda} & 0 & -\mu_S & 0 & 0 \\ 0 & 0 & \frac{c\beta\Lambda_S\mu}{\mu_S\Lambda} & \frac{c\beta\Lambda_S\mu}{\mu_S\Lambda} & \frac{c\beta\Lambda_S\mu}{\mu_S\Lambda} & 0 & 0 & -a_6 & 0 \\ 0 & 0 & 0 & 0 & 0 & 0 & 0 & \lambda_S & -\mu_S \end{pmatrix}$$

where

$$a_1 = (\gamma_L+\lambda+\mu),\ a_2 = (\theta+\sigma_L+\mu),\ a_3 = (\omega+\gamma_D+\mu),\ a_4 = (\gamma_P+\mu),$$

$$a_5 = (\mu + \Omega),\ a_6 = (\mu_S + \lambda_S).$$

The characteristic equation $|J_{E_1^0} - MI| = 0$ is given by

$$(M + \mu)(M + \mu_S)(M^7 + C_1M^6 + C_2M^5 + C_3M^4 + C_4M^3 + C_4M^2 + C_6M + C_7) = 0,$$

where

$$\begin{aligned}
C_1 &= (\sum_{i=1}^{6} a_i + \mu_S),\\
C_2 &= (\mu_S \sum_{i=1}^{6} + \sum_{1\le i<j\le 6} a_ia_j + \Omega\gamma_L),\\
C_3 &= (\mu_S \sum_{1\le i<j\le 6} a_ia_j + \sum_{1\le i<j<k\le 6} a_ia_ja_k + \Omega\gamma_L(a_6 + a_4 + a_2 + a_3 + \mu_S)),\\
C_4 &= (\mu_S \sum_{1\le i<j<k\le 6} a_ia_ja_k + \sum_{1\le i<j<k<l\le 6} a_ia_ja_ka_l + \Omega\gamma_L((a_6 + \mu_S)(a_4 + a_2 + a_3) + a_6\mu_S\\
&\quad + a_4a_3 + a_2a_3 + a_4a_2) - \frac{c\beta\Lambda_S\mu b\lambda\lambda_S}{\Lambda\mu_S}),\\
C_5 &= \mu_S \sum_{1\le i<j<k<l\le 6} a_ia_ja_ka_l + \sum_{1\le i<j<k<l<m\le 6} a_ia_ja_ka_la_m + \frac{c\beta\Lambda_S\mu}{\Lambda\mu_S}(\theta + a_3 + a_4 - b\beta\lambda a_5\lambda_S)\\
&\quad + \Omega\lambda\theta(\omega\gamma_P + \gamma_D a_4) + \Omega\gamma_L(a_4a_3 + a_2a_3 + a_4a_2)(\mu_S + a_6) + \Omega\theta\gamma_D(a_6 + \lambda\mu_S)\\
&\quad + \Omega\gamma_L(a_6\mu_S(a_4 + a_2 + a_3) + a_4a_2a_3),\\
C_6 &= \mu_S \sum_{1\le i<j<k<l<m\le 6} a_ia_ja_ka_la_m + \prod_{i=1}^{6} a_i + \frac{c\beta\Lambda_S\mu}{\Lambda\mu_S}(\theta\omega + \theta a_4 - b\beta\lambda a_5\lambda_S(\theta + a_3 + a_4)\\
&\quad \Omega\lambda\theta(\omega\gamma_p + \gamma_D a_4)(a_6 + \mu_S) + \Omega\gamma_L a_4a_2a_3(a_6 + \mu_S) + \Omega a_6\mu_S(\theta\gamma_D + a_4a_3 + a_2a_3 + a_4a_2)\\
C_7 &= \mu_S \prod_{i=1}^{6} a_i + a_6\Omega\lambda\theta\mu_S(\omega\gamma_P + \gamma_D a_4) + \Omega\gamma_L a_4a_2a_3a_6 - \frac{c\beta\Lambda_S\mu}{\Lambda\mu_S}(\theta\omega + \theta a_4 + a_3a_4)b\beta\lambda\lambda_S a_5
\end{aligned}$$

The two eigenvalues, $-\mu$, $-\mu_s$ are obtained directly. The remaining eigenvalues are found by solving the polynomial. Using the Routh-Hurwitz criteria, the system will be locally asymptotically stable if the following conditions hold:

$$\begin{aligned}
&C_i > 0, \quad \text{for } i = 1, 2, 3, 4, 5, 6, 7,\\
&C_1C_2 - C_3 > 0,\\
&(C_1C_2 - C_3)(C_1C_4 - C_2C_3) > C_1^2(C_3^2 - C_1C_5),\\
&(C_1C_2 - C_3)(C_1C_2C_3 - C_3^2 - C_2^2C_4) > C_1C_3(C_1C_4 - C_2^2),\\
&(C_1C_2 - C_3)(C_1C_2C_3C_4 - C_3^3 - C_2^2C_4C_3 - C_1C_2C_5 + C_3C_5) > C_1C_3(C_1C_4C_3 - C_2^2C_3 - C_1^2C_6),\\
&(C_1C_2 - C_3)(C_1C_2C_3C_4C_5 - C_3^4 - C_2^2C_4C_3^2 - C_1C_2C_5C_3 + C_3^2C_5 - C_1C_2C_6 + C_3C_6) > C_1C_3(C_1C_4C_3C_5\\
&-C_2^2C_3C_5 - C_1^2C_7).
\end{aligned}$$

Therefore, if the five previously stated inequalities are satisfied, $E_1^0$ is locally asymptotically stable.

#### S.3 Calculation of Bifurcation analysis

Let us denote $S = x_1,\ L_L = x_2,\ I_L = x_3,\ I_D = x_4,\ I_P = x_5,\ R = x_6,\ S_S = x_7,\ E_S = x_8,\ I_S = x_9$, so that the VL only model can be written as follows:

$$
\begin{aligned}
\frac{dS}{dt} &= \Lambda + \Omega R_L - (f_L + \mu)S, \\
\frac{dL_L}{dt} &= f_L S - (\gamma_L + \lambda + \mu)L_L, \\
\frac{dI_L}{dt} &= \lambda L_L - (\theta + \sigma_L + \mu)I_L, \\
\frac{dI_D}{dt} &= \theta I_L - (\omega + \gamma_D + \mu)I_D, \\
\frac{dI_P}{dt} &= \omega I_D - (\gamma_P + \mu)I_P, \\
\frac{dR_L}{dt} &= \gamma_P I_P + \gamma_D I_D + \gamma_L L - (\mu + \Omega)R_L, \\
\frac{dS_S}{dt} &= \Lambda_s - f_s S_S - \mu_s S_S, \\
\frac{dE_s}{dt} &= f_s S_S - (\mu_s + \lambda_s)E_S, \\
\frac{dI_S}{dt} &= \lambda_s E_S - \mu_s I_S. \qquad (1)
\end{aligned}
$$

where $f_L = \frac{b\beta I_S}{N},\ f_S = \frac{c\beta(I_L+I_D+I_P)}{N}$. The Jacobian of the above system at the DFE $E_1^0$ at the chosen bifurcation parameters $\beta$, obtained by equating $R_{L0} = 1$ is

$$
J_{E_1^0} = \begin{pmatrix}
-\mu & 0 & 0 & 0 & 0 & \Omega & 0 & 0 & -b\beta^* \\
0 & -a_1 & 0 & 0 & 0 & 0 & 0 & 0 & b\beta^* \\
0 & \lambda & -a_2 & 0 & 0 & 0 & 0 & 0 & 0 \\
0 & 0 & \theta & -a_3 & 0 & 0 & 0 & 0 & 0 \\
0 & 0 & 0 & \omega & -a_4 & 0 & 0 & 0 & 0 \\
0 & \gamma_L & 0 & \gamma_D & \gamma_P & -a_5 & 0 & 0 & 0 \\
0 & 0 & \frac{-c\beta^*\Lambda_S\mu}{\mu_S\Lambda} & \frac{-c\beta^*\Lambda_S\mu}{\mu_S\Lambda} & \frac{-c\beta^*\Lambda_S\mu}{\mu_S\Lambda} & 0 & -\mu_S & 0 & 0 \\
0 & 0 & \frac{c\beta^*\Lambda_S\mu}{\mu_S\Lambda} & \frac{c\beta^*\Lambda_S\mu}{\mu_S\Lambda} & \frac{c\beta^*\Lambda_S\mu}{\mu_S\Lambda} & 0 & 0 & -a_6 & 0 \\
0 & 0 & 0 & 0 & 0 & 0 & 0 & \lambda_S & -\mu_S
\end{pmatrix}
$$

where

$$
a_1 = (\gamma_L + \lambda + \mu),\ a_2 = (\theta + \sigma_L + \mu),\ a_3 = (\omega + \gamma_D + \mu),\ a_4 = (\gamma_P + \mu),
$$
$$
a_5 = (\mu + \Omega),\ a_6 = (\mu_S + \lambda_S).
$$

In the analysis of the linearized system with $\beta = \beta^* =$, we observe that it exhibits zero eigenvalues. Leveraging center manifold theory [1, 2], we delve into the dynamics near $\beta = \beta^*$. Utilizing the theorem stated in [2], we demonstrate the stability of the VL only endemic equilibrium point ($E_1^*$). Subsequently, we derive the left and right eigenvector as a column matrix $w = [w_1, w_2, w_3, w_4, w_5, w_6, w_7, w_8, w_9]^T$, where

$$
w_1 = \frac{b\beta\lambda\Omega[(\gamma_P + \mu)(\omega + \gamma_D + \mu) + \gamma_D(\gamma_P + \mu) + \gamma_P\omega\theta]}{\mu(\gamma_P + \mu)(\omega + \gamma_D + \mu)(\gamma_L + \lambda + \mu)(\theta + \sigma_L + \mu)(\mu + \Omega)} - \frac{b\beta}{\mu},\ w_2 = \frac{b\beta w_9}{(\gamma_L + \lambda + \mu)},\ , w_8 = w_8 > 0
$$

$$
w_3 = \frac{b\beta\lambda}{(\gamma_L + \lambda + \mu)(\theta + \sigma_L + \mu)} w_9,\ w_4 = \frac{\theta b\beta\lambda}{(\omega + \gamma_D + \mu)(\gamma_L + \lambda + \mu)(\theta + \sigma_L + \mu)},
$$

$$
w_5 = \frac{\omega\theta b\beta\lambda}{(\gamma_P + \mu)(\omega + \gamma_D + \mu)(\gamma_L + \lambda + \mu)(\theta + \sigma_L + \mu)},\ w_8 = \frac{\mu_S}{\lambda_S} w_9,
$$

$$
w_6 = \frac{b\beta\lambda[(\gamma_P + \mu)(\omega + \gamma_D + \mu) + \gamma_D(\gamma_P + \mu) + \gamma_P\omega\theta]}{(\gamma_P + \mu)(\omega + \gamma_D + \mu)(\gamma_L + \lambda + \mu)(\theta + \sigma_L + \mu)(\mu + \Omega)},
$$

$$w_7 = \frac{cb\beta^2\Lambda_S\mu\lambda}{\Lambda\mu_S}\frac{(\gamma_P+\mu)(\omega+\gamma_D+\mu)+\theta(\gamma_P+\mu)+\omega\theta}{(\gamma_P+\mu)(\omega+\gamma_D+\mu)(\gamma_L+\lambda+\mu)(\theta+\sigma_L+\mu)},\ w_9 = w_9 > 0$$

and the left eigenvector (represented as a row matrix) associated with the zero eigenvalue is obtained as $v = [v_1, v_2, v_3, v_4, v_5, v_6, v_7, v_8, v_9]$, where

$$v_1 = 0,\ v_2 = v_2 > 0,\ v_3 = \frac{(\gamma_L+\lambda+\mu)}{\lambda}v_2,$$
$$v_4 = \frac{(\theta+\sigma_L+\mu)(\gamma_L+\lambda+\mu)(\mu_S+\lambda_S)\mu_S^2\Lambda - c\beta\Lambda_S\mu\lambda^2\beta b}{\theta\lambda\mu_S(\mu_S+\lambda_S)}v_2,$$
$$v_5 = \frac{(\gamma_L+\lambda+\mu)}{\lambda}v_2,\ v_6 = v_7 = 0,\ v_8 = \frac{\lambda_S b\beta v_2}{\mu_S(\mu_S+\lambda_S)},\ v_9 = \frac{b\beta v_2}{\mu_S}.$$

Proceeding with this theorem, to compute the values of a and b for conduction, the bifurcations analysis. This involves finding the non-zero partial derivatives and our formulated system (1) at the disease-free equilibrium point ($E_1^0$). These partial derivatives are

$$\frac{\partial g_8}{\partial x_7\partial x_3} = \frac{c\beta\mu}{\Lambda} = \frac{\partial g_8}{\partial x_7\partial x_3}$$
$$\frac{\partial g_8}{\partial x_7\partial x_4} = \frac{c\beta\mu}{\Lambda} = \frac{\partial g_8}{\partial x_7\partial x_4}$$
$$\frac{\partial g_8}{\partial x_7\partial x_5} = \frac{c\beta\mu}{\Lambda} = \frac{\partial g_8}{\partial x_7\partial x_5}$$
$$\frac{\partial g_2}{\partial x_9\partial \beta} = b,\ \frac{\partial g_8}{\partial x_5\partial \beta} = \frac{c\Lambda_S\mu}{\Lambda\mu_S}$$
$$\frac{\partial g_8}{\partial x_3\partial \beta} = \frac{c\Lambda_S\mu}{\Lambda\mu_S},\ \frac{\partial g_8}{\partial x_4\partial \beta} = \frac{c\Lambda_S\mu}{\Lambda\mu_S}$$
$$a = 2\frac{c\beta\mu v_8 w_7(w_3+w_4+w_5)}{\Lambda} < 0,\ \text{since}\ \ w_7 < 0$$
$$b = bv_2w_8 + \frac{c\Lambda_S\mu}{\Lambda\mu_S}v_8(w_3+w_4+w_5) > 0$$

#### S.4 Derivation of the basic reproduction number for the human subsystem

Construction of $\mathfrak{F}_2$ and $\mathfrak{V}_2$ matrices.

The new infection and transition terms for the compartments $I_H$, $I_A$, and $T$ are defined as

$$\mathfrak{F}_2 = \begin{pmatrix} \frac{\beta_H(I_H+\alpha_H I_A)S}{N} \\ 0 \\ 0 \end{pmatrix},\quad \mathfrak{V}_2 = \begin{pmatrix} (\psi+\nu+\mu)I_H - \tau T \\ (\tau+\mu)T - \psi I_H \\ (\sigma_A+\mu)I_A - \nu I_H \end{pmatrix}.$$

The Jacobian matrices of $\mathfrak{F}_2$ and $\mathfrak{V}_2$ with respect to the infected compartments are denoted by $F_2$ and $V_2$, respectively. The next generation matrix is then given by

$$F_2V_2^{-1} = \begin{pmatrix} \frac{\beta_H(\tau+\mu)(\sigma_A+\mu+\alpha_H\nu)}{(\sigma_A+\mu)[(\psi+\nu+\mu)(\tau+\mu)-\psi\tau]} & \frac{\beta_H\tau(\sigma_A+\mu+\alpha_H\nu)}{(\sigma_A+\mu)[(\psi+\nu+\mu)(\tau+\mu)-\psi\tau]} & \frac{\alpha_H\beta_H}{\mu+\sigma_A} \\ 0 & 0 & 0 \\ 0 & 0 & 0 \end{pmatrix}.$$

The largest eigenvalue of $F_2V_2^{-1}$ gives the basic reproduction number, which simplifies to

$$R_{0H} = \frac{\beta_H(\tau+\mu)(\sigma_A+\mu+\alpha_H\nu)}{(\sigma_A+\mu)\left[(\psi+\nu+\mu)(\tau+\mu)-\psi\tau\right]}.$$

#### S.5 Proof of Theorem 3.1

The Jacobian of the system at the disease-free equilibrium point $E_2^0$ is obtained as follows,

$$J_{E_2^0} = \begin{pmatrix} -\mu & -\beta_H & 0 & -\alpha_H\beta_H \\ 0 & \beta_H - (\psi+\nu+\mu) & \tau & \alpha_H\beta_H \\ 0 & \psi & -(\tau+\mu) & 0 \\ 0 & \nu & 0 & -(\sigma_A+\mu), \end{pmatrix}$$

The characteristic equation $|J_{E_2^0} - MI| = 0$, here one eigenvalue is found directly $-\mu$ and other three eigen values are roots of the following polynomial

$$M^3 + C_1M^2 + C_2M + C_3 = 0,$$

where,

$$C_1 = (\psi + \nu + \tau + \sigma_A + 3\mu - \beta_H)$$

$$C_2 = (\psi+\nu+\mu-\beta_H)(\tau+\mu) + (\psi+\nu+\mu-\beta_H)(\sigma_A+\mu) + (\tau+\mu)(\sigma_A+\mu) - \psi\tau - \nu\alpha_H\beta_H$$

$$C_3 = (1 - R_{0H})(\sigma_A+\mu)((\psi+\nu+\mu)(\tau+\mu) - \psi\tau)$$

Using the Routh-Hurwitz criteria, if the conditions $C_1 > 0$, $C_3 > 0$, and $C_1C_2 > C_3$ are satisfied, the system is locally asymptotically stable. In this case, it is clear that $C_1 > 0$, $C_3 > 0$, and $C_1C_2 > C_3$, so $E_0$ is locally asymptotically stable.

#### S.6 Calculation of Bifurcation analysis

Let us denote $S = x_1,\ I_H = x_2,\ T = x_3,\ I_A = x_4$, so that the HIV only model can be written as follows:

$$\begin{aligned} \frac{dx_1}{dt} = g_1 &= \Lambda - (\frac{\beta_H(I_H + \alpha_H}{N} + \mu)x_1, \\ \frac{dx_2}{dt} = g_2 &= \frac{\beta_H(I_H + \alpha_H}{N}x_1 + \tau x_3 - (\psi+\nu+\mu)x_2, \\ \frac{dx_3}{dt} = g_3 &= \psi x_2 - (\tau+\mu)x_3, \\ \frac{dx_4}{dt} = g_4 &= \nu x_2 - (\mu+\sigma_A)x_4,. \end{aligned} \quad (2)$$

The Jacobian of the above system at the DFE $E_2^0$ at the chosen bifurcation parameters $\beta_H$, obtained by equating $R_{H0} = 1$ is

$$\beta_H = \frac{(\sigma_A+\mu)((\psi+\nu+\mu)(\tau+\mu) - \psi\tau)}{(\tau+\mu)(\sigma_A+\mu+\alpha_H\nu)}$$

$$J_{E_2^0} = \begin{pmatrix} -\mu & -\beta_H^* & 0 & -\alpha_H\beta_H^* \\ 0 & \beta_H^* - (\psi+\nu+\mu) & \tau & \alpha_H\beta_H^* \\ 0 & \psi & -(\tau+\mu) & 0 \\ 0 & \nu & 0 & -(\sigma_A+\mu), \end{pmatrix}$$

In the analysis of the linearized system with $\beta_H = \beta_H^* =$, we observe that it exhibits zero eigenvalues. Leveraging center manifold theory [1,2], we delve into the dynamics near $\beta_H = \beta_H^*$. Utilizing the theorem stated in [2], we demonstrate the stability of the HIV only endemic equilibrium point ($E_2^*$). Subsequently, we derive the left and right eigenvector as a column matrix $w = [w_1, w_2, w_3, w_4]^T$, where

$$w_1 = -\frac{\beta_H^*(\sigma_A+\mu+\alpha_H\nu)}{\mu(\sigma_A+\mu)}w_2,\ w_2 = w_2,\ w_3 = \frac{\psi}{\tau+\mu}w_2,\ w_4 = \frac{\nu}{\sigma_A+\mu}w_2$$

and the left eigenvector (represented as a row matrix) associated with the zero eigenvalue is obtained as $v = [v_1, v_2, v_3, v_4]$, where

$$v_1 = 0,\ v_2 = v_2,\ v_3 = \frac{\tau v_2}{\tau + \mu},\ v_4 = \frac{\alpha_H \beta_H v_2}{\sigma_A + \mu}$$

Proceeding with this theorem, to compute the values of a and b for conduction, the bifurcations analysis. This involves finding the non-zero partial derivatives along with our formulated system (2) at the disease-free equilibrium point ($E_1^0$). These partial derivatives are

$$\frac{\partial g_2}{\partial x_1 \partial x_2} = \frac{\beta_H \mu}{\Lambda} = \frac{\partial g_2}{\partial x_2 \partial x_1}$$

$$\frac{\partial g_2}{\partial x_1 \partial x_4} = \frac{\alpha_H \beta_H \mu}{\Lambda} = \frac{\partial g_2}{\partial x_4 \partial x_1}$$

$$\frac{\partial g_2}{\partial x_2 \partial \beta_H} = 1,\ \frac{\partial g_2}{\partial x_2 \partial \beta_H} = \alpha_H$$

$$a = -2(\frac{\beta_H^2 \mu}{\Lambda})(\frac{\sigma_A + \mu + \alpha_H \nu}{\sigma_A + \mu})^2 w_2^2 v_2 < 0,$$

$$b = 2\frac{\sigma_A + \mu + \alpha_H \nu}{\sigma_A + \mu} v_2 w_2 > 0$$

#### S.7 Proof of Theorem 4.1

$$
J = \begin{pmatrix}
-\mu & 0 & 0 & 0 & 0 & \Omega & -\beta_H & -\beta_H & -\beta_H & -\beta_H & -\beta_H & 0 & -\beta_H\alpha_H & -\beta_H\alpha_H & -\beta_H\alpha_H & -\beta_H\alpha_H & -\beta_H\alpha_H & 0 & 0 & 0 & 0 & -b\beta \\
0 & -a_1 & 0 & 0 & 0 & 0 & 0 & 0 & 0 & 0 & 0 & 0 & 0 & 0 & 0 & 0 & 0 & 0 & 0 & 0 & 0 & b\beta \\
0 & \lambda & -a_2 & 0 & 0 & 0 & 0 & 0 & 0 & 0 & 0 & 0 & 0 & 0 & 0 & 0 & 0 & 0 & 0 & 0 & 0 & 0 \\
0 & 0 & \theta & -a_3 & 0 & 0 & 0 & 0 & 0 & 0 & 0 & 0 & 0 & 0 & 0 & 0 & 0 & 0 & 0 & 0 & 0 & 0 \\
0 & 0 & 0 & \omega & -a_4 & 0 & 0 & 0 & 0 & 0 & 0 & 0 & 0 & 0 & 0 & 0 & 0 & 0 & 0 & 0 & 0 & 0 \\
0 & \gamma_L & 0 & \gamma_D & \gamma_P & -a_5 & 0 & 0 & 0 & 0 & 0 & 0 & 0 & 0 & 0 & 0 & 0 & 0 & 0 & 0 & 0 & 0 \\
0 & 0 & 0 & 0 & 0 & 0 & -a_6 & \beta_H & \beta_H & \beta_H & \beta_H & 0 & \beta_H\alpha_H & \beta_H\alpha_H & \beta_H\alpha_H & \beta_H\alpha_H & \beta_H\alpha_H & \theta_1\Omega & 0 & 0 & 0 & 0 \\
0 & 0 & 0 & 0 & 0 & 0 & 0 & -a_7 & 0 & 0 & 0 & 0 & 0 & 0 & 0 & 0 & 0 & 0 & 0 & 0 & 0 & 0 \\
0 & 0 & 0 & 0 & 0 & 0 & 0 & \alpha_1\lambda & -a_8 & \xi_1 & 0 & 0 & 0 & 0 & 0 & 0 & 0 & 0 & 0 & 0 & 0 & 0 \\
0 & 0 & 0 & 0 & 0 & 0 & 0 & 0 & \kappa_1\theta & -a_9 & 0 & 0 & 0 & 0 & 0 & 0 & 0 & 0 & 0 & 0 & 0 & 0 \\
0 & 0 & 0 & 0 & 0 & 0 & 0 & 0 & 0 & \rho_1\omega & -a_{10} & 0 & 0 & 0 & 0 & 0 & 0 & 0 & 0 & 0 & 0 & 0 \\
0 & 0 & 0 & 0 & 0 & 0 & \psi & \phi_2\psi & \phi_4\psi & \phi_1\psi & \phi_3\psi & a_{11} & 0 & 0 & 0 & 0 & 0 & \psi & 0 & 0 & 0 & 0 \\
0 & 0 & 0 & 0 & 0 & 0 & \nu & 0 & 0 & 0 & 0 & 0 & -a_{12} & 0 & 0 & 0 & 0 & 0 & \theta_2\Omega & 0 & 0 & 0 \\
0 & 0 & 0 & 0 & 0 & 0 & 0 & \delta_2\nu & 0 & 0 & 0 & 0 & 0 & -a_{13} & 0 & 0 & 0 & 0 & 0 & 0 & 0 & 0 \\
0 & 0 & 0 & 0 & 0 & 0 & 0 & 0 & \delta_4\nu & 0 & 0 & 0 & 0 & \alpha_2\lambda & -a_{14} & \xi_2 & 0 & 0 & 0 & 0 & 0 & 0 \\
0 & 0 & 0 & 0 & 0 & 0 & 0 & 0 & 0 & \delta_1\nu & 0 & 0 & 0 & 0 & \kappa_2\theta & -a_{15} & 0 & 0 & 0 & 0 & 0 & 0 \\
0 & 0 & 0 & 0 & 0 & 0 & 0 & 0 & 0 & 0 & \delta_3\nu & 0 & 0 & 0 & 0 & \rho_2\omega & -a_{16} & 0 & 0 & 0 & 0 & 0 \\
0 & 0 & 0 & 0 & 0 & 0 & 0 & \varphi_1\gamma_L & 0 & \psi_1\gamma_D & \zeta_1\gamma_P & 0 & 0 & 0 & 0 & 0 & 0 & -a_{17} & 0 & 0 & 0 & 0 \\
0 & 0 & 0 & 0 & 0 & 0 & 0 & 0 & 0 & 0 & 0 & 0 & 0 & \varphi_2\gamma_L & 0 & \psi_2\gamma_D & \zeta_1\gamma_P & \nu & -a_{18} & 0 & 0 & 0 \\
0 & 0 & -a_{19} & -a_{19} & -a_{19} & 0 & 0 & 0 & -\alpha_S a_{19} & -\alpha_S a_{19} & -\alpha_S a_{19} & 0 & 0 & 0 & -\omega_S a_{19} & -\omega_S a_{19} & -\omega_S a_{19} & 0 & 0 & -\mu_S & 0 & 0 \\
0 & 0 & a_{19} & a_{19} & a_{19} & 0 & 0 & 0 & \alpha_S a_{19} & \alpha_S a_{19} & \alpha_S a_{19} & 0 & 0 & 0 & \omega_S a_{19} & \omega_S a_{19} & \omega_S a_{19} & 0 & 0 & 0 & -a_{20} & 0 \\
0 & 0 & 0 & 0 & 0 & 0 & 0 & 0 & 0 & 0 & 0 & 0 & 0 & 0 & 0 & 0 & 0 & 0 & 0 & 0 & \lambda_S & -\mu_S
\end{pmatrix}
$$

where,

$$a_1 = (\gamma_L + \lambda + \mu),\ a_2 = (\theta + \sigma_L + \mu),\ a_3 = (\omega + \gamma_D + \mu), a_4 = (\gamma_P + \mu),\ a_5 = (\mu + \Omega),$$

$$a_6 = (\psi + \nu + \mu - \beta_H),\ a_7 = (\varphi_1\gamma_L + \phi_2\psi + \alpha_1\lambda + \delta_2\nu + \mu),\ a_8 = (\kappa_1\theta + \phi_4\psi + \delta_4\nu + \gamma_1\sigma_L + \mu),$$

$$a_9 = (\xi_1 + \psi_1\gamma_D + \phi_1\psi + \mu + \rho_1\omega + \delta_1\nu),\ a_{10} = (\zeta_1\gamma_P + \delta_3\nu + \phi_3\psi + \mu),\ a_{11} = (\tau + \mu),\ a_{12} = \sigma_A + \mu),$$

$$a_{13} = (\varphi_2\gamma_L + \sigma_A + \mu + \alpha_2\lambda).\ a_{14} = (\kappa_2\theta + \gamma_1\sigma_L + \gamma_2\sigma_A + \mu),\ a_{15} = (\xi_2 + \sigma_A + \mu + \psi_2\gamma_D + \rho_2\omega),$$

$$a_{16} = (\zeta_2\gamma_P + \sigma_A + \mu),\ a_{17} = (\theta_1\Omega + \psi + \nu + \mu),\ a_{18} = (\theta_2\Omega + \sigma_A + \mu),\ a_{19} = \frac{c\beta\Lambda_S\mu}{\Lambda\mu_S},\ a_{20} = (\lambda_S + \mu_S).$$

The characteristic equation

$$|J_{E^0} - MI| = P(M) = 0,$$

12 eigenvalues are found directly:

$$\begin{aligned} P(M) &= (M+\mu)(M+\mu_S)(M+a_3)(M+a_4)(M+a_5)(M+a_7)(M+a_{10})(M+a_{11})(M+a_{13}) \\ &\quad (M+a_{16})(M+a_{17})(M+a_{18})Q(M), \end{aligned}$$

where

$$\begin{aligned} Q(M) &= [(M+a_1)(M+a_2)(M+a_{20})(M+\mu_S) - a_{19}\lambda\lambda_S b\beta] \\ &\quad \times[M^2 + (a_8+a_9)M + a_8a_9 - \kappa_1\theta\xi_1] \\ &\quad \times[M^2 + (a_{14}+a_{15})M + a_{14}a_{15} - \kappa_2\theta\xi_2] \\ &\quad \times[M^2 + (a_6+a_{12})M + a_6a_{12} - \alpha_H\beta_H\nu]. \end{aligned}$$

#### S.8 Calculation of Optimal Control

$$
\begin{aligned}
\frac{d\lambda_1}{dt} = -\frac{\partial \mathcal{H}_C}{\partial S} &= f_1\frac{(N-S)}{N^2}(\lambda_1-\lambda_2)+f_3\frac{(N-S)}{N^2}(\lambda_1-\lambda_7)+\epsilon_2 f_3\frac{L}{N^2}(\lambda_8-\lambda_2)+\epsilon_4 f_3\frac{I_l}{N^2}(\lambda_9-\lambda_3)\\
&+\epsilon_1 f_3\frac{I_D}{N^2}(\lambda_{10}-\lambda_4)+\epsilon_3 f_3\frac{I_P}{N^2}(\lambda_{11}-\lambda_5)+f_3\frac{R}{N^2}(\lambda_{18}-\lambda_6)\\
&+\eta_1 f_1\frac{I_H}{N^2}(\lambda_8-\lambda_7)+\eta_2 f_1\frac{I_A}{N^2}(\lambda_{14}-\lambda_{13})+f_2\frac{S_S}{N^2}(\lambda_{21}-\lambda_{20})+\mu\lambda_1.\\
\frac{d\lambda_2}{dt} = -\frac{\partial \mathcal{H}_C}{\partial L} &= f_1\frac{S}{N^2}(\lambda_2-\lambda_1)+f_3\frac{S}{N^2}(\lambda_7-\lambda_1)+\lambda_b(\lambda_2-\lambda_3)+\gamma_l(\lambda_2-\lambda_6)\\
&+\epsilon_2 f_3\frac{(N-L)}{N^2}(\lambda_2-\lambda_8)+\epsilon_4 f_3\frac{I_l}{N^2}(\lambda_9-\lambda_3)+\epsilon_1 f_3\frac{I_d}{N^2}(\lambda_{10}-\lambda_4)\\
&+\epsilon_3 f_3\frac{I_P}{N^2}(\lambda_{11}-\lambda_5)+f_3\frac{R}{N^2}(\lambda_{18}-\lambda_6)+\eta_1 f_1\frac{I_H}{N^2}(\lambda_8-\lambda_7)\\
&+\eta_2 f_1\frac{I_A}{N^2}(\lambda_{14}-\lambda_{13})+f_2\frac{S_S}{N^2}(\lambda_{21}-\lambda_{20})+\mu\lambda_2,\\
\frac{d\lambda_3}{dt} = -\frac{\partial \mathcal{H}_C}{\partial I_L} &= \frac{f_1 S}{N^2}(\lambda_2-\lambda_1)+\frac{f_3 S}{N^2}(\lambda_7-\lambda_1)+\epsilon_2 f_3\frac{L}{N^2}(\lambda_8-\lambda_2)+\epsilon_4 f_3\frac{(N-I_L)}{N^2}(\lambda_3-\lambda_9)\\
&+\theta(\lambda_3-\lambda_4)+\epsilon_1 f_3\frac{I_D}{N^2}(\lambda_{10}-\lambda_4)+\epsilon_3 f_3\frac{I_P}{N^2}(\lambda_{11}-\lambda_5)+f_3\frac{R}{N^2}(\lambda_{18}-\lambda_6)\\
&+\eta_1 f_1\frac{I_H}{N^2}(\lambda_8-\lambda_7)+\eta_2 f_1\frac{I_A}{N^2}(\lambda_{14}-\lambda_{13})+b_0 S_s(\lambda_{20}-\lambda_{21})+\lambda_3(\sigma_L+\mu)-W_1,\\
\frac{d\lambda_4}{dt} = -\frac{\partial \mathcal{H}_C}{\partial I_D} &= \mu\lambda_4-w_4+\frac{f_1 S}{N^2}(\lambda_2-\lambda_1)+\frac{f_3 S}{N^2}(\lambda_7-\lambda_1)+\epsilon_2 f_3\frac{L}{N^2}(\lambda_8-\lambda_2)\\
&+\epsilon_4 f_3\frac{I_L}{N^2}(\lambda_9-\lambda_3)+\omega(\lambda_4-\lambda_5)+\gamma_D(\lambda_4-\lambda_6)+\epsilon_1 f_3\frac{(N-I_D)}{N^2}(\lambda_4-\lambda_{10})\\
&+\epsilon_3 f_3\frac{I_P}{N^2}(\lambda_{11}-\lambda_5)+f_3\frac{R}{N^2}(\lambda_{18}-\lambda_6)+\eta_1 f_1\frac{I_H}{N^2}(\lambda_8-\lambda_7)+\eta_2 f_1\frac{I_A}{N^2}(\lambda_{14}-\lambda_{13})\\
&+b_1 S_S(\lambda_{20}-\lambda_{21}),\\
\frac{d\lambda_5}{dt} = -\frac{\partial \mathcal{H}_C}{\partial I_P} &= \mu\lambda_5-W_7+\frac{f_1 S}{N^2}(\lambda_2-\lambda_1)+\frac{f_3 S}{N^2}(\lambda_7-\lambda_1)+\epsilon_2 f_3\frac{L}{N^2}(\lambda_8-\lambda_2)\\
&+\epsilon_4 f_3\frac{I_L}{N^2}(\lambda_9-\lambda_3)+\gamma_P(\lambda_5-\lambda_6)+\epsilon_1 f_3\frac{I_D}{N^2}(\lambda_{10}-\lambda_4)\\
&+\epsilon_3 f_3\frac{(N-I_P)}{N^2}(\lambda_5-\lambda_{11})+f_3\frac{R}{N^2}(\lambda_{18}-\lambda_6)+\eta_1 f_1\frac{I_H}{N^2}(\lambda_8-\lambda_7)\\
&+\eta_2 f_1\frac{I_A}{N^2}(\lambda_{14}-\lambda_{13})+b_{01} S_S(\lambda_{20}-\lambda_{21}),\\
\frac{d\kappa_6}{dt} = -\frac{\partial \mathcal{H}_C}{\partial R_L} &= \mu\lambda_6+\frac{f_1 S(\lambda_2-\lambda_1)}{N^2}+\frac{f_3 S(\lambda_7-\lambda_1)}{N^2}+\Omega(\lambda_6-\lambda_1)+\frac{\epsilon_2 f_3 L(\lambda_8-\lambda_2)}{N^2}+\frac{\epsilon_4 f_3 I_l(\lambda_9-\lambda_3)}{N^2}\\
&+\frac{\epsilon_1 f_3 I_d(\lambda_{10}-\lambda_4)}{N^2}+\frac{\epsilon_3 f_3 I_p(\lambda_{11}-\lambda_5)}{N^2}+\frac{f_3(N-R)(\lambda_6-\lambda_{18})}{N^2}+\frac{\eta_1 f_1 I_h(\lambda_8-\lambda_7)}{N^2}\\
&+\frac{\eta_2 f_1 I_a(\lambda_{14}-\lambda_{13})}{N^2}+\frac{f_2 S_s(\lambda_{21}-\lambda_{20})}{N^2},\\
\frac{d\lambda_7}{dt} = -\frac{\partial \mathcal{H}_C}{\partial I_H} &= \mu\lambda_7-w_{11}+\frac{a_1 S(\lambda_1-\lambda_7)}{N^2}+\frac{f_1 S(\lambda_2-\lambda_1)}{N^2}+\epsilon_2 a_1 L(\lambda_2-\lambda_8)+\epsilon_4 I_l a_1(\lambda_3-\lambda_9)\\
&+\epsilon_1 I_d a_1(\lambda_4-\lambda_{10})+\epsilon_3 I_p a_1(\lambda_5-\lambda_{11})+a_1 R(\lambda_6-\lambda_{18})+\frac{\eta_2 I_a f_1(\lambda_{14}-\lambda_{13})}{N^2}\\
&+\psi(\lambda_7-\lambda_{12})+\nu(\lambda_7-\lambda_{13})+\frac{\eta_1 f_1(N-I_h)(\lambda_7-\lambda_8)}{N^2}+\frac{S_s f_2(\lambda_{21}-\lambda_{20})}{N^2}
\end{aligned}
$$

$$\begin{aligned}
\frac{d\lambda_8}{dt} = -\frac{\partial \mathcal{H}_C}{\partial I_{HL_L}} &= \mu\lambda_8 + \frac{a_2 S(\lambda_1 - \lambda_7)}{N^2} + \frac{f_1 S(\lambda_2 - \lambda_1)}{N^2} + \epsilon_2 a_2 L(\lambda_2 - \lambda_8) + \epsilon_4 I_l a_2(\lambda_3 - \lambda_9) \\
&+ \epsilon_1 I_d a_2(\lambda_4 - \lambda_{10}) + \epsilon_3 I_p a_2(\lambda_5 - \lambda_{11}) + a_2 R(\lambda_6 - \lambda_{18}) + \frac{\eta_2 I_a f_1(\lambda_{14} - \lambda_{13})}{N^2} \\
&+ \varphi_1\gamma_L(\lambda_8 - \lambda_{18}) + \phi_2\psi(\lambda_8 - \lambda_{12}) + \alpha_1\lambda(\lambda_8 - \lambda_9) + d_2\nu(\lambda_8 - \lambda_{14}) \\
&+ \frac{\eta_1 f_1 I_H(\lambda_7 - \lambda_8)}{N^2} + \frac{S_S f_2(\lambda_{21} - \lambda_{20})}{N^2}, \\
\frac{d\lambda_9}{dt} = -\frac{\partial \mathcal{H}_C}{\partial I_{HL}} &= (\mu + \sigma_L g_1)\lambda_9 - W_2 + \frac{a_4 S(\lambda_1 - \lambda_7)}{N^2} + \frac{f_1 S(\lambda_2 - \lambda_1)}{N^2} + \epsilon_2 a_4 L(\lambda_2 - \lambda_8) + \epsilon_4 I_l a_4(\lambda_3 - \lambda_9) \\
&+ \epsilon_1 I_d a_4(\lambda_4 - \lambda_{10}) + \epsilon_3 I_p a_4(\lambda_5 - \lambda_{11}) + a_4 R(\lambda_6 - \lambda_{18}) + \frac{\eta_2 I_a f_1(\lambda_{14} - \lambda_{13})}{N^2} \\
&+ \frac{\eta_1 f_1 I_h(\lambda_8 - \lambda_7)}{N^2} + k_1\theta(\lambda_9 - \lambda_{10}) + \phi_4\psi(\lambda_9 - \lambda_{12}) + d_4\nu(\lambda_9 - \lambda_{15}) \\
&+ \frac{S_s b_3(\lambda_{20} - \lambda_{21})}{N^2}, \\
\frac{d\lambda_{10}}{dt} = -\frac{\partial \mathcal{H}_C}{\partial I_{HD}} &= \mu\lambda_{10} - w_5 + \frac{a_3 S(\lambda_1 - \lambda_7)}{N^2} + \frac{f_1 S(\lambda_2 - \lambda_1)}{N^2} + \epsilon_2 a_3 L(\lambda_2 - \lambda_8) + \epsilon_4 I_l a_3(\lambda_3 - \lambda_9) \\
&+ \epsilon_1 I_d a_3(\lambda_4 - \lambda_{10}) + \epsilon_3 I_p a_3(\lambda_5 - \lambda_{11}) + a_3 R(\lambda_6 - \lambda_{18}) + \frac{\eta_2 I_a f_1(\lambda_{14} - \lambda_{13})}{N^2} \\
&+ \xi_1(\lambda_{10} - \lambda_9) + g_d\psi_1(\lambda_{10} - \lambda_{18}) + \phi_1\psi(\lambda_{10} - \lambda_{12}) + \rho_1\omega(\lambda_{10} - \lambda_{11}) \\
&+ d_1\nu(\lambda_{10} - \lambda_{16}) + \frac{\eta_1 f_1 I_h(\lambda_8 - \lambda_7)}{N^2} + \frac{S_s b_2(\lambda_{20} - \lambda_{21})}{N^2} \\
\frac{d\lambda_{11}}{dt} = -\frac{\partial \mathcal{H}_C}{\partial I_{HP}} &= \mu\lambda_{11} + w_8 + \frac{a_5 S(\lambda_1 - \lambda_7)}{N^2} + \frac{f_1 S(\lambda_2 - \lambda_1)}{N^2} + \epsilon_2 a_5 L(\lambda_2 - \lambda_8) + \epsilon_4 I_l a_5(\lambda_3 - \lambda_9) \\
&+ \epsilon_1 I_d a_5(\lambda_4 - \lambda_{10}) + \epsilon_3 I_p a_5(\lambda_5 - \lambda_{11}) + a_5 R(\lambda_6 - \lambda_{18}) + \frac{\eta_2 I_a f_1(\lambda_{14} - \lambda_{13})}{N^2} \\
&+ \zeta_1 g_p(\lambda_{11} - \lambda_{18}) + d_3\nu(\lambda_{17} - \lambda_{11}) + \phi_3\psi(\lambda_{11} - \lambda_{12}) \\
&+ \frac{\eta_1 f_1 I_h(\lambda_8 - \lambda_7)}{N^2} + \frac{S_s b_4(\lambda_{20} - \lambda_{21})}{N^2} \\
\frac{d\lambda_{12}}{dt} = -\frac{\partial \mathcal{H}_C}{\partial T} &= \mu\lambda_{12} + \frac{f_1 S(\lambda_2 - \lambda_1)}{N^2} + \frac{f_3 S(\lambda_7 - \lambda_1)}{N^2} + \frac{\epsilon_2 f_3 L(\lambda_8 - \lambda_2)}{N^2} + \frac{\epsilon_4 f_3 I_l(\lambda_9 - \lambda_3)}{N^2} \\
&+ \frac{\epsilon_1 f_3 I_d(\lambda_{10} - \lambda_4)}{N^2} + \frac{\epsilon_3 f_3 I_p(\lambda_{11} - \lambda_5)}{N^2} + \frac{f_3 R(\lambda_{18} - \lambda_6)}{N^2} + \frac{\eta_1 f_1 I_h(\lambda_8 - \lambda_7)}{N^2} \\
&+ \frac{\eta_2 f_1 I_a(\lambda_{14} - \lambda_{13})}{N^2} + \frac{f_2 S_s(\lambda_{21} - \lambda_{20})}{N^2} + \tau(\lambda_{12} - \lambda_7)
\end{aligned}$$

$$
\begin{aligned}
\frac{d\kappa_{13}}{dt} = -\frac{\partial \mathcal{H}_C}{\partial I_A} &= (\mu+\sigma_A)\lambda_{13} - w_{12} + \frac{a_6 S(\lambda_1-\lambda_7)}{N^2} + \frac{f_1 S(\lambda_2-\lambda_1)}{N^2} + \frac{\epsilon_2 a_6 L(\lambda_2-\lambda_8)}{N^2} \\
&+ \frac{\epsilon_4 I_l a_6(\lambda_3-\lambda_9)}{N^2} + \frac{\epsilon_1 I_d a_6(\lambda_4-\lambda_{10})}{N^2} + \frac{\epsilon_3 I_p a_6(\lambda_5-\lambda_{11})}{N^2} + \frac{a_6 R(\lambda_6-\lambda_{18})}{N^2} \\
&+ \frac{\eta_2(N-I_a) f_1(\lambda_{13}-\lambda_{14})}{N^2} + \frac{\eta_1 f_1 I_h(\lambda_8-\lambda_7)}{N^2} + \frac{S_s f_2(\lambda_{21}-\lambda_{20})}{N^2}, \\
\frac{d\lambda_{14}}{dt} = -\frac{\partial \mathcal{H}_C}{\partial I_{AL_L}} &= (\sigma_A+\mu)\lambda_{14} + \frac{a_7 S(\lambda_1-\lambda_7)}{N^2} + \frac{f_1 S(\lambda_2-\lambda_1)}{N^2} + \frac{\epsilon_2 a_7 L(\lambda_2-\lambda_8)}{N^2} \\
&+ \frac{\epsilon_4 I_l a_7(\lambda_3-\lambda_9)}{N^2} + \frac{\epsilon_1 I_d a_7(\lambda_4-\lambda_{10})}{N^2} + \frac{\epsilon_3 I_p a_7(\lambda_5-\lambda_{11})}{N^2} + \frac{a_7 R(\lambda_6-\lambda_{18})}{N^2} \\
&+ \frac{\eta_2 I_a f_1(\lambda_{14}-\lambda_{13})}{N^2} + \varphi_2\gamma_L(\lambda_{14}-\lambda_{19}) + \alpha_2\lambda b(\lambda_{14}-\lambda_{15}) \\
&+ \frac{\eta_1 f_1 I_h(\lambda_8-\lambda_7)}{N^2} + \frac{S_s f_2(\lambda_{21}-\lambda_{20})}{N^2}, \\
\frac{d\lambda_{15}}{dt} = -\frac{\partial \mathcal{H}_C}{\partial I_{AL}} &= (\mu+g_1\sigma_L+g_2\sigma_A)\lambda_{15} - w_3 + \frac{a_8 S(\lambda_1-\lambda_7)}{N^2} + \frac{f_1 S(\lambda_2-\lambda_1)}{N^2} + \frac{\epsilon_2 a_8 L(\lambda_2-\lambda_8)}{N^2} \\
&+ \frac{\epsilon_4 I_l a_8(\lambda_3-\lambda_9)}{N^2} + \frac{\epsilon_1 I_d a_8(\lambda_4-\lambda_{10})}{N^2} + \frac{\epsilon_3 I_p a_8(\lambda_5-\lambda_{11})}{N^2} + \frac{a_8 R(\lambda_6-\lambda_{18})}{N^2} \\
&+ \frac{\eta_2 I_a f_1(\lambda_{14}-\lambda_{13})}{N^2} + k_2\theta(\lambda_{15}-\lambda_{16}) + \frac{\eta_1 f_1 I_h(\lambda_8-\lambda_7)}{N^2} + \frac{S_s b_5(\lambda_{20}-\lambda_{21})}{N^2}. \\
\frac{d\lambda_{16}}{dt} = -\frac{\partial \mathcal{H}_C}{\partial I_{AD}} &= (\mu+\sigma_A)\lambda_{16}(j+1) - w_6 + a_9 S(j+1)\frac{\lambda_1(j+1)-\lambda_7(j+1)}{N^2} \\
&+ \frac{f_1 S(j+1)}{N^2}(\lambda_2(j+1)-\lambda_1(j+1)) \quad + \epsilon_2 a_9 L(j+1)(\lambda_2(j+1)-\lambda_8(j+1)) \\
&\quad + \epsilon_1 I_d(j+1) a_9(\lambda_4(j+1)-\lambda_{10}(j+1)) + \epsilon_3 I_p(j+1) a_9(\lambda_5(j+1)-\lambda_{11}(j+1)) \\
&\quad + a_9 R(j+1)(\lambda_6(j+1)-\lambda_{18}(j+1)) + \eta_2 I_a(j+1) f_1 \frac{\lambda_{14}(j+1)-\lambda_{13}(j+1)}{N^2} \\
&\quad + \xi_2(\lambda_{16}(j+1)-\lambda_{15}(j+1)) + \psi_2 g_d(\lambda_{16}(j+1)-\lambda_{19}(j+1)) \\
&\quad + \rho_2\omega(\lambda_{16}(j+1)-\lambda_{17}(j+1)) + \epsilon_4 I_l(j+1) a_9(\lambda_3(j+1)-\lambda_9(j+1)) \\
&+ \eta_1 f_1 I_h(j+1)\frac{\lambda_8(j+1)-\lambda_7(j+1)}{N^2} + S_s(j+1) b_6 \frac{\lambda_{20}(j+1)-\lambda_{21}(j+1)}{N^2}, \\
\frac{d\lambda_{17}}{dt} = -\frac{\partial \mathcal{H}_C}{\partial I_{AP}} &= (\mu+\sigma_A)\lambda_{17}(j+1) - w_9 + a_{10} S(j+1)\frac{\lambda_1(j+1)-\lambda_7(j+1)}{N^2} \\
&\quad + \epsilon_2 a_{10} L(j+1)(\lambda_2(j+1)-\lambda_8(j+1)) + \epsilon_4 I_l(j+1) a_{10}(\lambda_3(j+1)-\lambda_9(j+1)) \\
&\quad + \epsilon_1 I_d(j+1) a_{10}(\lambda_4(j+1)-\lambda_{10}(j+1)) + \epsilon_3 I_p(j+1) a_{10}(\lambda_5(j+1)-\lambda_{11}(j+1)) \\
&\quad + a_{10} R(j+1)(\lambda_6(j+1)-\lambda_{18}(j+1)) + \eta_2 I_a(j+1) f_1 \frac{\lambda_{14}(j+1)-\lambda_{13}(j+1)}{N^2} \\
&\quad + \zeta_2 g_p(\lambda_{17}(j+1)-\lambda_{19}(j+1)) + \eta_1 f_1 I_h(j+1)\frac{\lambda_8(j+1)-\lambda_7(j+1)}{N^2} \\
&\quad + S_s(j+1) b_7 \frac{\lambda_{20}(j+1)-\lambda_{21}(j+1)}{N^2} + \frac{f_1 S(j+1)}{N^2}(\lambda_2(j+1)-\lambda_1(j+1))
\end{aligned}
$$

$$
\begin{aligned}
\frac{d\lambda_{18}}{dt} = -\frac{\partial \mathcal{H}_C}{\partial I_{HR_L}} &= \mu\lambda_{18}(j+1) + f_1 S(j+1)(j+1)\frac{\lambda_2(j+1)-\lambda_1(j+1)}{N^2} \\
&+f_3\frac{S(j+1)}{N^2}(\lambda_7(j+1)-\lambda_1(j+1)) + \nu(\lambda_{18}-\lambda_{19}) \\
&\quad +\epsilon_2 f_3\frac{L(j+1)}{N^2}(\lambda_8(j+1)-\lambda_2(j+1)) + \epsilon_4 f_3 I_l(j+1)\frac{\lambda_9(j+1)-\lambda_3(j+1)}{N^2} \\
&\quad +\epsilon_1 f_3 I_d(j+1)\frac{\lambda_{10}(j+1)-\lambda_4(j+1)}{N^2} + \psi(\lambda_{18}(j+1)-\lambda_{12}(j+1)) \\
&\quad +\epsilon_3 f_3 I_p(j+1)\frac{\lambda_{11}(j+1)-\lambda_5(j+1)}{N^2} + f_3 R(j+1)\frac{\lambda_{18}(j+1)-\lambda_6(j+1)}{N^2} \\
&\quad +\eta_1 f_1 I_h(j+1)\frac{\lambda_8(j+1)-\lambda_7(j+1)}{N^2} + \eta_2 f_1 I_a(j+1)\frac{\lambda_{14}(j+1)-\lambda_{13}(j+1)}{N^2} \\
&\quad +f_2 S_s(j+1)\frac{\lambda_{21}(j+1)-\lambda_{20}(j+1)}{N^2} + \theta_1\Omega(\lambda_{18}(j+1)-\lambda_7(j+1)) \\
\frac{d\lambda_{19}}{dt} = -\frac{\partial \mathcal{H}_C}{\partial I_{AR_L}} &= (\sigma_A+\mu)\lambda_{19}(j+1) + f_1 S(j+1)\frac{\lambda_2(j+1)-\lambda_1(j+1)}{N^2} + f_3\frac{S(j+1)}{N^2}(\lambda_7(j+1)-\lambda_1(j+1)) \\
&\quad +\epsilon_2 f_3\frac{L(j+1)}{N^2}(\lambda_8(j+1)-\lambda_2(j+1)) + \epsilon_4 f_3 I_l(j+1)\frac{\lambda_9(j+1)-\lambda_3(j+1)}{N^2} \\
&\quad +\epsilon_1 f_3 I_d(j+1)\frac{\lambda_{10}(j+1)-\lambda_4(j+1)}{N^2} + \epsilon_3 f_3 I_p(j+1)\frac{\lambda_{11}(j+1)-\lambda_5(j+1)}{N^2} \\
&\quad +f_3 R(j+1)\frac{\lambda_{18}(j+1)-\lambda_6(j+1)}{N^2} + \eta_1 f_1 I_h(j+1)\frac{\lambda_8(j+1)-\lambda_7(j+1)}{N^2} \\
&\quad +\eta_2 f_1 I_a(j+1)\frac{\lambda_{14}(j+1)-\lambda_{13}(j+1)}{N^2} + f_2 S_s(j+1)\frac{\lambda_{21}(j+1)-\lambda_{20}(j+1)}{N^2} \\
&\quad +\theta_2\Omega(\lambda_{19}(j+1)-\lambda_{13}(j+1)) \\
\frac{d\lambda_{20}}{dt} = -\frac{\partial \mathcal{H}_C}{\partial S_s} &= \lambda_{20}(j+1)(\mu_s + du_3(j+1)) + f_s(\lambda_{20}(j+1)-\lambda_{21}(j+1)) \\
\frac{d\lambda_{21}}{dt} = -\frac{\partial \mathcal{H}_C}{\partial E_s} &= \lambda_{21}(j+1)(\mu_s + du_3(j+1)) + \lambda_s(\lambda_{21}(j+1)-\lambda_{22}(j+1)), \\
\frac{d\lambda_{22}}{dt} = -\frac{\partial \mathcal{H}_C}{\partial I_s} &= \lambda_{22}(j+1)(\mu_s + du_3(j+1)) + \frac{b\beta(1-u_1(j+1))}{N}S(j+1)(\lambda_1(j+1)-\lambda_2(j+1)) \\
&\quad +\eta_1\frac{b\beta(1-u_1(j+1))}{N}I_h(j+1)(\lambda_7(j+1)-\lambda_8(j+1)) \\
&\quad +\eta_2\frac{b\beta(1-u_1(j+1))}{N}I_a(j+1)(\lambda_{13}(j+1)-\lambda_{14}(j+1)) - W_{10} \qquad (3)
\end{aligned}
$$

where

$$
\begin{aligned}
a_1 &= \beta_H(1-u_2)\frac{(N-I_H)}{N^2}, \quad b_0 = c\beta\alpha_S(1-u_1)\frac{(N-I_L)}{N^2},\\
a_2 &= \beta_H(1-u_2)\frac{(N-I_{HL_L})}{N^2}, \quad b_1 = c\beta\alpha_S(1-u_1)\frac{(N-I_D)}{N^2},\\
b_{01} &= c\beta\alpha_S(1-u_1)\frac{(N-I_P)}{N^2}, \quad a_3 = \beta_H(1-u_2)\frac{(N-I_{HD})}{N^2},\\
b_2 &= c\beta\alpha_S(1-u_1)\frac{(N-I_{HD})}{N^2}, \quad a_4 = \beta_H(1-u_2)\frac{(N-I_{HL})}{N^2},\\
b_3 &= c\beta\alpha_S(1-u_1)\frac{(N-I_{HL})}{N^2}, \quad a_5 = \beta_H(1-u_2)\frac{(N-I_{HP})}{N^2},\\
b_4 &= c\beta\alpha_S(1-u_1)\frac{(N-I_{HP})}{N^2}, \quad a_6 = \beta_H(1-u_2)\alpha_H\frac{(N-I_A)}{N^2},\\
a_7 &= \beta_H(1-u_2)\alpha_H\frac{(N-I_{AL_L})}{N^2}, \quad a_8 = \beta_H(1-u_2)\alpha_H\frac{(N-I_{AL})}{N^2},\\
b_5 &= c\beta\omega_S(1-u_1)\frac{(N-I_{AL})}{N^2}, \quad a_9 = \beta_H(1-u_2)\alpha_H\frac{(N-I_{AD})}{N^2},\\
b_6 &= c\beta\omega_S(1-u_1)\frac{(N-I_{AD})}{N^2}, \quad a_{10} = \beta_H(1-u_2)\alpha_H\frac{(N-I_{AP})}{N^2},\\
b_7 &= c\beta\omega_S(1-u_1)\frac{(N-I_{AP})}{N^2}, \quad c_1 = b\beta(1-u_1)/N.
\end{aligned}
$$

$$
\begin{aligned}
f_1 &= b\beta I_S(1-u_1),\\
f_2 &= c\beta(1-u_1)\left[(I_L+I_D+I_P)+\alpha_S(I_{HL}+I_{HD}+I_{HP})+\omega_s(I_{AL}+I_{AD}+I_{AP})\right],\\
f_3 &= \beta_H(1-u_2)\left[(I_H+I_{HL_L}+I_{HL}+I_{HD}+I_{HP})+\alpha_H(I_A+I_{AL_L}+I_{AL}+I_{AD}+I_{AP})\right].
\end{aligned}
$$

#### S.9 Proof of Theorem 6.1

To validate this theorem, we employ [4]. We note that the controls in this instance are not negative. In order to minimize the issue, the objective functional in (u1, u2, u3) must satisfy the required convexity. By definition, the collection of control variables, $u_1, u_2, u_3$ is convex. The integrand $W_1I_L + W_2I_{HL} + W_3I_{AL} + W_4I_D + W_5I_{HD} + W_6I_{AD} + W_7I_P + W_8I_{HP} + W_9I_{AP} + W_{10}I_S + W_{11}I_H + W_{12}I_A + \frac{1}{2}W_{13}u_1^2 + \frac{1}{2}W_{14}u_2^2 + \frac{1}{2}W_{15}u_3^2$. of the functional $J$ is convex on $\Gamma$, and its state variables are bounded. Since there are optimal controls for minimizing the convexity on $\Gamma$ (**??**),(**??**) We utilize Pontryagin's maximum principle to derive the necessary conditions to find the optimal solutions in the following manner, since there are optimal controls for minimizing the functional subject of systems. Assuming that $(z, u)$ represents the best solution to an optimal control problem, it follows that $\lambda = \lambda_1, \lambda_2, ...\lambda_i$ for $i = 1 \to 22$, a non-trivial vector function, must satisfy the following conditions:

$$
\frac{dz}{dt} = \frac{\partial \mathcal{H}_{\mathcal{C}}(t,z,u,\lambda)}{dt}, \quad 0 = \frac{\partial \mathcal{H}_{\mathcal{C}}(t,z,u,\lambda)}{\partial u} \text{ at } u^\star, \quad \frac{d\lambda}{dt} = \frac{\partial \mathcal{H}_{\mathcal{C}}(t,z,u,\lambda)}{\partial z}.
$$

#### S.10 Proof of Theorem 6.2

We use results in [3] and the previous theorem 6.1 to establish this theorem. Using the optimally condition: $\frac{\partial \mathcal{H}_{\mathcal{C}}}{\partial u_1} = 0$, $\frac{\partial \mathcal{H}_{\mathcal{C}}}{\partial u_2} = 0$, $\frac{\partial \mathcal{H}_{\mathcal{C}}}{\partial u_3} = 0$, we get,

$$
\frac{\partial \mathcal{H}_{\mathcal{C}}}{\partial u_1} = W_{13}u_1 + h_1S(\lambda_2-\lambda_1) + \eta_1h_1I_H(\lambda_8-\lambda_7) + \eta_2h_1I_A(\lambda_{14}-\lambda_{13}) + h_2S_S(\lambda_{21}-\lambda_{20})
$$

$$
\implies u_1 = \frac{h_1S(\lambda_1-\lambda_2) + \eta_1h_1I_H(\lambda_7-\lambda_8) + \eta_2h_1I_A(\lambda_{13}-\lambda_{14}) + h_2S_S(\lambda_{20}-\lambda_{21}}{W_{13}},
$$

$$\frac{\partial \mathcal{H_C}}{\partial u_2} = W_{14}u_2+h_3S(\lambda_7-\lambda_1)+\epsilon_2h_3L(\lambda_8-\lambda_2)+\epsilon_4h_3I_L(\lambda_9-\lambda_3)+\epsilon_1h_3I_D(\lambda_{10}-\lambda_4)+\epsilon_3h_3I_P(\lambda_{11}-\lambda_5)+h_3R(\lambda_{18}-\lambda_6),$$

$$\implies u_2 = \frac{h_3S(\lambda_1-\lambda_7)+\epsilon_3h_3L(\lambda_2-\lambda_8)+\epsilon_4h_3I_L(\lambda_3-\lambda_9)+\epsilon_1h_3I_D(\lambda_4-\lambda_{10})+\epsilon_3h_3I_P(\lambda_5-\lambda_{11}+h_3R(\lambda_6-\lambda_{18}}{W_14}.$$

$$\frac{\partial \mathcal{H_C}}{\partial u_3} = W_{15}u_3 - bS_S\lambda_{20} - bE_S\lambda_{21} - \lambda_{22}bI_S,$$

$$\implies u_3 = \frac{bS_S\lambda_{20}+bE_S\lambda_{21}+bI_S\lambda_{22}}{W_{15}}.$$

where

$$h_1 = \frac{-b\beta I_S}{N},\ h_2 = \frac{-c\beta\{(I_L+I_D+I_P)+\alpha_S(I_{HL}+I_{HD}+I_{HP})+\omega_S(I_{AL}+I_{AD}+I_{AP}\}}{N},$$

$$h_3 = \frac{-\beta_H\{(I_L+I_{HL_L}+I_{HL}+I_{HD}+I_{HP})+\alpha_H(I_A+I_{AL_L}+I_{AL}+I_{AD}+I_{AP}}{N}$$

Here 0 is the lower bound and 1 is the upper bound for the controls $u_i's$. This indicates that $u_1 = u_2 = u_3 = 0$ if $\tilde{u_1} < 0; \tilde{u_2} < 0; \tilde{u_3} < 0$ and $u_1 = u_2 = u_3 = 1$ if $\tilde{u_1} > 1;\ \tilde{u_2} > 1;\ \tilde{u_3} > 1$
Hence for these optimal controls $u_1^\star,\ u_2^\star,\ u_3^\star$ we have find the Optimum values of J.

### References


[1] Jack Carr. *Applications of centre manifold theory*, volume 35. Springer Science & Business Media, 2012.

[2] Carlos Castillo-Chavez and Baojun Song. Dynamical models of tuberculosis and their applications. *Mathematical Biosciences & Engineering*, 1(2):361–404, 2004.

[3] Suzanne Lenhart and John T Workman. *Optimal control applied to biological models.* Chapman and Hall/CRC, 2007.

[4] Lev Semenovich Pontryagin. *Mathematical theory of optimal processes.* Routledge, 2018.
